# VALORIS: A privacy-aware logistic regression method for vertically partitioned data within a novel privacy risk assessment framework

**DOI:** 10.1101/2025.07.15.25331536

**Authors:** Félix Camirand Lemyre, Marie-Pier Domingue, Jean-Philippe Morissette, Anita Burgun, Jean-François Ethier

**Affiliations:** Groupe de recherche interdisciplinaire en informatique de la santé (GRIIS), https://griis.ca/; Département de mathématiques, Faculté des sciences, Université de Sherbrooke, Sherbrooke, Canada; Département de médecine, Faculté de médecine et des sciences de la santé, Université de Sherbrooke, Sherbrooke, Canada; Health Data Research Network Canada; Institut Imagine, Université Paris Cité, Paris, France; Department of Medical Informatics, Necker Hospital, AP-HP, Paris, France

**Keywords:** Distributed analysis, Distributed algorithm, Vertically partitioned data, Logistic regression model, Health analytics, Statistical inference

## Abstract

Life sciences research increasingly relies on variables held by different entities, such as clinical, laboratory, environmental, and genomic data. Due to legal, ethical, and social acceptability constraints, these data often cannot be shared across organizations holding them. As a result, they cannot be pooled, and analyses must be conducted within the framework of vertically partitioned data. Supporting such analyses requires methods that protect privacy. However, the mere fact that line-level data are not exchanged should not be mistaken for true privacy protection. We introduce VALORIS (Vertically partitioned Analytics under the LOgistic Regression model for Inference in Statistics), a novel method that enables statistical inference under a logistic regression model without disclosing any individual-level data—including the outcome variable. VALORIS is a practical, communication-efficient algorithm that requires no third-party coordinator. Most importantly, it includes a novel framework for evaluating privacy, allowing users to distinguish among different levels of privacy preservation.

## 1 Introduction

Exploring new frontiers in life sciences research often requires integrating complementary types of data collected from a common set of observations to uncover novel relationships and identify opportunities for intervention. Variables of interest are frequently diverse in nature, such as clinical, genomic, laboratory, and socioeconomic data, and are typically held in different public or private organizations. This type of data fragmentation constitutes a vertical partitioning, where each data source holds different types of information about the same observations.

As an example, health research projects investigating outcome–covariate relationships among the Canadian population could require statistical analyses that combine, within a single statistical model, provincial medico-administrative data (held by organizations such as the Institut de la statistique du Québec or the Ontario Institute for Clinical Evaluative Sciences) with other sources, such as genomic data from the Canadian Partnership for Tomorrow’s Health (CanPath) cohort [1] or data from the Canadian Longitudinal Study on Aging (CLSA) [2]. However, legal, ethical, and social acceptability constraints prevent the pooling of such data in one organization or the other, an approach that is typically feasible only in smaller-scale projects where these constraints are sometimes more manageable. Commercial considerations may also impose additional limitations, as companies serving as data sources might restrict the transfer of their data. In all such cases, each organization must ensure that the sensitive data it holds cannot be accessed or inferred by unauthorized parties.

Methodologies that enable statistical analyses by coordinating computation across collaborative data-holding entities, without requiring line-level data to leave the environments in which they are held, are often grouped under the umbrella of distributed statistical analytics. Supporting large-scale collaborative projects falls within the mandate of the Health Data Research Network Canada (HDRN), which has established a dedicated work stream on distributed analysis.

Several distributed statistical inference approaches have been proposed for horizontally partitioned data [3], where different data holders each possess information on different observations but with the same set of variables, as well as for various types of vertically partitioned data [4–8]. This article focuses on the logistic regression model, which is widely used in life sciences research, and more specifically on parameter estimation and standard error computation from non-centralizable vertically partitioned data. These, in turn, enable confidence interval (CI) construction and hypothesis testing for the model parameters.

For the targeted techniques, the first criterion for HDRN endorsement is that the approach ensures privacy protection—specifically, avoiding the sharing of raw data outside the data nodes and providing formal confidentiality guarantees, including protection against reverse-engineering. It should also be communication-efficient, given that many data partners require human review of outgoing numerical outputs; and should produce results comparable to those from classical pooled maximum likelihood analyses (see, e.g., [9]), which is essential to build trust among users accustomed to centralized approaches. However, our literature review did not identify any method that meets all of these criteria.

To our knowledge, only one method has been proposed to perform maximum likelihood-based logistic regression inference without requiring raw data to leave its original environment [10, 11] (although standard errors, CIs and hypothesis testing are not explicitely addressed, the procedure can be extended with minor modifications to do so). However, as acknowledged by the original authors and noted in a review [12], the method inherently involves a large number of communication rounds and leaves unresolved some concerns about potential data leakage. These limitations make the approach unsuitable for our intended applications.

As far as we are aware, the only other method that provides the targeted statistical inference techniques and yields results comparable to classical pooled maximum likelihood analyses is the VERTIGO algorithm [13], along with its complementary extension VERTIGO-CI [14], which has been used in a work published in 2025 in Nature Communications [15]. However, this method requires sharing the outcome (that is, local raw data) to every participating node, which is inherently undesirable from a privacy standpoint. Moreover, it serves as a salient example of how, even in the absence of direct sharing of line-level data (in this case, covariate data), aggregated outputs can still be exploited by participating entities to reconstruct or infer sensitive line-level information through reverse-engineering techniques. We recently demonstrated that, in many practical situations, running the VERTIGO and VERTIGO-CI algorithms renders the raw covariate data reconstructible from the disclosed quantities [16]. This prior work from our group underscores that the distributed nature of a procedure does not inherently guarantee privacy protection (see also, e.g., [17]). For a method to be considered privacy-preserving, the numerical outputs shared during the analysis should be assessed to ensure they do not contain sufficient information to enable the reconstruction of raw data through reverse-engineering.

One challenge in doing so is that, to our knowledge, the literature on distributed statistical inference lacks both a formal framework for systematically assessing the privacy-preserving capabilities of a given method and, more broadly, clear guidelines on what it means for a method to be privacy-preserving.

Taking this into account, the aim of this work is to propose a novel privacy-aware approach for computing parameter estimates and their standard errors for a maximum-likelihood-based logistic regression model with vertically partitioned data. Our approach provides a clear and practical method for assessing whether, when deploying in a given setting our proposed algorithm, called VALORIS—Vertically-partitioned Analytics under the LOgistic Regression model for Inference in Statistics—, there is a risk that line-level data could be reverse-engineered. Specifically, it allows data nodes to determine whether line-level data can be guaranteed to remain protected from re-identification given the specific quantities disclosed, such as parameter estimates or their standard errors, based on the number and nature (binary or continuous) of the variables stored at each node. To address the absence of a formal privacy assessment framework, we define and examine two levels of privacy. The first level reflects a pattern we have consistently observed in the literature, where a method is typically labeled privacy-preserving if it operates in a distributed manner and avoids the direct transmission of line-level data, see e.g. [18]. The second level introduces a more stringent criterion: it additionally requires that line-level data cannot be reconstructed through reverse-engineering techniques.

To illustrate the feasibility and accuracy of our approach, we present two real-data examples in this paper. The first is a reproducible example using the MIMIC-IV database [19]. The second is a clinical case study that investigates potential factors associated with kidney failure in pediatric patients with chronic kidney disease (CKD), using de-identified structured data from Necker–Enfants Malades Hospital. This example, involving clinical and laboratory data, extends previous efforts aimed at improving the diagnosis and treatment of renal ciliopathy patients [20]. We also provide a third example through a publicly accessible online repository, featuring a synthetic dataset we created along with the accompanying R code for its analysis [21].

## 2 Results

### 2.1 Model settings

This work focuses on the binary logistic regression model with *p* covariates. Let Y ∈ {−1, 1} be a random variable representing the binary outcome, and let **X** ∈ ℝ^*p*^ denote the associated random vector of covariates (see Supplementary Tables 1 for detailed notation glossaries). The conditional distribution of Y given **X** = ***x*** is specified, for *y* ∈ {−1, 1}, as

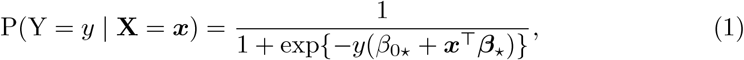

where *β*_0*⋆*_ ∈ ℝ and ***β***_*⋆*_ = [*β*_1*⋆*_, …, *β*_*p⋆*_]^⊤^ ∈ ℝ^*p*^ respectively denote the true (but unknown) intercept and covariate parameters. The data required for the analysis consist of a sample of *n* independent realizations of the random pair (**X**, Y), denoted by 𝒟 = {(***x***_1_, *y*_1_), …, (***x***_*n*_, *y*_*n*_)}, where each covariate vector is written as ***x***_*i*_ = [*x*_*i*1_, …, *x*_*ip*_]^⊤^. The vector of response variables is denoted by ***y*** = [*y*_1_, …, *y*_*n*_]^⊤^ {−1, 1} ^*n*^ and the covariate data matrix is denoted by ***X*** = [***x***_1_ …***x***_*n*_]^⊤^ ∈ ℝ^*n×p*^.

**Table 1.** Wald-type (1–α) CIs obtained at level α = 0.05 with MIMIC-IV database for a logistic regression model and *Death* as outcome.

| Covariate-node | Variable | Vertical Partition CI | Centralized CI<sup>1</sup> |
| --- | --- | --- | --- |
| 1 | Age | [0.022, 0.031] | [0.022, 0.031] |
| 1 | Myocardial infarction | [0.235, 0.557] | [0.235, 0.560] |
| 1 | Renal disease | [0.045, 0.342] | [0.046, 0.341] |
| 1 | Malignant cancer | [0.758, 1.042] | [0.761, 1.043] |
| 2 | Transport (helicopter) | [-0.700, 0.997] | [-0.620, 0.919] |
| 2 | Transport (walk-in) | [-0.376, -0.075] | [-0.376, -0.077] |
| 2 | Triage temperature | [-0.152, -0.037] | [-0.149, -0.041] |
| 2 | Triage sbp | [-0.006, 0.000] | [-0.006, 0.000] |
| 2 | Triage dbp | [-0.015, -0.005] | [-0.015, -0.005] |
| 2 | Triage respiratory rate > 20 | [0.301, 0.648] | [0.308, 0.643] |
| 3 | First-day urine output | [-0.001, 0.000] | [-0.001, 0.000] |
| 3 | First-day heart rate mean | [0.028, 0.036] | [0.028, 0.036] |
| 3 | First-day glucose mean | [0.001, 0.004] | [0.001, 0.004] |

<sup>1</sup>The CI's for the centralized setting were computed using the *glm* function in R with a non-penalized logistic regression model.

In the vertically partitioned data setting considered for this work, covariate information is partitioned across *K ≥* 2 distinct locations, referred to as *covariate-nodes*. Each covariate-node *k* ∈ {1, …, *K*} holds a matrix 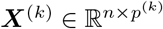 containing the values for all observations in the analytical dataset of *p*^(*k*)^ covariates distinct from those at other nodes, so that 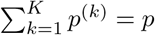. We assume that the columns of ***X*** are ordered so that the first *p*^(1)^ columns correspond to the covariates stored at covariate-node 1, the next *p*^(2)^ columns to those stored at covariate-node 2, and so on. The full matrix ***X*** can be written in block form as [***X***^(1)^, …, ***X***^(*K*)^].

We assume that the response vector ***y*** is stored at a single location, referred to as the *response-node*, and is not shared with the other nodes. Response-node and covariate-node are not mutually exclusive roles, such that a data-node can hold both the response and covariates. We also assume that datasets are aligned across all partitions ***X***^(1)^, …, ***X***^(*K*)^ and that this alignment corresponds to the structure of ***y***, i.e., Patient 1 appears in row 1 of all partitions and in the first entry of ***y***, and so forth.

### 2.2 Overview of the approach

To estimate (*β*_0*⋆*_, ***β***_*⋆*_), we consider for *λ >* 0 the following ridge-penalized logistic regression problem

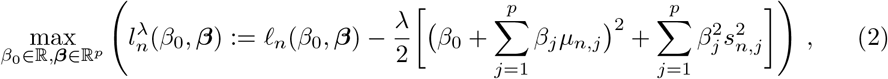

where *ℓ*_*n*_(*β*_0_, ***β***) denotes the unpenalized log-likelihood of the logistic regression model whose formula is given in Equation (9) in Methods 4.2, and *µ*_*n,j*_ and *s*_*n,j*_ are respectively the empirical mean and standard deviation of the *j*^th^ covariate, for *j* ∈ {1, …, *p*}.

In the objective function at (2), we choose *λ* sufficiently small to satisfy 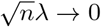 as *n* → ∞. This condition ensures that its maximizer is statistically equivalent to the (unpenalized) standard maximum likelihood estimate, in the sense that their difference becomes negligible relative to the sampling variability (of order 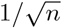) as the sample size increases (see Methods 4.2.2 and Supplementary Methods 2).

The maximization problem in (2) is solved numerically, with the solution denoted by 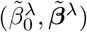. This is achieved by addressing its dual minimization formulation, given in (3) below, which is solvable in our vertically partitioned data setting. The algorithm takes as input a user-defined tolerance parameter *ϵ >* 0, which defines the stopping criterion for the dual optimization procedure and ensures an upper bound on the difference between the numerical and the exact solutions to (2) (see Methods 4.2.4).

#### 2.2.1 The new VALORIS algorithm

Our algorithm VALORIS begins by requiring every covariate-node *k* to compute the mean 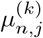 and standard deviation 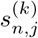 of each column in its covariate matrix ***X***^(*k*)^. Using these, the covariate-node constructs a centered and scaled version of the matrix, denoted by 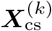, and then computes its local Gram matrix as 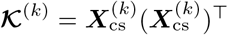. This matrix is then sent to the response-node.

Once these matrices have been received, the response-node numerically solves the dual minimization problem

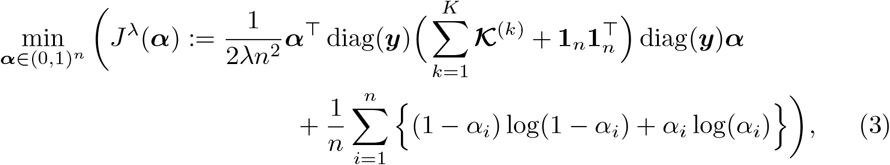

where **1**_*n*_ ∈ ℝ^*n*^ denotes the vector with all entries equal to one.

To solve (3), since ***α*** is restricted to the domain (0, 1)^*n*^ (see Methods 4.2.4), our implementation employs the Two-Metric Projected Newton method [22, 23].

Once a numerical solution 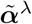 to (3) has been obtained, the response-node sends the following quantity to covariate-node *k*, for *k* ∈ {1, …, *K*}:

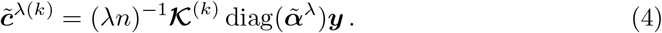

From this, each covariate-node *k* ∈ {1, …, *K*} retrieves the numerical estimates 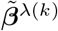, corresponding to the components of 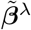 associated with the covariates stored at that node, by solving the following system of equations:

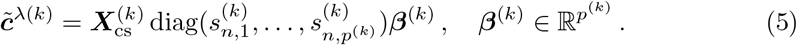

To enable covariate-nodes to compute standard errors, the response-node computes the matrix 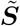 defined as

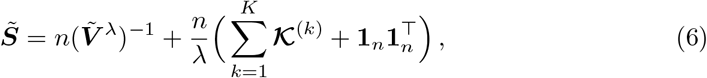

where 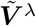 is a diagonal matrix whose entry (*i, i*) is given by

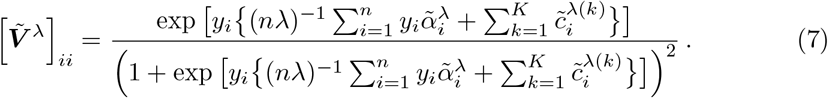

The response-node also generates a matrix 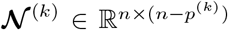 in the null-space of **𝒦** ^(*k*)^, and sends 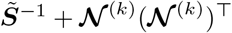 to each covariate-node *k* ∈ {1, …, *K*}.

To compute the standard error of 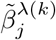, for *j* ∈ {1, …, *p*^(*k*)^}, covariate-node *k* uses the following expression:

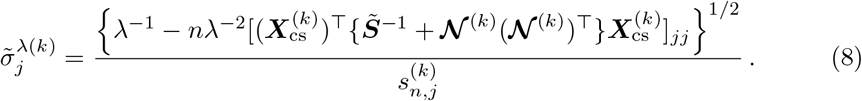

The rationale behind the algorithm and its theoretical correctness are detailed in Methods 4.2. The full VALORIS algorithm is stated in Algorithm 1.

##### Algorithm 1 VALORIS: Vertically partitioned Analytics under the LOgistic Regression model for Inference in Statistics

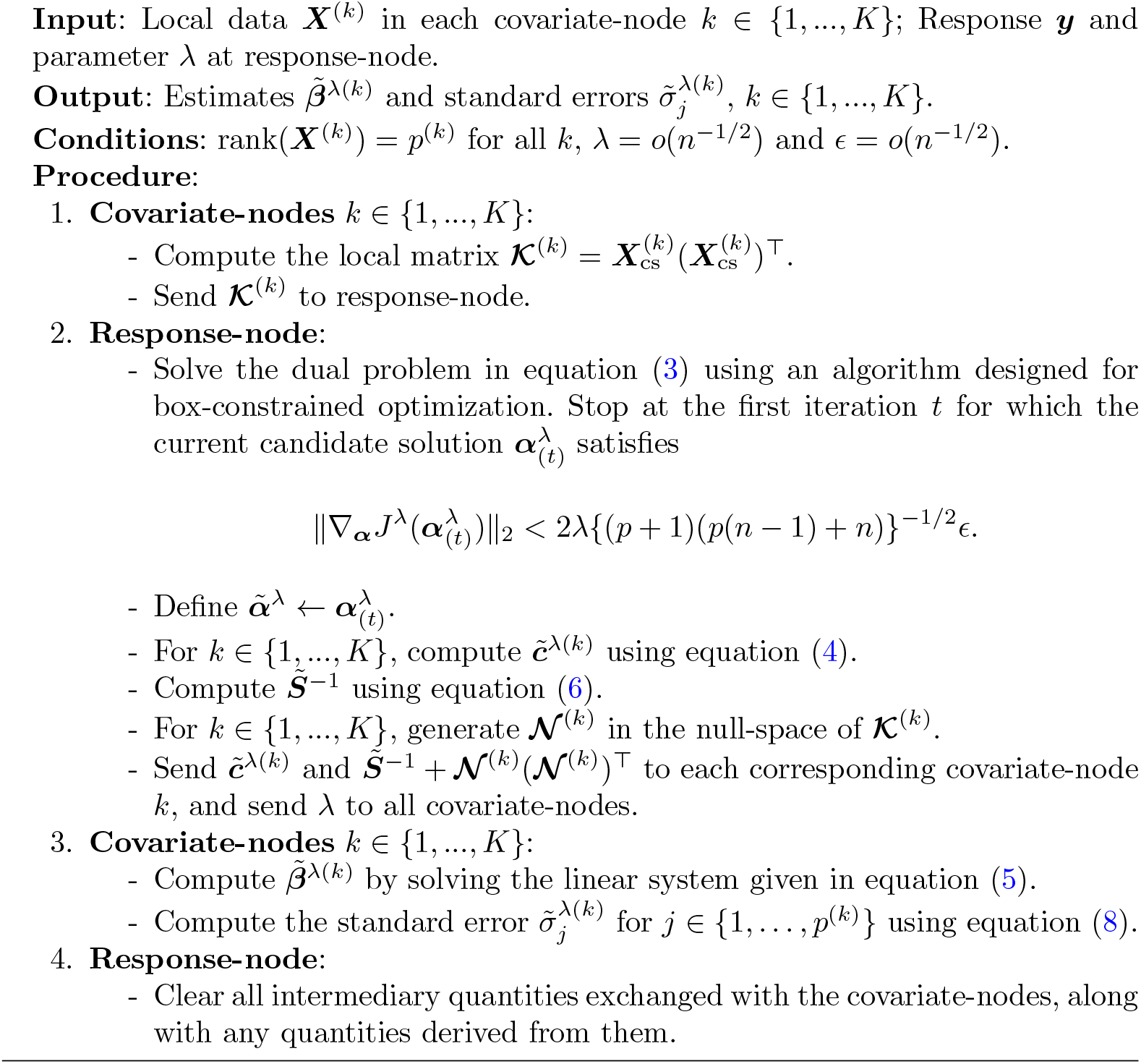

The algorithm was implemented in R [24] and implementation is available through the link in Supplementary Notes 1.

The response-node does not disclose either ***y*** or 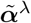 during the procedure, and the covariate-nodes are only required to share their local Gram matrices **𝒦**^(*k*)^. The workflow illustrating the information exchanged throughout the process is shown in Figure 1. A single round of communication between the covariate-nodes and the response-node is required to obtain the parameter estimates and their associated standard errors.

**Fig. 1.**
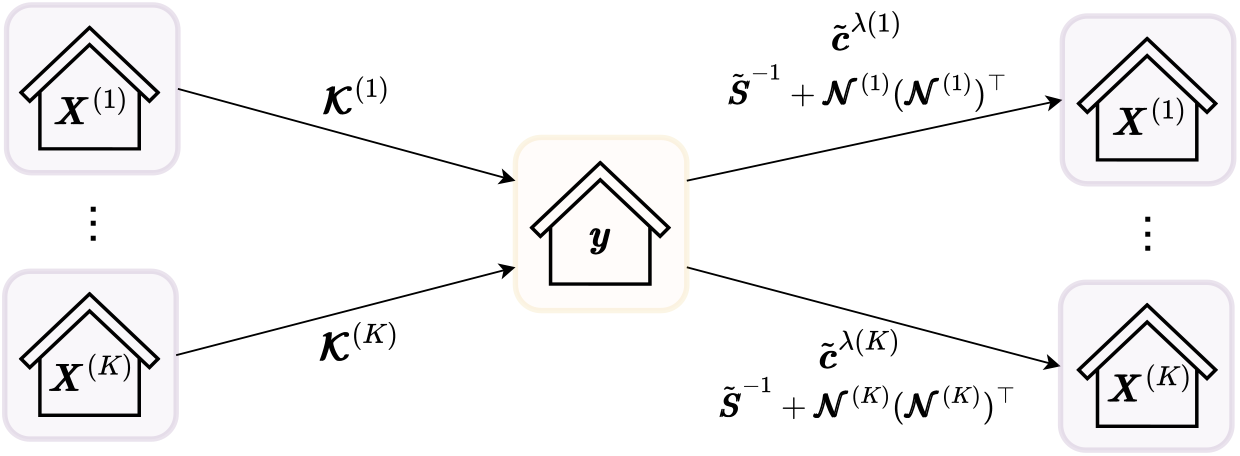
Workflow associated with running the VALORIS algorithm.

### 2.3 Privacy analysis

For covariate-nodes, our analysis focuses on the ability of the response-node to infer any entry of the scaled data matrices 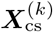; conversely, for the response-node, we assess the ability of a given covariate-node to infer any entry of the outcome vector ***y***.

We analyze privacy by investigating two levels of protection guarantees, reflecting the different interpretations of privacy that users may hold. Achieving **Privacy Level I** implies that no line-level data are explicitly exchanged as part of the approach. It can be seen from Algorithm 1 that the VALORIS algorithm neither requires nor involves the exchange of line-level data, thereby satisfying this level of privacy protection. As such, it will not be discussed further. Achieving **Privacy Level II** implies that line-level data cannot be uniquely recovered (or reverse-engineered) from the quantities exchanged or disclosed during the execution of the approach. This level of protection may depend on contextual factors such as variable types (e.g., continuous vs. binary) and the number of variables per data node. When this guarantee holds, it means that for every entry in the original dataset, there exists at least one *admissible candidate dataset*, consistent with the disclosed quantities, that differs at that entry. Admissible candidate datasets are defined as datasets that (a) are consistent with all exchanged and disclosed quantities available to any party that could, at a given time, attempt to reconstruct the original dataset, and (b) respect the nature of each variable (e.g., binary or continuous).

Privacy Level II may be achieved to varying extents, depending on the size and structure of the set of admissible candidate datasets. For example, the overall set may be infinite, yet the values taken by a specific variable (i.e., a covariate or the response, as recorded across all *n* observations) may still vary in only two ways across all admissible candidates. To formalize this, we define, for each variable in a dataset, its number of *admissible candidate configurations* as the number of admissible candidate datasets in which the values for that variable differ in at least one observation. Given a set of quantities that could be used to attempt reconstruction of the original dataset, we subclassify Privacy Level II among three *levels of indirection* representing the smallest number of admissible candidate configurations across all variables of the considered dataset: **Level II-(i)** if it is equal to 2; **Level II-(ii)** if that number is a finite number greater than 2; **Level II-(iii)** if it is infinite. Alternative definitions for the indirection levels could have been considered, see Methods 4.3.3.

Acknowledging the possibility of adversarial behavior by the response-node—such as retaining information, or retrieving and exploiting external data for re-identification purposes, we divide the privacy analysis into two subsections. Noting that Step 4 of Algorithm 1 specifies that all intermediary quantities exchanged with the covariate-nodes—along with any quantities derived from them—must be cleared, the analysis in Results 2.3.1 proceeds under the following Procedural Assumptions (PA#): (PA1) Algorithm 1 is executed as intended, and (PA2) the algorithm is run in an environment with no access to external information. The privacy analysis in Results 2.3.2 illustrates the impact on privacy loss when these assumptions are sequentially relaxed.

Throughout this assessment, we rely on a set of assumptions that are typically met in practice. Our analysis assumes that at least two covariate-nodes—including one potentially co-located at the response-node—participate in the analysis, and that the scaled covariate values of each individual *i* ∈ {1, …, *n*} that are stored at node *k* are all different, that is, that 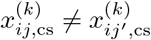 for *j* ≠ *j*^*′*^. It is also assumed that the sample size is such that *p*(*p* + 3)*/*2 *< n*, and that there at least exists two observations such that *y*_*i*_ = 1 and two such that *y*_*i*_ = −1.

#### 2.3.1 Privacy analysis of Algorithm 1 under Procedural Assumptions (PA1) and (PA2)

Our analysis treats covariate-nodes data and response-node data separately.

##### Privacy analysis for covariate-nodes data

In our algorithm, each covariate-node *k*, for all *k* ∈ {1, …, *K*}, shares with the response-node its local Gram matrix 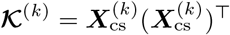. Once Algorithm 1 has been executed, covariate-nodes may wish to disclose to all parties a subset—or all—of the parameter estimates and standard errors, or alternatively, two-tailed p-values. The primary risk of re-identification arises from the possibility that the response-node could reverse-engineer covariate-node *k*’s line-level data by combining the disclosed quantities with the information it possesses at the time of their disclosure. Under Procedural Assumptions (PA1) and (PA2), this entails evaluating the risk that the response-node may be able to re-identify 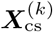 based on, in turn, **𝒦**^(*k*)^ and a subset—or all—of the parameter estimates, standard errors, or two-tailed *p*-values.

If a node serves as both the response-node and a covariate-node, then it has direct access to the line-level data for the covariates it holds, and the latter re-identification risk does not apply to this node.

We present all settings that were mathematically shown to achieve Privacy Level II using decision trees that incorporate the corresponding indirection levels, under the assumption that at least one covariate-node—other than the one possibly located at the response-node—holds at least one continuous covariate.

The trees presented in Figure 2 indicate which analysis results can be disclosed under each setting, given the associated indirection level. Since the impact of disclosed quantities differs depending on the presumably known support of the covariates, we use one decision tree that pertains to binary covariates, and another for continuous covariate. For covariate-nodes holding both types, each tree is to be applied separately according to the number of covariates of each type. The resulting indirection levels may differ between types; for instance, Level II-(iii) may be reached for continuous covariates, while a lower level may apply to the binary ones.

**Fig. 2.**
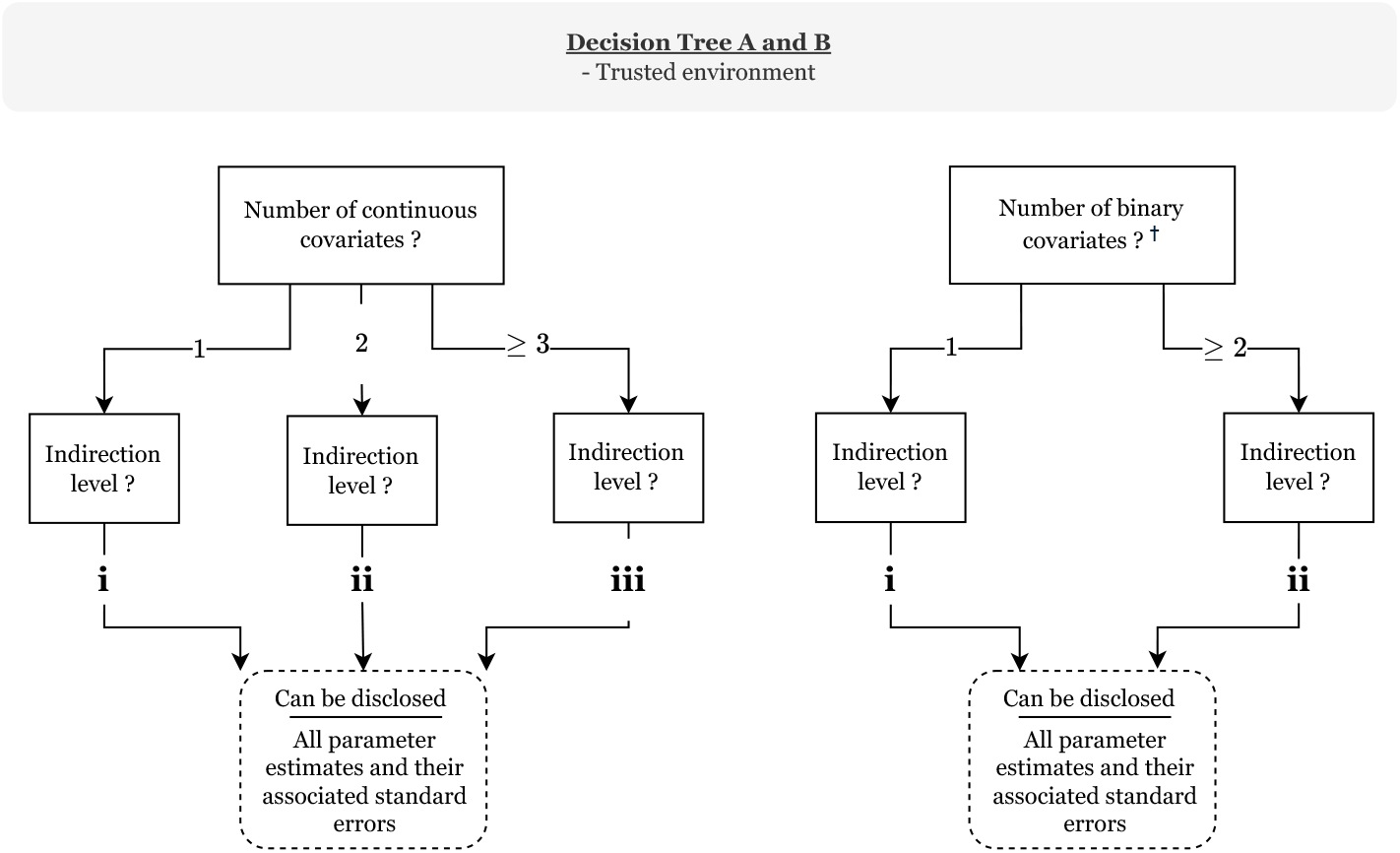
Decision trees for covariate-nodes to follow before using VALORIS under trusted environment, i.e. under Assumptions (PA1) and (PA2). ^†^At least one covariate-node—other than the one possibly located at the response-node—holds at least one continuous covariate.

If the setting required for an analysis is not represented in a tree, this means no guarantee regarding the reach of Privacy Level II can be provided in this setting when using the VALORIS algorithm. The theoretical arguments supporting their construction are provided in Methods 4.3.1.

In cases where all covariate-nodes—except possibly the one located at the response-node—contain only binary covariates, Privacy Level II may not be attained.

##### Privacy-preserving analysis for the response-node data

In our algorithm, the response-node shares with each covariate-node *k*, for all *k* ∈ {1, …, *K*}, the vector 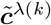 and the matrix 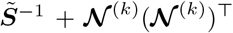. Therefore, the primary risk of re-identification arises from the possibility that a given adversarial covariate-node could reverse-engineer the response vector ***y*** from those quantities.

Knowledge of 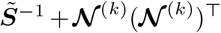, whether or not accompanied by 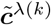, does not provide any exploitable information for reverse-engineering.

As for 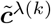, a necessary condition for achieving Privacy Level II is that the sample size exceeds the total number of covariates held at covariate-node *k*; otherwise, reverse-engineering is feasible. However, this condition is not sufficient to ensure that both values −1 and 1 can be considered admissible for every entry of the response-vector, and it remains unclear whether a general theoretical condition—valid across all datasets—can be established to guarantee resistance to reverse-engineering. We consequently developed an empirical criterion, in the form of a verification algorithm that can be executed at the response-node prior to transmitting 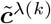, to ascertain whether Privacy Level II is satisfied and to provide a lower bound on the number of admissible candidate configurations for the response vector. This criterion is developed under the assumption that at least one continuous covariate is held outside the adversarial covariate-node (see section Methods 4.3.2). An implementation of this criterion is available through Supplementary Notes 1. Based on numerical simulations, the empirical criterion is likely to be satisfied when the sample size is sufficiently large relative to the number of covariates at node *k*—for example, when *n ≥* 100 and *p*^(*k*)^ *≤* 10. When this criterion is met, Privacy Level II-(ii) is achieved.

If the response-node also serves as a covariate-node, the assumption that at least one continuous covariate is held outside the adversarial covariate-node ensures that all parameter estimates and their associated standard errors can be disclosed while maintaining Privacy Level II for all *p*^(*k*)^ *≥* 1, with indirection Level II-(ii) for continuous covariates and Level II-(ii) for binary covariates. The latter assumption is satisfied if the response-node also serves as a covariate-node including one continuous variable.

In the absence of continuous covariates held outside the adversarial covariate-node—i.e., when all other covariate-nodes contain only binary covariates—Privacy Level II may not be attained, since the adversarial node could theoretically enumerate all possible datasets, and in some cases, only one may be consistent with the quantities it possesses.

#### 2.3.2 Privacy analysis of Algorithm 1, with step-wise alleviation of Procedural Assumptions (PA1) and (PA2)

##### Privacy analysis of Algorithm 1, alleviating Procedural Assumption (PA1)

Since relaxing (PA1) does not affect the privacy assessment of the response-node’s data, we focus on the case of data held by covariate-nodes. To illustrate the impact of relaxing Procedural Assumption (PA1), while maintaining Procedural Assumption (PA2), we repeated the analysis from Results 2.3.1 to assess the re-identification risk for a covariate-node’s data, assuming that intermediary quantities exchanged with the covariate-nodes, as well as any quantities derived from them, were not cleared at the end of Algorithm 1. The resulting decision trees are presented in Figure 3.

**Fig. 3.**
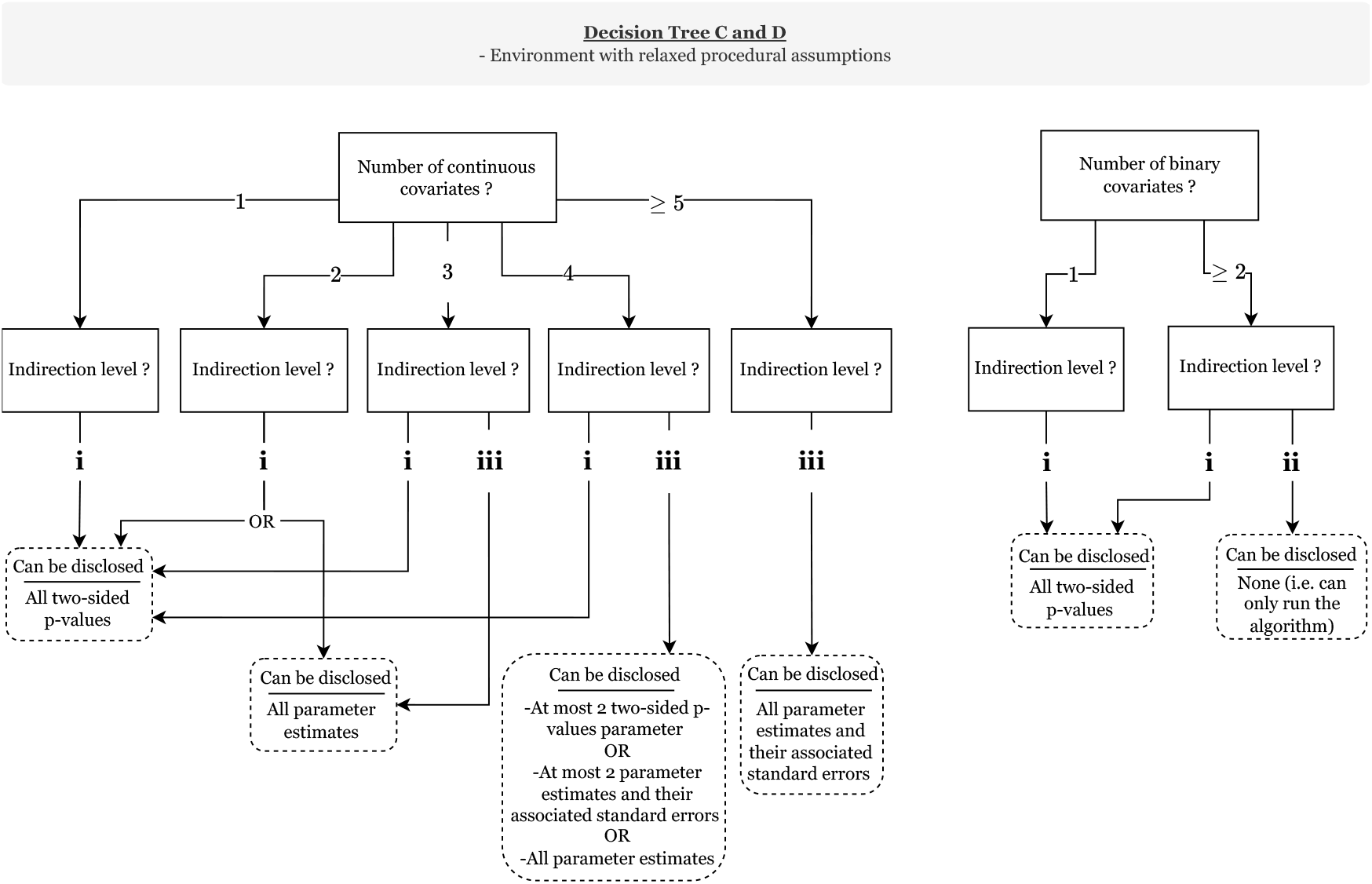
Decision trees for covariate-nodes to follow before using the VALORIS algorithm under reduced assumptions environment, i.e. under Assumption (PA2), alleviating Assumption (PA1).

##### Assessment of privacy loss in examples where Procedural Assumptions (PA1) and (PA2) are alleviated

External information can take many forms, and a privacy analysis is required to account for the specific characteristics of each. We illustrate this through two scenarios in which external information concerning covariate means is accessed: (a) when the exact values of the covariate means are known (e.g. the data come from population-based sources or cohorts with publicly available covariate means, or the analysis reuses data from previous studies in which means were disclosed); and (b) when the exact values are not known, but relationships between them are (e.g., a covariate representing a rare disease expected to have a lower prevalence than a covariate representing sex).

In the case of binary covariates, because of the binary support and the size and structure of the associated indirection level, knowledge of the mean as per scenario (a) generally allows the response-node to directly recover line-level data. In scenario (b), if a single binary covariate representing a rare event is held at a given covariate-node, its associated column can often be identified among the limited set of possibilities defined by sign-flips and permutation. In contrast, for continuous covariates, information regarding the mean does not enable the recovery of line-level data when the indirection level is *Infinite*. When the covariate-node holds exactly two continuous covariates and parameter estimates are disclosed, the response-node can reconstruct the uncentered and unscaled covariate data matrices up to column permutations, and may potentially infer line-level data if one obtained column is incompatible with the known covariate support (e.g., negative values for age).

### 2.4 Application on real health data

We provide two application examples of our approach using covariate splits designed to reflect realistic vertical partitions. Parameter estimates and standard errors were computed in the vertically partitioned setting using the proposed method VALORIS, and then compared with those obtained from a non-penalized logistic regression in a centralized setting, performed in R (version 4.4.1) [24]. The empirical criterion regarding the privacy-preserving properties for the response-vector was met in both applications.

#### 2.4.1 MIMIC-IV database

A total of 13 covariates and one outcome were selected from the MIMIC-IV database [19], vertically partitioned across three modules treated as distinct data nodes. The Hospitalization (HOSP) module was assigned the outcome variable *death* and four covariates. The Emergency Department (ED) and Intensive Care Unit (ICU) modules served as additional covariate-nodes, contributing six and three covariates, respectively. Only complete-case observations were retained, resulting in a sample size of *n* = 13677.

CIs were computed using our proposed method with *λ* = *n*^−1^ and *ϵ* = 5*n*^−1^. The results are presented in Table 1.

#### 2.4.2 Kidney failure among children patients with chronic kidney disease

In this application, we investigated potential associations between kidney failure within two years of baseline and various characteristics in a cohort of children with CKD. To guide our analysis, we drew on a study that proposed two Cox models to predict kidney failure in adults with CKD: a model with four baseline covariates, and a model that included four additional serum measurements [25, 26]. In practice, serum measurements may be stored in separate databases that are potentially non-centralizable and inaccessible for research purposes.

The cohort included pediatric patients from Necker–Enfants Malades Hospital (Paris, France) with potential CKD stage 3 or 4 at baseline and complete data (*n* = 97). Data were partitioned across two nodes: one with the outcome *kidney failure at two years after baseline* and the covariates *age, sex, estimated glomerular filtration rate (eGFR)* and *urine albumin-creatinine ratio (uACR)*; the other containing serum measurements.

For our algorithm, we set *λ* = 10^−4^ and *ϵ* = *n*^−1^. The results are provided in Table 2.

**Table 2.** Logistic regression model parameter estimates obtained with kidney failure at two years after baseline as outcome.

| Covariate-node | Variable | Vertical Partition $\hat{\beta}^\lambda(\hat{\sigma}^\lambda)$ | Centralized $\hat{\beta}(\hat{\sigma})^1$ |
| --- | --- | --- | --- |
| 1 | Age | 0.051(0.059) | 0.051(0.059) |
| 1 | Sex (male) | -0.773(0.533) | -0.773(0.533) |
| 1 | eGFR | -0.008(0.027) | -0.008(0.027) |
| 1 | uACR | -0.206(0.161) | -0.206(0.161) |
| 2 | Serum albumin | -0.645(0.439) | -0.645(0.439) |
| 2 | Serum bicarbonate | -0.100(0.081) | -0.100(0.081) |
| 2 | Serum calcium | 0.345(0.524) | 0.345(0.524) |
| 2 | Serum phosphate | 0.519(0.289) | 0.519(0.289) |

A log-transformation was applied to uACR due to a skewed distribution [26].
<sup>1</sup>The estimates for the centralized setting were computed using the *glm* function in R with a non-penalized logistic regression model.

## 3 Discussion

The new method VALORIS enables statistically valid estimations of parameters and their standard errors in logistic regression models using vertically partitioned data. Unlike existing methods, it does not require the response variable to be shared across all parties, nor the disclosure of individual probabilities of having the response equal to 1, while also providing transparency regarding the risk of re-identifying line-level information. A single back-and-forth round of communication between the covariate-nodes and the response-node is required, making the method well suited for contexts in which data nodes must manually review the exchanged quantities.

Our privacy analysis considers various scenarios in which different sets of numerical results (parameter estimates, standard errors, p-values) are disclosed at the end of the procedure, across settings that vary in both the number and nature of the covariates, and the decision trees we provided are meant to be screened by users each time they intend to apply the VALORIS algorithm. Our procedure enables covariate-nodes to make informed decisions about which quantities to disclose, based on the associated privacy risks, given the procedural assumptions they are comfortable adopting and the desired indirection level to achieve. This privacy-awareness sets this method apart from previous approaches to statistical analysis using logistic regression with vertically partitioned data.

Making procedural assumptions is essential—and central—to conducting a privacy analysis with mathematical guarantees, as done in this work. A setting not being shown to achieve Privacy Level II does not necessarily imply that line-level data would be recoverable by an unauthorized party. In some cases, this outcome reflects the inability to provide mathematical guarantees due to the complexity of the equations involved under the given assumptions. Each application context should therefore carefully evaluate which assumptions are applicable and conduct its privacy analysis.

External information that pertains to a dataset can sometimes be known or available from outside sources, and its nature impacts the privacy risk assessment that must be conducted. In this paper, we illustrated this by evaluating the re-identification risk to a covariate-node’s data in scenarios where the response-node has access to covariate means. Other types of external information would necessitate a distinct re-identification risk assessment, but the framework presented here could be reused to address this by treating such external sources as covariate-nodes.

Obviously, the privacy assessment provided in this paper does not cover all scenarios that may arise in practice. For example, while discrete covariates with finite support can be conservatively assessed using the binary case as a reference, those with more than two observed values may provide greater privacy protection. The current assessment serves as a conservative foundation, but more specific scenarios—particularly when a covariate-node holds a mix of discrete and continuous covariates—warrant further investigation that could lead to less restrictive settings.

The results in Tables 1 and 2 show that the estimates and standard errors—or equivalently, the CIs—are nearly identical between the vertical and centralized settings, as expected given the theoretical demonstrations provided below. These findings highlight the accuracy of the proposed method for the specified values of *ϵ* and *λ* relative to the centralized benchmark. The CKD application serves as a compelling proof of concept for analysis with vertically partitioned data. It motivates further investigation into factors associated with kidney failure in pediatric cohorts, particularly by incorporating additional covariates distributed across different entities. For example, integrating genomic data from a national cohort with clinical data from hospital information systems could help identify renal ciliopathy cases among CKD patients, which could improve understanding of condition-specific risk factors and support targeted interventions.

Among the numerical and computational limitations, our current implementation of the algorithm can lead to time-consuming computations at the response-node for large sample sizes *n*, due to some operations involving *n × n* matrices. Other box-constrained algorithms could be investigated to optimize those operations. In addition, the current version of the method is limited to complete-case analyses, and further development is needed to incorporate a strategy for handling missing data.

## 4 Methods

The Methods section is organized as follows. Methods 4.1 introduces the main notations used throughout the paper (complemented by glossaries in Supplementary Tables 1). The core methodology underlying the algorithm—including parameter estimation and standard error computation—is presented in Methods 4.2. Since parameter estimation involves solving a dual optimization problem, Methods 4.2.4 provides additional details on the dual estimation procedures, with a focus on the stopping criterion. The final two sections address the privacy analysis methodologies: Methods 4.3.1 covers the covariate-nodes’ data, and Methods 4.3.2 addresses the response-node’s data. Supporting mathematical derivations are provided in the Supplementary Methods.

### 4.1 Notation

In this work, non-italic and non-italic bold letters respectively represent random variables and random vectors. Lowercase italic bold letters denote vector-valued quantities, while uppercase italic bold are used for matrices. The notation *a*_*j*_ represents the *j*^th^ component of a vector ***a*** ∈ ℝ^*p*^, and the operator diag(***a***) refers to the *p × p* diagonal matrix obtained with the entries of ***a*** on the diagonal. We define the norm 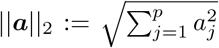. For iterative computations over ***a***, the step count *t* appears as an index in parenthesis (e.g. ***a***_(*t*)_). For a function *f*_***θ***_ depending on ***θ***, the gradient (column-vector) and Hessian with respect to ***θ*** are noted *∇*_***θ***_*f*_***θ***_ and 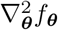. We write *a*_*n*_ = *o*(*b*_*n*_) if *a*_*n*_*/b*_*n*_ →− 0 as *n* →− ∞.

We recall that *µ*_*n,j*_ and *s*_*n,j*_ represent the mean and standard deviation of the *j*th column in ***X***, that is,

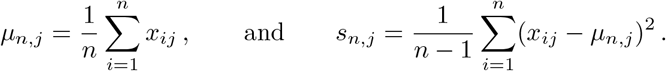

The vector ***x***_*i*,cs_ denotes the centered and scaled covariates for observation *i*, with entries *x*_*ij*,cs_ = (*x*_*ij*_ − *µ*_*n,j*_)*/s* _*n,j*_.

In Results 2.1, we used 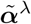 to denote the (approximate) numerical solution to (3) and, although not explicitly stated, we used a tilde to denote quantities computed from 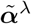 —such as 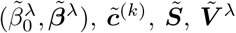, and 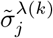. In the Methods, we adopt a different notation to refer to the exact solution of (3), which will be denoted by 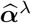, and we use a hat to indicate quantities derived from it (e.g., 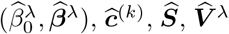, and 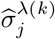). We make this distinction to separate numerical approximations from theoretical quantities used in theoretical analyses, and to explicitly define and track the approximation error (see Methods 4.2.4).

### 4.2 Theoretical arguments supporting the validity of the VALORIS algorithm

Methods 4.2.1 briefly recalls how the unknown parameters (*β*_0*⋆*_, ***β***_*⋆*_) of the logistic regression model defined in (1) are typically estimated from the data in 𝒟 in a pooled, centralized setting, along with the corresponding computation of their standard errors. This is followed by an overview of the main similarities and differences between our proposed method and existing approaches for logistic regression with vertically partitioned data. We then provide the methodological details underlying the parameter estimation procedure and standard errors computations in Methods 4.2.2 and 4.2.3 respectively.

At the heart of our algorithm is the dual optimization program in (3). In standard primal-dual optimization settings where strong duality holds—that is, when the optimal values of the primal and dual objectives coincide—the convergence of a dual algorithm is often assessed by comparing the values of the primal and dual objectives at a given iteration. However, this is not feasible in our vertically partitioned setting, as the dual optimization is performed at the CC, which does not have access to the covariate data and thus cannot evaluate the primal objective. To address this, we propose in Methods 4.2.4 an original stopping criterion for the dual algorithm, which is integrated in the VALORIS algorithm, that ensures an upper bound on the difference between the candidate primal solution—reconstructed from the dual variables—and the true primal optimum.

#### 4.2.1 Background on logistic regression and positioning of the proposed method

When the data in 𝒟 are centralized to a single data node, (*β*_0*⋆*_, ***β***_*⋆*_) are typically estimated by solving the following log-likelihood maximization problem:

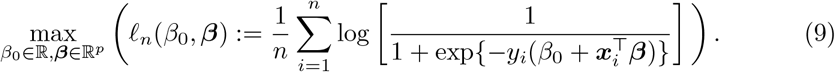

The solutions 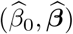 of the latter problem, called the maximum likelihood estimates, are generally found using a Newton-Raphson algorithm or a variant of it. The variance-covariance matrix of the maximum likelihood estimates 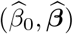 can be estimated by the inverse of the observed Fisher information matrix, defined as

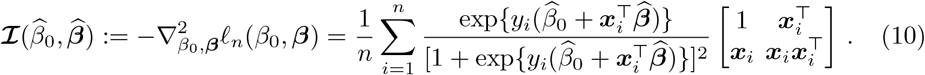

Standard errors of parameter estimates are computed upon extracting the diagonal entries of 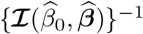, taking the square-root and multiplying it by *n*^−1*/*2^.

In the literature, computing 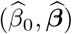 and their standard errors in a vertically partitioned setting has been addressed in [10, 11], using an approach based on “secure sums” and “secure matrix product” algorithms. As mentioned in the introduction, this approach involves a high volume of communication between nodes and does not eliminate privacy risks, making it unsuitable for our intended applications.

Our approach is different, and approximates 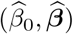 by solving a penalized version of (9), whose dual formulation enables computation in a vertically partitioned setting using a single round of communication. The penalty parameter is chosen to be sufficiently small so that the resulting estimates remain statistically equivalent to 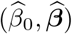.

Dual optimization for logistic regression with vertically partitioned data has previously been used in the VERTIGO algorithm [13] and in the paper from our group revisiting the VERTIGO algorithm [16], in the context of ridge regression (focusing on point estimates without CIs). The related VERTIGO-CI algorithm in [14] also uses this approach for logistic regression with CIs, also relying on a small penalty parameter as we do in this paper. However, aside from the fact that as noted in the introduction their approach allow for reverse-engineering of covariate-nodes data, the penalized version of (9) considered in that work differs from ours, and their methodology lacks the theoretical justification that we provide in this section for appropriately choosing *λ*.

In contrast to existing approaches based on dual optimization, our method does not require sharing the response vector across data nodes. Additionally, our procedure for enabling covariate-nodes to estimate their covariate-related parameters and compute their corresponding standard errors is fundamentally different from all previously proposed methods.

#### 4.2.2 Computing parameter estimates

To estimate (*β*_0*⋆*_, ***β***_*⋆*_) in our vertically partitioned setting, our approach relies on approximating (9) with the penalized version 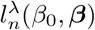 defined in (2) for *λ >* 0, and to consider 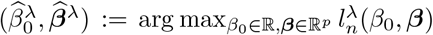. Under appropriate assumptions, if *λ* is chosen to be sufficiently small and defined as a function of *n* such that *λ* = *λ*(*n*) = *o*(*n*^−1*/*2^), then the approximation error incurred by using 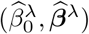 in place of the standard maximum likelihood estimator 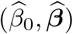 becomes negligible relative to the sampling variability. That is, under such conditions, 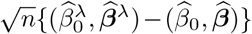 converges in probability to 0 (see Supplementary Methods 2 for a proof).

Recall *J*^*λ*^(***α***) defined in (3), which represents the dual problem to the maximization problem in (2). For any *λ >* 0, the solution 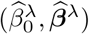 can be computed as

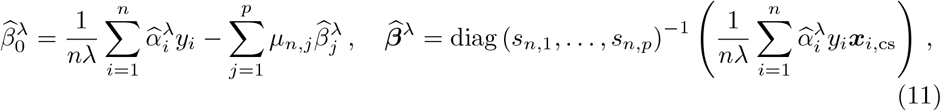

where 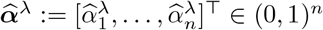 is the unique minimizer of *J*^*λ*^ (***α***) over (0, 1)^*n*^ (see Supplementary Methods 1, Proposition S1 for a proof).

As can be seen from the expression of *J*^*λ*^(***α***), it depends on the covariate data only through 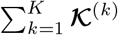, where 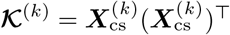 denotes the local Gram matrices. To solve (3) in our vertically partitioned setting, each covariate-node *k* ∈ {1, …, *K*} is required to compute and send the matrix 𝒦^(*k*)^ to the response-node. The response-node can then solve the minimization problem on its own and obtain 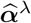 (see details in Methods 4.2.4).

From this, the response-node cannot compute 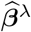 using the relationship in (11), as this quantity depends on the covariate data in a form that cannot be reconstructed from the entries of the matrices **𝒦**^(*k*)^. Recalling 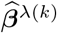 denotes the *p*^(*k*)^ components of 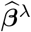 corresponding to the covariates stored at node *k*, we note from (11) that 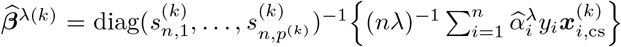. However, because the covariate-nodes in our framework do not have access to the entries of the response vector ***y***, even if the response-node would send 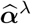 to the covariate-nodes, the latter does not possess all the necessary information to compute 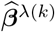 using this relationship as in [13, 16].

In light of these observations, to enable each covariate-node to compute 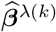, we require the response-node to send a distinct vector to each covariate-node which allows to construct a system of equations whose unique solution is 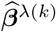. To see how this is done, noting from expressing 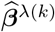 in a vector-matrix notation that

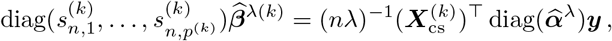

it follows that 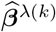 satisfies

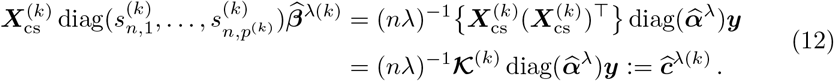

If 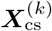 has full column rank, it follows that 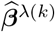 is the unique solution of the system of equations

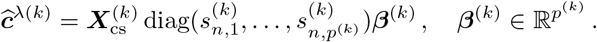

#### 4.2.3 Computing standard errors of parameter estimates

In our vertically partitioned setting, the observed Fisher information matrix 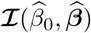 defined in (10) cannot be directly computed, as the estimates 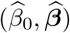 themselves are not accessible.

Alternatively, we show in Supplementary Methods 2 that, under the assumption used above that *λ* is chosen to be sufficiently small and defined as a function of *n* such that *λ* = *o*(*n*^−1*/*2^), it holds for *j* ∈ {2, …, *p*+1} that 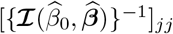 is asymptotically equivalent to 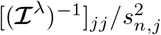, with

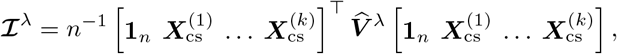

where 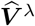 is a diagonal matrix whose diagonal entries 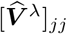 satisfy

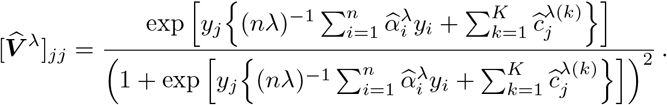

Therefore, when *λ* is small, the standard error of each component *j* of 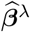 can be consistently estimated using *s* _*n,j*_ and the (*j* + 1)^th^ diagonal entry of {**ℐ** ^*λ*^}^−1^. However, even if the response-node is able to compute 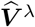, the expression for **ℐ**^*λ*^ poses a challenge; it is not straightforward to identify a suitable strategy that would allow either the response-node, which lacks access to each ***X***^(*k*)^, or any of the covariate-node, which lack access to 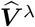 as well as to data matrices from other covariate-nodes, to compute these standard errors.

Our approach to addressing this challenge builds on the Woodbury matrix identity [27]. This result states that, given a *p×p* invertible matrix ***A***, a *n×n* invertible matrix ***C***, and *p × n* matrices ***U*** and ***W***, if ***C***^−1^ + ***W A***^−1^***U*** is invertible, we have

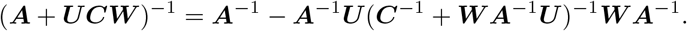

For any *η >* 0, and letting ***I***_*r*_ denote the ℝ *×* ℝ identity matrix, the Woodbury matrix identity allows to express {**ℐ**^*λ*^ + *η****I***_*p*+1_}^−1^ as

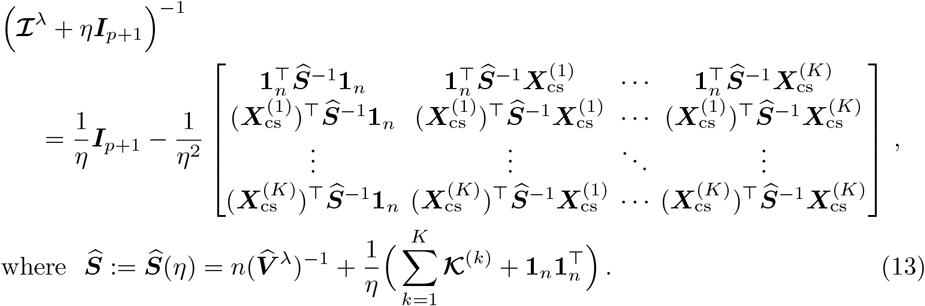

The parameter *η* should be chosen small enough so that each entry of the matrix (**ℐ**^*λ*^ + *η****I***_*p*+1_)^−1^ is approximately equal to the corresponding entry of (**ℐ**^*λ*^)^−1^, but large enough to ensure that the expression in (13) remains numerically stable. We experienced good numerical results with the choice *η* = *λ/n*.

The diagonal entries of {**ℐ**^*λ*^ +*η****I***_*p*+1_}^−1^ can be extracted from the terms of the form 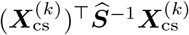. As the matrix ***Ŝ***^−1^ can be entirely computed at the response-node, it can be computed and sent to each covariate-node *k* to allow them to obtain the diagonal entries of (**ℐ**^*λ*^ + *η****I***_*p*+1_)^−1^ corresponding to their covariate data by computing the diagonal entries of 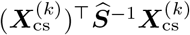.

In addition, we note that, for any matrix 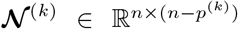 in the null-space of 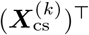 (i.e. such that 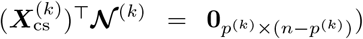, it holds that 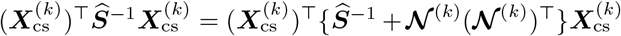. Since both expressions yield identical results, we choose to share the intermediate quantity ***Ŝ***^−1^ + **𝒩**^(*k*)^(**𝒩** ^(*k*)^)^⊤^ with the covariate-nodes instead of transmitting ***Ŝ***^−1^. The quantity **𝒩**^(*k*)^ can be generated at the response-node from the null-space of **𝒦** ^(*k*)^ because the null-space of (***X***^(*k*)^)^⊤^ is the null-space of ***X***^(*k*)^(***X***^(*k*)^)^⊤^ [28]. Although ***Ŝ***^−1^ could have been shared directly, we adopt a preventive approach by transmitting ***Ŝ***^−1^ + **𝒩**^(*k*)^(**𝒩** ^(*k*)^)^⊤^ instead, to mitigate the risk of information leakage from ***Ŝ***^−1^, as discussed in Methods 4.3.1. Similar strategies for protecting exchanged quantities have been proposed in prior work, including [6].

Covariate-node *k* can use the simplified expression in (8) to compute the standard errors of the parameter estimate 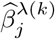 for *j* ∈ {1, …, *p*^(*k*)^}.

#### 4.2.4 Algorithm for solving the dual optimization program and its stopping criterion

As mentioned in Results 2.2.1, the response-node solves the minimization problem in (3) to obtain 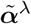 from which the 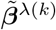 ‘s are calculated. The details of the box-constrained convex optimization method used to compute 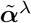 in our implementation are provided in Supplementary Methods 3. We now present the stopping criterion used in the VALORIS algorithm, which is independent of the specific optimization method employed, and which ensures that 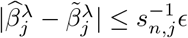 for all *j* ∈ {1, …, *p*}.

The stopping criterion to the minimization problem in (3) is based on the norm of the gradient of *J*^*λ*^(***α***). Specifically, if 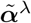 denotes a candidate solution obtained after a given number of iterations, this candidate is accepted as the final output if

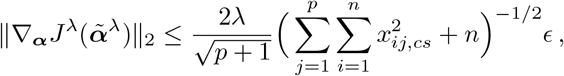

where the gradient of the dual objective function *J*^*λ*^(***α***) in (3) is given by

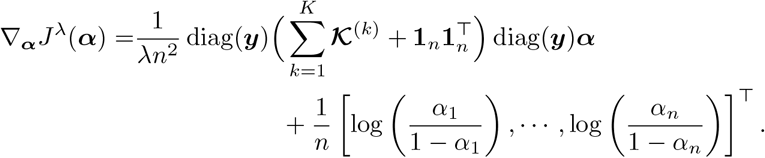

The condition can be further simplified using the relationship 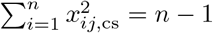 for all *j* ∈ {1, …, *p*}. This leads to

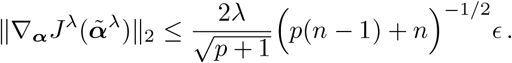

As we prove in Supplementary Methods 3 (see Section 3.2 therein), this entails the bound 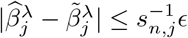 for all *j* ∈ {1, …, *p*}.

### 4.3 Methodology for conducting the privacy analysis

Recall from Results 2.3 that, to acknowledge the possibility of adversarial behavior by the response-node—such as retaining information, or retrieving and exploiting external data for re-identification purposes, a privacy analysis divided into two parts was carried out. The first part, whose results are presented in Results 2.3.1, proceeds under the Procedural Assumption (PA1) stating that Algorithm 1 is executed as specified, including Step 4, which requires the response-node to clear all intermediary quantities exchanged with the covariate-nodes—along with any quantities derived from them, and Procedural Assumption (PA2) stating that the algorithm is run in an environment with no access to external information. The second part, presented in Results 2.3.2, illustrates the impact on privacy loss when Procedural Assumption (PA1) is no longer upheld while (PA2) is still assumed, and subsequently when both Procedural Assumptions (PA1) and (PA2) are relaxed.

The methodology underlying this privacy analysis is divided into two subsections. Methods 4.3.1 examines the privacy-preserving properties of quantities shared by a covariate-node that is not co-located with the response-node. Methods 4.3.2 focuses on the privacy-preserving properties of quantities shared by the response-node, including the case where the response-node also serves as a covariate-node. Across both subsections, we develop and apply a methodological framework based on the notion of *admissible candidate datasets*, introduced at the beginning of Results 2.3—that is, admissible candidate matrices for covariate data and admissible candidate vectors for the response vector. Methods 4.3.3 then links these subsections to the results presented in Results 2.3.

Recall from Results 2.1 and Algorithm 1 (see also Methods 4.1) that the VALORIS algorithm uses the approximate solution 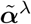 to (3) to derive several quantities, some of which are exchanged or disclosed. Also recall from Methods 4.1 the notation 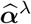 for the exact solution to (3), as well as the notation used for the quantities derived from it, introduced to distinguish theoretical values used in analysis from their numerical approximations. For the purposes of the privacy analysis, we assume that the theoretical solution 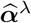 and its derived quantities are used, exchanged, and disclosed throughout. In addition to facilitating analytical derivations, this assumption provides an upper bound on potential privacy loss, as numerical implementations may introduce small perturbations that, in some cases, obscure exact reconstruction without eliminating the underlying risk.

#### 4.3.1 Methodology to assess the privacy-preserving properties of quantities shared by a given covariate-node not co-located with the response-node

Consider a given covariate-node *k* ∈ {1, …, *K*}, and recall that its local data matrix and its scaled version are denoted by ***X***^(*k*)^ and 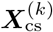 respectively. Our analysis focuses on the potential reconstruction of the scaled data matrix 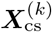, even though access to 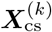 does not necessarily permit recovery of the original data matrix ***X***^(*k*)^ (it does so in the case of binary covariates, but not necessarily for continuous covariates—see below). This methodological choice makes the results for continuous covariates more robust to adversarial nodes accessing external information regarding either covariates mean or covariates standard deviation, and thereby represents an additional layer of privacy protection.

To derive the results related to the privacy analysis of the entries of 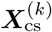 under each set of assumed Procedural Assumptions, we begin by examining the response-node’s ability to re-identify the entries of 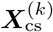 based on the set of quantities or information available to it at a given point in time during the execution of Algorithm 1. Each set of assumed Procedural Assumptions determines a different set of quantities or information that may become simultaneously available to the response-node. The results of the privacy analysis are derived by examining the stage (or stages) at which this set contains the most information, that is:

- Under Procedural Assumptions (PA1) and (PA2), the response-node has temporary access to the Gram matrix 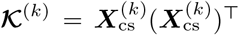, before **𝒦**^(*k*)^ and any quantities derived from it are cleared in Step 4 of the algorithm. Then, it may have access to 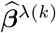, the 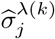 ‘s, or a subset of these quantities, depending on whether covariate-node *k* chooses to disclose them. Accordingly, two stages are examined: the stage before **𝒦**^(*k*)^ is cleared, and the stage at the end of the estimate disclosure process.
- In the scenario where Procedural Assumption (PA1) is no longer assumed but (PA2) is, the response-node has access to the Gram matrix **𝒦**^(*k*)^, in addition to 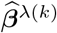, the 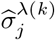’s, or a subset of these quantities, depending on whether covariate-node *k* chooses to disclose them. Accordingly, a single stage is examined—namely, the point at the end of the estimate disclosure process.
- In the illustrative scenarios where Procedural Assumptions (PA1) and (PA2) are no longer assumed, the analysis assumes that the response-node has access to the Gram matrix **𝒦**^(*k*)^, as well as to 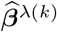, the 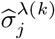 ‘s, or a subset of these quantities, depending on whether covariate-node *k* chooses to disclose them, in addition to the external information described in Results 2.3.2. A single stage is examined, the point at the end of the estimate disclosure process.

In studying these scenarios, our analysis aims to assess whether, when attempting to solve for 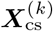 using the disclosed quantities and available information, whenever applicable, the solution space—defined as the set of data matrices compatible with the available quantities or information—contains, for every entry of 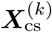, at least two *admissible candidate* matrices that differ at that entry. We define an admissible candidate matrix ***A*** for 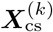 as one that satisfies the following two conditions: (a) the disclosed quantities could have been equivalently computed from ***A*** in place of 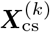, in which case ***A*** is called a *candidate* matrix for 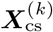; and (b) there exists a matrix ***A***_0_ such that for each *j* ∈ {1, …, *p*^(*k*)^}, all entries in column *j* of ***A***_0_ lie in the support 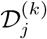 of covariate *j*—e.g., {0, 1} for binary covariates and ℝ for continuous ones, and such that ***A*** is a column-wise centered and scaled version of ***A***_0_, in which case ***A*** is said to be *admissible*.

Throughout our analysis, we will use the fact that if a matrix ***A*** is a candidate matrix for 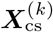 and 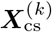 contains continuous covariates, then ***A*** is also an admissible candidate matrix for 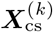, as the structure of ***X***^(*k*)^ imposes no constraints on the de-scaled and de-centered version of ***A***. However, this is not the case when 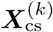 contains binary covariates. In this case, since the entries of the matrix 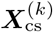 have the form

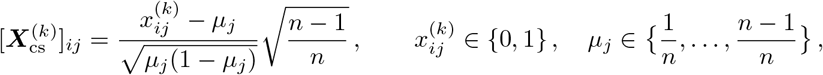

the requirement for a candidate matrix ***A*** to be admissible for 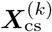 is that there exists 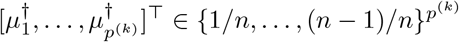 such that

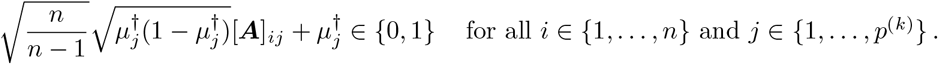

This requirement often narrows substantially the set of candidate matrices for 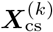, compared to the continous covariates case.

Our privacy analysis pertaining to each of the above scenarios builds on a mathematical characterization of the set of admissible candidate matrices for 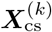, given the availability to the response-node, of:

1. the local Gram matrix **𝒦**^(*k*)^;
2. the local Gram matrix **𝒦**^(*k*)^ and a subset—or all— of the 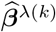’s;
3. the local Gram matrix **𝒦**^(*k*)^ and a subset—or all— of the 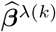’s with their corresponding standard errors;
4. the local Gram matrix **𝒦**^(*k*)^ and a subset—or all— of the two-sided p-values;
5. a subset—or all— of the 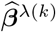’s with their corresponding standard errors, without the local Gram matrix **𝒦**^(*k*)^ and quantities derived from it.

In what follows, we provide the characterization of each set of admissible candidate matrices, along with the underlying mathematical foundations. We then describe how the decision trees were derived from these characterizations.

We analyze binary and continuous covariates separately. That is, throughout the privacy analysis, each matrix ***X***^(*k*)^ is assumed to contain either only binary covariates or only continuous covariates. In practice, a covariate-node, say node *k*, may hold both types. In such cases, the privacy analysis can still be applied by considering separately the subsets of binary covariates ***X***^(*k*,bin)^ and continuous covariates ***X***^(*k*,cont)^ held at the same node, and independently assessing the privacy risks associated with the quantities disclosed from each subset, as if they were held by distinct covariate-nodes. Proceeding in this way yields to conservative privacy risk assessments, since, for instance, the local Gram matrices **𝒦**^(*k*,bin)^ = ***X***^(*k*,bin)^(***X***^(*k*,bin)^)^⊤^ and **𝒦**^(*k*,cont)^ = ***X***^(*k*,cont)^(***X***^(*k*,cont)^)^⊤^ are not transmitted separately to the response-node; instead, only their sum, **𝒦**^(*k*)^ = **𝒦**^(*k*,bin)^ +**𝒦**^(*k*,cont)^, is disclosed. We proceed in this way because it remains unclear, at this stage, under which conditions the above decomposition is unique, and thus whether there is a risk that the response-node could uniquely recover **𝒦**^(*k*,bin)^ and **𝒦** ^(*k*,cont)^from the aggregate matrix **𝒦**^(*k*)^.

##### 1. When only the local Gram matrix is available to the response-node

Let ℳ_*n,p*_(ℝ) be the set of *n × p* real-valued matrices. When the local Gram matrix **𝒦**^(*k*)^ is the only quantity shared from covariate-node *k* to the response-node, reverse-engineering 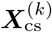 requires the response-node to solve for 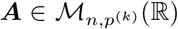 the system of equations **𝒦**^(*k*)^ = ***AA***^⊤^, under the constraints that ***A*** has empirical mean zero (i.e., ***A***^⊤^**1**_*n*_ = 0) and empirical variance one (i.e., 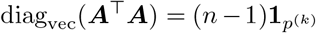, where diag_vec_(***Z***) denotes the vector of diagonal entries of any square matrix ***Z***). To analyze the ability of the response-node to do so, let

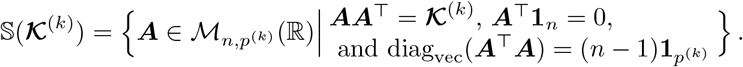

The set 𝕊 (**𝒦**^(*k*)^) consists of all candidate matrices for 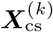 from which the disclosed quantity **𝒦**^(*k*)^ could have been equivalently computed. The following proposition provides a complete characterization of the matrices belonging to 𝕊 (**𝒦**^(*k*)^) (see Supplementary Methods 4.1 for a proof).

###### Proposition 1.

*Assume that* 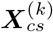 *has full column rank. Then*, ***A*** ∈ 𝕊 (**𝒦**^(*k*)^) *if and only if* 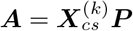, *with* 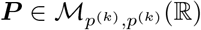 *an orthogonal matrix satisfying*

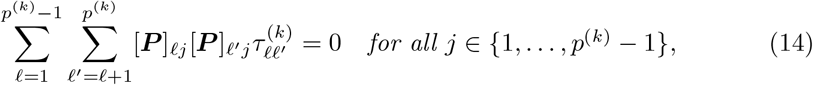

*where* 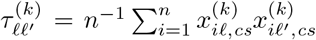 *corresponds to the entry* (*ℓ, ℓ*^*′*^) *of the correlation matrix*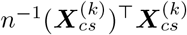.

It follows from Proposition 1 that 𝕊 (**𝒦**^(*k*)^) can be equivalently expressed as

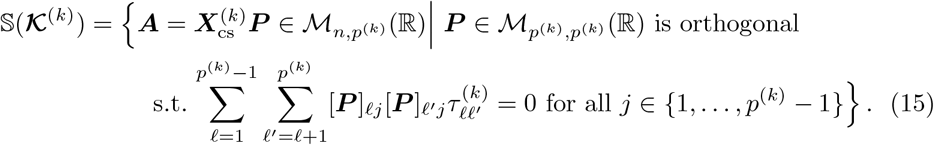

This proposition enables us to identify settings that ensure that, for all (*i, j*) ∈ {1, …, *n*} *×* {1, …, *p*^(*k*)^}, the solution space 𝕊 (**𝒦**^(*k*)^) contains at least two matrices ***A*** and ***A***^*′*^ such that 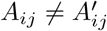. Consider the matrix 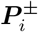, defined as the *p*^(*k*)^ *×p*^(*k*)^ identity matrix except for its *i*^th^ diagonal entry, where the value 1 is replaced by −1. Then, the matrix 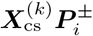 corresponds to a version of 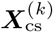 where the sign of each entry in the *i*^th^ column has been flipped. Since for all *i* ∈ {1, …, *p*^(*k*)^}it is straightforward to verify that 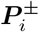 is an orthogonal matrix satisfying (14), we conclude from Proposition 1 that 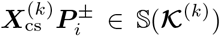. We conclude that if **𝒦**^(*k*)^ is the only quantity shared by covariate-node *k* with the response-node, then no restriction needs to be imposed on *p*^(*k*)^ to ensure that the solution space 𝕊 (**𝒦**^(*k*)^) contains at least two distinct matrices ***A*** and ***A***^*′*^ such that 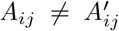 for all (*i, j*) ∈ {1, …, *n*} *×* {1, …, *p*^(*k*)^}. Indeed, because the matrices 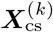 and 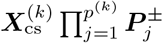 both belong to 𝕊 (**𝒦**^(*k*)^), the existence of such distinct candidates is guaranteed. We note that 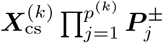 is an admissible candidate irrespective of the support for the covariates.

We next investigate the structure of the matrices belonging to 𝕊 (**𝒦**^(*k*)^), considering different cases based on the value of *p*^(*k*)^. When *p*^(*k*)^ = 1, we directly derive 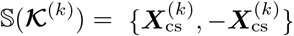. When *p*^(*k*)^ *≥* 2, any sign-permutation matrix 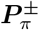, that is, any matrix with exactly one nonzero entry in each row and each column, where each nonzero entry is either 1 or −1, can be shown to satisfy 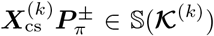, with 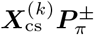 always being an admissible candidate for 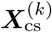. In the specific case where *p*^(*k*)^ = 2, since any 2 *×* 2 orthogonal matrix ***P*** := ***P***_*θ,d*_ has the form

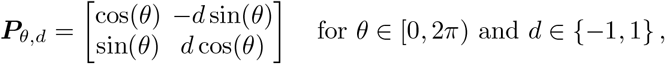

it follows from Proposition 1 that any matrix ***A*** in the set 𝕊 (**𝒦**^(*k*)^) can be written in the form 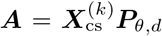 where the values of *θ* and *d* satisfy 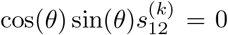. Therefore, if the columns of 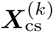 are not orthogonal, i.e. if 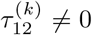, the only values of *θ* leading to 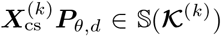 are {0, *π/*2, *π*, 3*π/*2}, for any *d* ∈ {−1, 1}, which corresponds to the case where 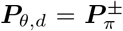, for a given sign-permutation matrix 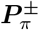. Therefore, one concludes that, in the case *p*^(*k*)^ = 2 and 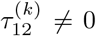, ***A*** is a candidate matrix for 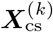 in the set 𝕊 (**𝒦**^(*k*)^) if and only if 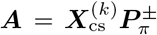. This implies that, upon disclosure of **𝒦**^(*k*)^, the response-node can only determine 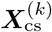 up to the signs or the ordering of its columns.

The case *p*^(*k*)^ *≥* 3 provides a setting where 𝕊 (**𝒦**^(*k*)^) often contains infinitely many distinct candidate matrices of the form 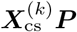. This is because, when *p*^(*k*)^ *≥* 3, and according to Proposition 1, the number of equations that ***P*** must satisfy for 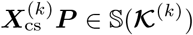 is smaller than the dimension of the space of *p*^(*k*)^ *× p*^(*k*)^ orthogonal matrices. Consequently, by appropriately parameterizing this space and formulating the constraints from Proposition 1 as equations on the associated parameters, the Implicit Function Theorem [29] can be used to establish the existence of infinitely many solutions. The logic behind this argument is illustrated in the case *p*^(*k*)^ = 3 in Supplementary Methods 4.3, which also contains the proof of the following result that formally establishes this claim.

###### Proposition 2.

*Assume that p*^(*k*)^ *≥* 3, *and that* 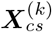 *has full column rank. Moreover, for* 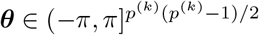, *let*

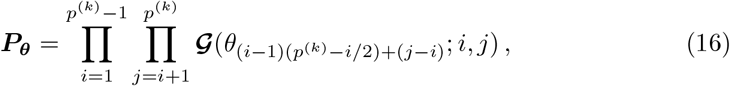

*where, for* 1 *≤* 𝕀 *< j ≤ p*^(*k*)^ *and* 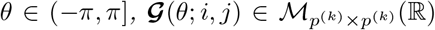 *denotes the Givens rotation matrix (see e*.*g. [30]) with entries* 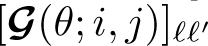 *defined as*

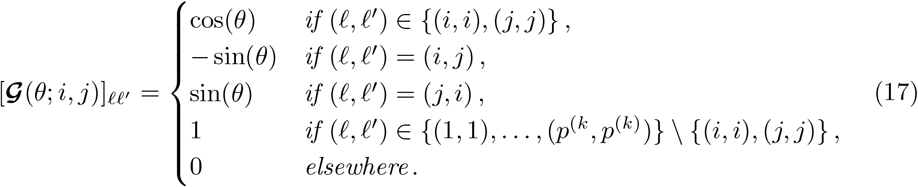

*Consider* 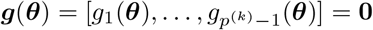, *where, for j* ∈ {1, …, *p*^(*k*)^ − 1},

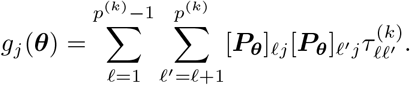

*Then*, 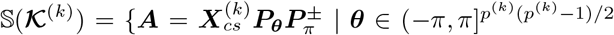 *and* ***g***(***θ***) = **0**}. *More-over, the cardinality of the set* 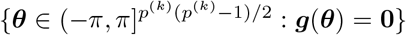 *is infinite, which implies that the cardinality of the set* 𝕊 (**𝒦**^(*k*)^) *is also infinite*.

###### Additional considerations for binary covariates

While Proposition 1 implies that any matrix ***A*** in the set 𝕊 (**𝒦**^(*k*)^) can be written in the form 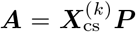, where ***P*** is an orthogonal matrix satisfying (14), it does not, however, ensure that ***A*** is an admissible candidate for 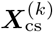. From the beginning of Methods 4.3.1, for a matrix ***A*** of binary covariates to be an admissible candidate for 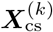, there must exist a vector of column means 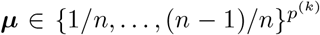 such that, for each *j*, all entries in column *j* of the matrix ***A*** diag(***s***) + **1**_*n*_***µ***^⊤^ lie within 𝒟_*j*_, the support of covariate *j*, with 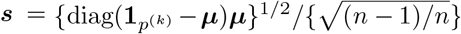, where ***a***^1*/*2^ represents the resulting vector when extracting the square root of each element of ***a***. This implies that each column of 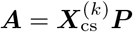 must take values in a set containing only two distinct elements. However, we show in Supplementary Methods 4.2 that, if 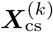 has 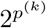 distinct rows, then any orthogonal matrix ***P*** such that 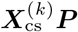 has only two distinct values per column must be a sign-permutation matrix. Therefore, when 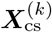 is derived from binary covariates and has 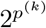 distinct rows, the only admissible candidate matrix ***A*** for 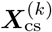 are of the form 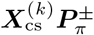.

Table 3 summarizes the solution sets of admissible candidates for 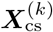, in the case of covariates held by a covariate-node *k* ∈ {1, …, *K*} that is not co-located with the response-node.

**Table 3.** Solution sets of admissible candidates for 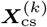, depending on the number of covariates at site *k* when **𝒦** ^(*k*)^ shared.

| $p^{(k)}$ | Continuous covariates<sup>1</sup> | Binary covariates<sup>1</sup> |
| --- | --- | --- |
| 1 | $\{-\mathbf{X}_{\text{cs}}^{(k)}, \mathbf{X}_{\text{cs}}^{(k)}\}$ | $\{-\mathbf{X}_{\text{cs}}^{(k)}, \mathbf{X}_{\text{cs}}^{(k)}\}$ |
| 2 | $\{\mathbf{X}_{\text{cs}}^{(k)} \mathbf{P}_{\pi}^{\pm}\}$ | $\{\mathbf{X}_{\text{cs}}^{(k)} \mathbf{P}_{\pi}^{\pm}\}$ |
| $\geq 3$ | $\{\mathbf{X}_{\text{cs}}^{(k)} \mathbf{P}_{\boldsymbol{\theta}} \mathbf{P}_{\pi}^{\pm}\}$, infinitely many $\boldsymbol{\theta}$ | $\{\mathbf{X}_{\text{cs}}^{(k)} \mathbf{P}_{\pi}^{\pm}\}$ |

<sup>1</sup>These represent the most restrictive theoretical solution sets that could arise. That is, in practice, the sets may include additional solutions depending on the structure of $\mathbf{X}_{\text{cs}}^{(k)}$ , particularly in cases where some covariates are orthogonal.

##### 2. When only the local Gram matrix and parameter estimates are available to the response-node

When the local Gram matrix **𝒦**^(*k*)^ is shared from covariate-node *k* to the response-node, along with a selected subset *J ⊆* {1, …, *p*^(*k*)^} of the components of the estimated parameters 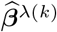 (which may include all components), reverse-engineering 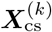 would require the response-node to search for a matrix ***A*** ∈ 𝕊 (**𝒦**^(*k*)^) that satisfies

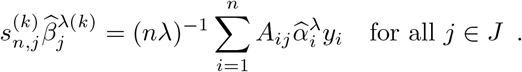

The values 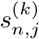 are not available to the response-node. We first focus on the case of continuous covariate data, and comments will be provided at the end of this subsection for the case of binary covariates. Since 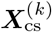 is computed from continuous covariate data, the local Gram matrix **𝒦**^(*k*)^ contains no information about their value: any choice for 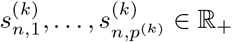 in the previous equation results in the same **𝒦**^(*k*)^. For a candidate matrix ***A*** for 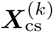 to be such that the disclosed quantities **𝒦**^(*k*)^ and 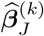 (where, throughout, for any vector ***a, a***_*J*_ denotes the subvector of ***a*** containing the entries indexed by *J*) could have been equivalently computed from either ***A*** or 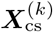, the response-node must identify a matrix ***A*** for which there exists a set of constants {*a*_*j*_ : *j* ∈ *J*} with strictly positive entries (*a*_*j*_ *>* 0 for all *j*) such that

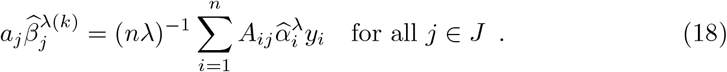

Given that all candidate matrices for 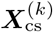 are of the form 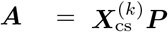 (recall Equation (15)), which implies 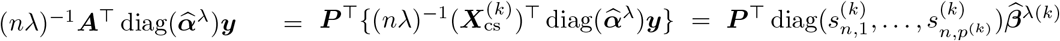, (18) holds if an only if 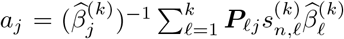 for all *j* ∈ *J*. The solution space of candidate matrices ***A*** for 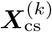 is therefore given by

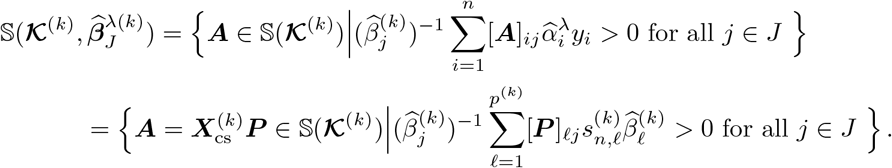

Once the response-node has identified a matrix ***A*** ∈ 𝕊 (**𝒦**^(*k*)^), it can readily construct from it a matrix ***A***^*′*^ (possibly equal to ***A***) such that 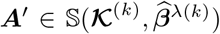. To illustrate this, take 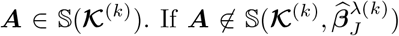, there exists a set of indices *J*^*′*^ *⊆ J* such that 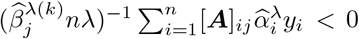 for all *j* ∈ *J*^*′*^. Since flipping the signs of all entries in any given column of a matrix in 𝕊 (**𝒦**^(*k*)^) yields another matrix that also belongs to 𝕊 (**𝒦**^(*k*)^), it follows that the matrix 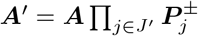 belongs to 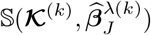.

In light of these observations, we draw the following conclusions from the preceding paragraph. <u>In the case *p*^(*k*)^ = 1</u>, since we have seen that 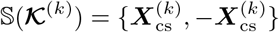, for any ***A*** ∈ 𝕊 (**𝒦**^(*k*)^), only one of ***A*** and 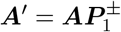 will be in 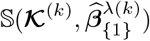. We conclude that, in this case, 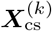 is the only element of 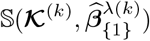. <u>In the case *p*^(*k*)^ = 2</u>, when 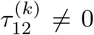, we have also seen that 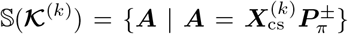. Therefore, in this case, we have 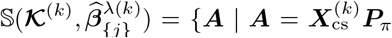, or 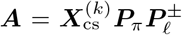 with *ℓ*≠ *j*,},where ***P***_*π*_ denotes a permutation matrix (i.e., either the identity matrix, or the matrix that permutes the two columns), and 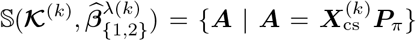.<u>In the case *p*^(*k*)^ ≥ 3</u>, for any *J ⊆* {1, …, *p*^(*k*)^}, it follows from the discussion above that the set 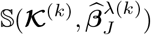 admits an infinite number of solutions whenever 𝕊 (**𝒦**^(*k*)^) does. From Proposition 2, the full column rank of 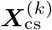 is sufficient to ensure the existence of infinitely many such solutions.

###### Additional considerations for binary covariates

Recall that when ***X***^(*k*)^ contains only binary entries—so that each column of 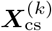 takes on exactly two distinct values—and if 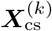 (or equivalently, ***X***^(*k*)^) has 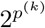 distinct rows, then any admissible candidate matrix ***A*** ∈ 𝕊 (**𝒦**^(*k*)^) must be of the form 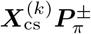. This implies that if the response-node is able to identify a single admissible candidate in 𝕊 (**𝒦**^(*k*)^), it can compute all candidates in 𝕊 (**𝒦**^(*k*)^). Also, note that since all columns of 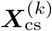 are centered and scaled, the proportion of positive entries in the *j*th column of 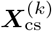 is equal to 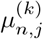, and the standard deviation of the covariate in column *j* can therefore be computed as

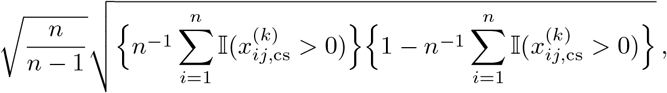

where 𝕀(*B*) is the indicator function taking value 1 if *B* is true, and 0 otherwise. Then, recalling equation (18), if 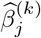 is disclosed for all *j* ∈ *J*, the response-node can narrow its search for 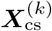 by identifying which admissible candidate ***A*** ∈ 𝕊 (**𝒦**^(*k*)^) satisfies the following equality:

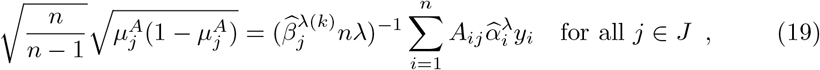

With 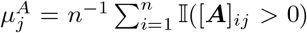. This typically results in a set of admissible candidates with a unique possibility for each column of 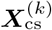 indexed by *J*, regardless of the cardinality of *J*. (If *J* = *p*^(*k*)^, then the resulting set typically reduces to 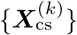 itself.)

##### 3. When only the local Gram matrix, parameter estimates and their standard errors are available to the response-node

We now turn our attention to the solution space of candidate matrices ***A*** when a subset *J*_sd_ *⊆* {1, …, *p*^(*k*)^}of standard error estimates 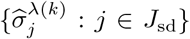 is disclosed. We focus on the case where these standard error estimates are released alongside the corresponding coefficient estimates, that is, if *J* corresponds to the subset of parameter estimates 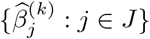 that are disclosed, then *J*_sd_ *⊆ J*.

Recall the expression of 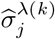 for *j* ∈ {1, …, *p*^(*k*)^}, given by

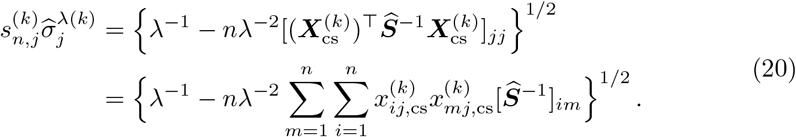

Since the values 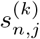 are not available to the response-node, but the coefficients 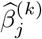 are known for *j* ∈ *J*_sd_, it follows from (18) that any candidate matrix 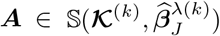 implicitly determines a corresponding candidate value for 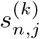, given by 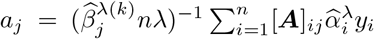. Consequently, reverse-engineering 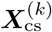 when the values 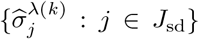 are disclosed requires the response-node to search for 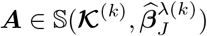 that satisfies, for all *j* ∈ *J*_sd_,

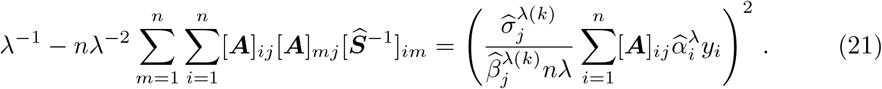

Letting 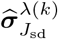 to denote the vector with entries 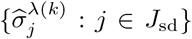, using the fact that any 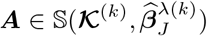 satisfies 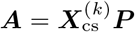 for some orthogonal matrix ***P***, the corresponding solution space is given by

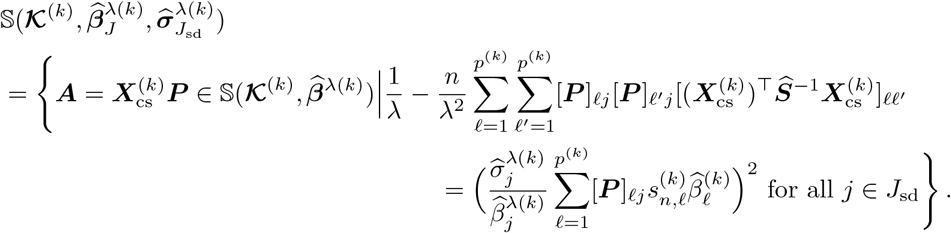

The next proposition can be viewed as an extension of Proposition 2 for the solution space when standard error estimates are disclosed. It can be proved using arguments similar to those employed in the proof of Proposition 2.

###### Proposition 3.

*Assume that p*^(*k*)^ *≥* 4, *and that* 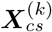 *has full column rank. Moreover, for* 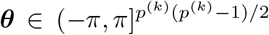, *let* ***P*** *and* ***g***(***θ***) *be defined as in Proposition 2*. *Consider* 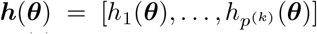 *and* 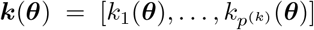, *where, for j* ∈ {1, …, *p*^(*k*)^},

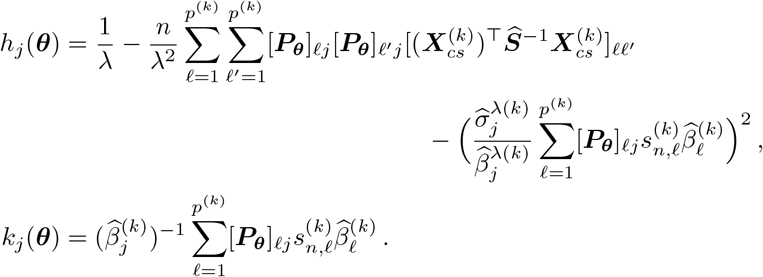

*Then, for* 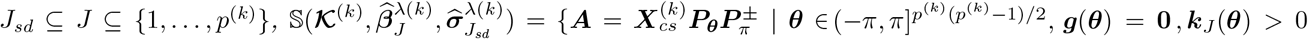, *and* 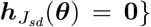. *Moreover, if* |*J*_*sd*_| *≤* (*p*^(*k*)^ − 1)(*p*^(*k*)^ − 2)*/*2 − 1, *the cardinality of the set* 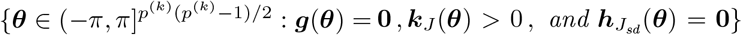 *is infinite, which implies that the cardinality of the set* 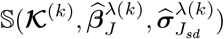 *is also infinite*.

##### 4. When only the local Gram matrix and two-sided p-values are available to the response-node

We now study the solution space of candidate matrices ***A*** when a subset *J ⊆* {1, …, *p*^(*k*)^} of two-sided p-values 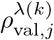 are available to the response-node, in addition to the local Gram matrix **𝒦**^(*k*)^. Given the one-to-one relationship between 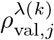 and 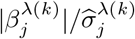, and using equations (18) and (20), for a candidate matrix ***A*** for 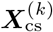 to be such that the disclosed quantities **𝒦**^(*k*)^ and 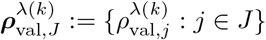 could have been equivalently computed from either ***A*** or 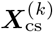, the response-node must identify a matrix ***A*** such that

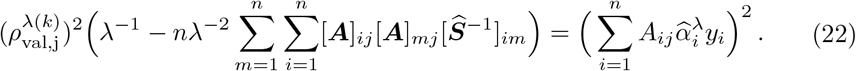

The latter equation is a re-expression of Equation (21) using 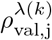 in place of 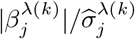. The corresponding solution space is given by

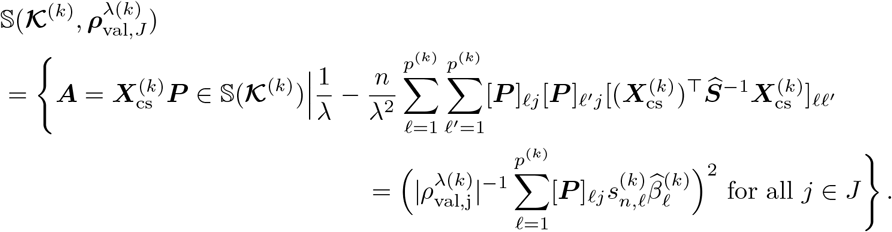

The expression of this latter solution space differs from that of 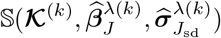 presented above. Specifically, since 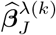—and in particular, its sign—is not available to the response-node, candidate matrices belonging to 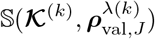, are not required to satisfy 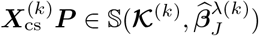. As a result, candidate matrices of the form 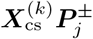 are always admissible elements of 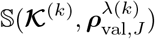.

##### 5. When only parameter estimates and their standard errors are available to the response-node, without the local Gram matrix and quantities derived from it

Recall from Methods 4.2.3 that 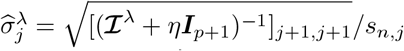, where *η* = *λ/n*, and where from Supplementary Methods 2.2, ***ℐ***^*λ*^ can be expressed as

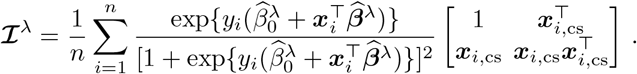

Also recall that 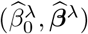 solve the maximization problem defined in (2) in the manuscript, which therefore implies that 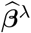 satisfies

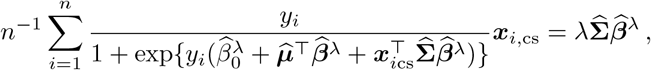

where we have introduced

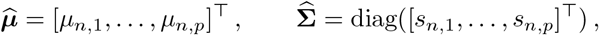

see Supplementary Methods 1 for details.

Suppose, without loss of generality, that the response-node also holds covariate data and is labeled as covariate-node *k* = 1, and consider the setting where only two nodes participate in the analysis: the response-node (also acting as a covariate-node) and a single additional covariate-node. The extension to scenarios involving more than two covariate-nodes follows analogously.

To analyze the privacy risk entailed when the set 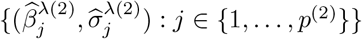 of parameter estimates and their associated standard errors—together with the response vector ***y*** and the covariate data it holds—is the only information available to the response-node, we adopt a deliberately more adverse scenario: for the purposes of this analysis only, we assume that, in addition to the 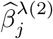 ‘s, the response-node has access to the full matrix **𝒥**^*λ*^ defined as

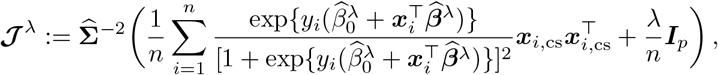

rather than solely to the 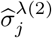’s, which satisfy 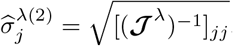. This conservative assumption simplifies the mathematical derivations and yields an upper bound on the potential privacy risk.

When at least one continuous covariate is held by a covariate-node located outside the response-node, then candidate matrices for 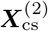 are column-centered and scaled matrix 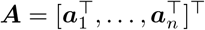 satisfying

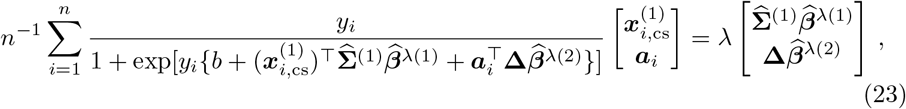

and

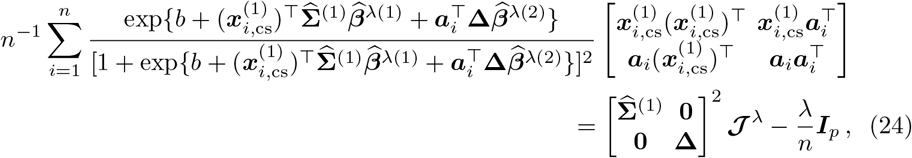

With 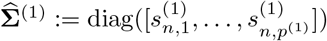, where 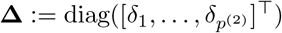 is the vector of unknown standard deviations, and *b* denotes the unknown intercept.

Assume temporarily that the only unknowns in the system of equations above are those associated with the (or one of the, if multiple exist) continuous variables held by covariate-node *k* = 2, and that all other entries of ***A*** are fixed and equal to those of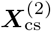. In this case, the system comprises *n* + 2 real-valued unknowns: the *n* entries of the continuous variable in ***A***, one candidate intercept *b*, and one candidate standard deviation corresponding to the continuous variable in **Δ**. On the other hand, the system imposes *p* + *p*(*p* + 1)*/*2 equality constraints: *p* equations from the first-order optimality condition in (23), and *p*(*p* + 1)*/*2 from the symmetry-reduced second-order condition in in (24) (since **𝒥**^*λ*^ is symmetric, the *p*(*p* − 1)*/*2 off-diagonal constraints are not independent). In addition, two further constraints are imposed to ensure that the column of ***A*** associated with the continuous variable is centered and scaled (i.e., has mean zero and variance one). Thus, the total number of equations is *p* + *p*(*p* + 1)*/*2 + 2, while the number of unknowns remains *n* + 2.

When the inequality *p* + *p*(*p* + 1)*/*2 *< n* holds, the number of unknowns exceeds the number of independent constraints. Since a solution lying in the interior of the feasible set exists (i.e., the configuration defined by 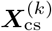), the constraint set defines a smooth manifold of positive dimension in a neighborhood of that point. Therefore, the system admits infinitely many solutions when *p*+*p*(*p*+1)*/*2 *< n*, provided that the component of 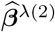 associated with the continuous variable is nonzero. This argument can be repeated for each continuous covariate held by covariate-node *k* = 2, showing that each associated column admits infinitely many admissible candidate configurations.

Now assume that, in addition to a continuous covariate, 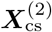 also includes a centered and scaled binary covariate. Consider the case where the entries of ***A*** corresponding to this binary covariate match those of 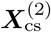, except for two entries—one originally positive and one originally negative in 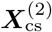—whose signs are flipped to preserve the column’s mean and variance (such a pair always exists under the assumption that ***X***_cs_ is not colinear with **1**_*n*_). As above, assume that all other entries of ***A*** are equal to those of 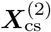, except for those in the column corresponding to the (or one of the, if multiple exist) continuous covariate. Now interpret the equations in (23) and (24) as defining a system in which the unknowns are the entries of ***A*** associated with the continuous covariate, its associated standard error, and the unknown intercept. Because the constraints vary smoothly with respect to the continuous covariate entries of ***A***, and provided that the vector ***y*** contains at least two entries equal to 1 and at least two equal to −1, the system admits at least one solution whenever *n* is larger than *p* + *p*(*p* + 1)*/*2. This is because, as the constraints behave smoothly, small changes in the continuous values can compensate for small mismatches elsewhere, allowing the system to adjust without violating the structure required by the observed quantities.

It follows that, for any entry in a column corresponding to a binary covariate in 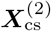, there exists an admissible candidate matrix in which that entry takes a different value.

###### Summary of scenarios 1 to 4 above where the response-node has access to the Gram matrix

In Table 4, we provide a summary of the solution sets for admissible candidates of 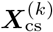 at the response-node for continuous and binary covariates at site *k*, when covariate-node *k* is not located at the response-node. Details are provided for settings in which the number of covariates and the quantities disclosed vary. For conciseness, in scenarios where standard errors are disclosed, we assume that the set of indices for the disclosed standard errors is the same as that for the disclosed parameter estimates, i.e., *J*_sd_ = *J*. We report only a subset of all admissible candidate matrices. Specifically, the reported subset is constructed so that, for each column, there exist at least two matrices in the subset that differ in at least one entry of that column. While additional admissible candidate matrices exist—including ones that allow more variation in columns not associated with the shared estimates (i.e., those with indices not in *J*)—they are omitted, as they do not yield additional information relevant to the assessment of privacy risk (see Methods 4.3.3).

**Table 4.**
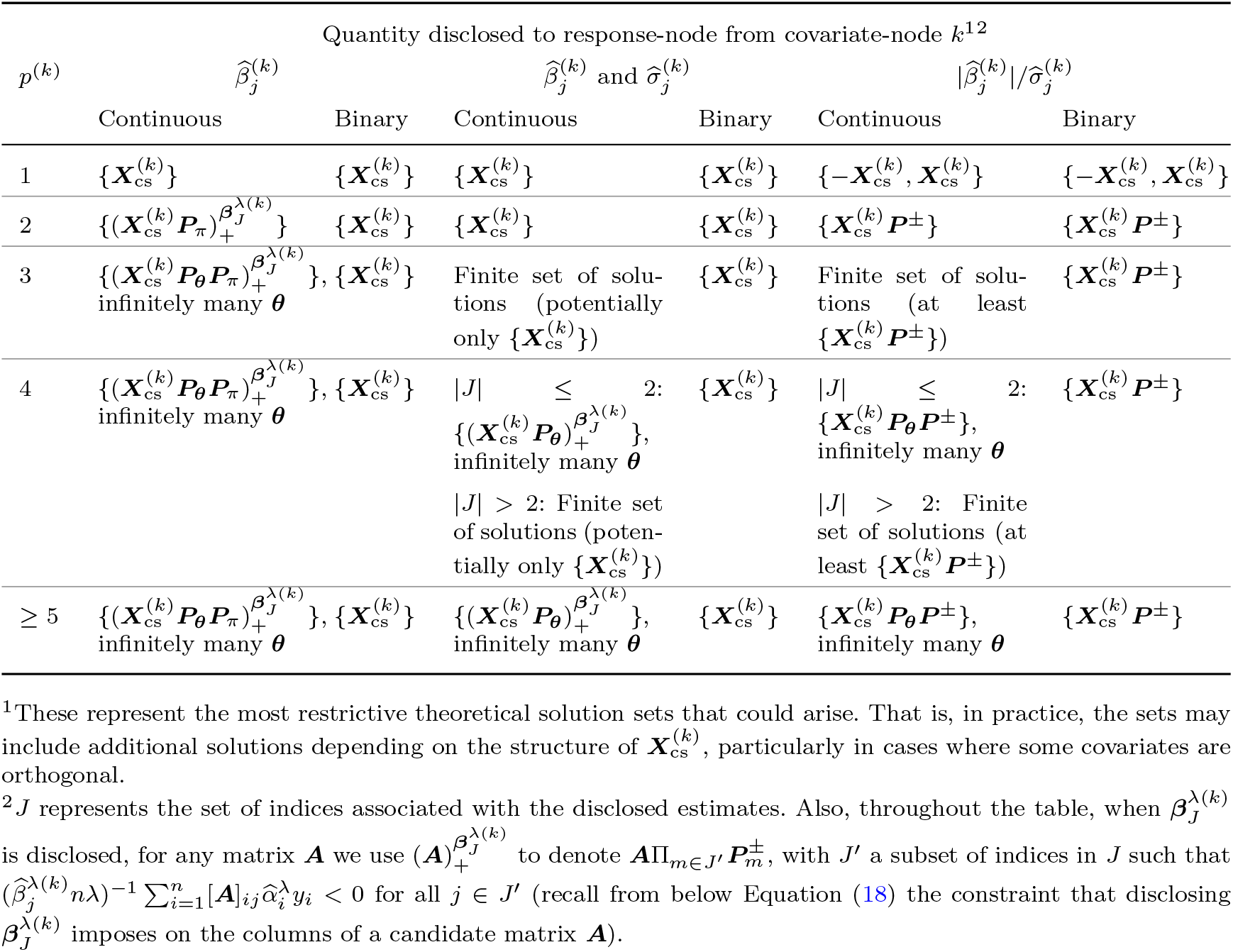
Solution set for admissible candidates of 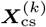 at the response-node depending on the quantities disclosed with index *j* ∈ *J*.

#### 4.3.2 Methodology to assess the privacy-preserving properties of quantities shared by the response-node

Consider a given covariate-node *k* ∈ {1, …, *K*}, assumed to be adversarial, that is located outside of the response-node, and recall that the response-node shares with the latter the vector 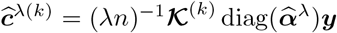 and the matrix ***Ŝ***^−1^ + **𝒩** ^(*k*)^(**𝒩** ^(*k*)^)^⊤^, respectively, where **𝒩**^(*k*)^ is in the null-space of 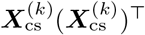 (and of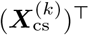). In the following analysis, we examine covariate-node k’s ability to re-identify any entry of *y* based on **ĉ**^*λ*(*k*)^ and ***Ŝ***^−1^ + **𝒩** ^(*k*)^(**𝒩** ^(*k*)^)^⊤^.

We assume that at least two covariate-nodes participate in the analysis (including the possibility that one is co-located with the response-node), that ***X***^(*k*)^ has full-column rank, that *n > p*^(*k*)^ and that at least one continuous covariate is held outside of covariate-node *k*.

First, ***Ŝ***^−1^ cannot be retrieve from ***Ŝ***^−1^ + **𝒩**^(*k*)^(**𝒩** ^(*k*)^)^⊤^ at covariate-node k, since

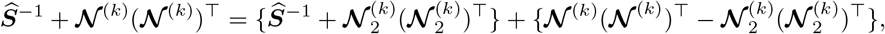

where **𝒩** _2_ is any other matrix selected in the null-space of (***X***^(*k*)^)^⊤^. Since 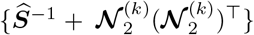 is symmetric and invertible (because ***Ŝ*** is symmetric positive-definite (as defined in (13)), and the sum of a positive-definite and a positive semi-definite matrix is itself positive-definite) while the matrix 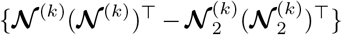 lies in the null space of 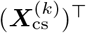, the values of ***Ŝ***^−1^ cannot be recovered.

We now examine the ability of covariate-node *k* to retrieve the response-vector *y* from 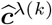. To do this, and recalling that covariate-node *k* has access to 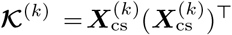 and to *λ*, let

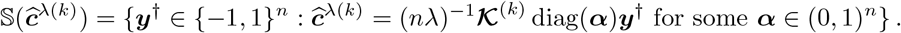

Since we have assumed that at least one continuous covariate is held outside of covariate-node *k*, ***α*** can be treated as a vector with real-valued entries. Moreo-ever, since we have assumed that ***X***^(*k*)^ has full-column rank, then so does ***X***^(*k*)^ and as *n > p*^(*k*)^, the null-space of **𝒦**^(*k*)^ has dimension *n* − *p*^(*k*)^ *>* 0. Letting ***W*** denote an *n ×* (*n* − *p*^(*k*)^) matrix of linearly independent columns spanning the null-space of **𝒦** ^(*k*)^, any solution ***x***_0_ satisfying **ĉ** ^*λ*(*k*)^ = (*nλ*)^−1^ **𝒦**^(*k*)^ ***x***_0_ can be expressed as 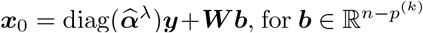,, the solution space 𝕊(***ĉ***^*λ*(*k*)^) can be re-expressed as

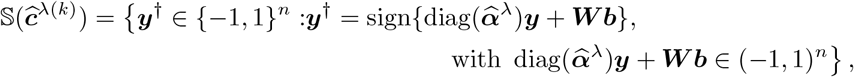

where, in the above equation, the function sign(·), when applied to a vector, is understood component-wise: it returns −1 for each negative entry and 1 for each positive entry. To derive this expression, we also used the fact that any ***y***^†^ ∈ {−1, 1}^*n*^ satisfying **ĉ** ^*λ*(*k*)^ = (*nλ*)^−1^ **𝒦**^(*k*)^ diag(***α***)***y***^†^ for some ***α*** ∈ (0, 1) satisfies ***y*** ^†^= sign(diag(***α***)***y***^†^).

To ensure sharing ***c***^(*k*)^ achieves Privacy-Level II, one needs to verify that, for 1 *≤* 𝕀 *≤ n*, there exists a vector ***y***^†(*i*)^ ∈ {−1, 1}^*n*^ with 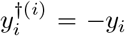, such that ***y***^†(*i*)^ ∈ 𝕊 (**ĉ**)^*λ*(*k*)^. This can be done using linear programming algorithm designed to find feasible solutions under linear inequality constraints.

It remains to address the case where a covariate-node is co-located with the response-node, such that an adversarial covariate-node could attempt to infer the covariate data held by the response-node. The re-identification risk in this setting is conservatively assessed using the privacy analysis from the scenario *5. When only parameter estimates and their standard errors are available to the response-node, without access to the local Gram matrix or any quantities derived from it*, as discussed above. In this case, the risk of re-identifying the response-node’s covariate data is lower than that of re-identifying a covariate-node’s data by the response-node, since the adversarial covariate-node does not have access to ***y***.

#### 4.3.3 Summary of the assessments for the privacy analysis: connecting Methods 4.3.1 and 4.3.2 to Results 2.3

Recall from Results 2.3 that given a set of quantities that could be used to attempt reconstruction of 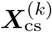, for each each column, we assess its number of *admissible candidate configurations*—defined as the number of admissible candidate matrices in which the values for that column differ in at least one entry from those in other admissible datasets, and that we defined therein three *levels of indirection* based on the smallest such number across all variables of the considered dataset: **Level II-(i)** if it is equal to 2; **Level II-(ii)** if that number is a finite number greater than 2; **Level II-(iii)** if it is infinite.

##### Alternative definitions of the indirection levels

Alternative definitions could have been based on the total number of admissible candidate datasets, or on the minimum number—over all entries in the dataset—of candidate datasets that differ at that entry. Our choice to define the levels of indirection based on the minimal number of admissible candidate configurations across all variables is motivated by two considerations. First, for a given dataset, depending on the quantities disclosed, it may occur that some variables admit infinitely many admissible configurations, while others admit only a finite number. In such cases, relying on the total number of admissible candidate datasets can be misleading. Second, for binary variables, the minimum number—over all entries—of candidate datasets that differ at a given entry is bounded by 2, since each entry can only take one of two possible values. In contrast, the number of admissible candidate configurations for a variable can be substantially larger, offering arguably a more meaningful characterization of uncertainty.

##### Derivation of the decision trees

Using the retained definitions of indirection levels, the decision trees presented in Results 2.3.1 were derived from the theoretical arguments established in the cases *1. When only the local Gram matrix is available to the response-node* and *5. When only parameter estimates and their standard errors are available to the response-node, without the local Gram matrix and quantities derived from it* treated above. The decision trees presented in Results 2.3.2 were derived from the theoretical arguments established in the cases *1. When only the local Gram matrix is available to the response-node, 2. When only the local Gram matrix and parameter estimates are available to the response-node, 3. When only the local Gram matrix, parameter estimates and their standard errors are available to the response-node* and *4. When only the local Gram matrix and two-sided p-values are available to the response-node*. The decision trees were derived to qualitatively reflect the admissible candidates described in Tables 3 and 4. In producing the trees, we used the assumption that the scaled covariate values of each individual *i* ∈ {1, …, *n*} that are stored at node *k* are all different, that is, that 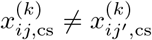 for *j*≠ *j*^*′*^. This assumption ensures that any column-permuted version of 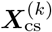 yields a candidate configuration that differs from the original in every entry. This condition is not restrictive in practice, as it holds with probability one for continuous covariates, and also holds for binary covariates whenever the sample means of the variables differ—which is nearly always observed in real-world data.

##### General considerations for the binary vs continuous covariate cases

The re-identification risks differ for continuous and binary covariates. In the binary case, each admissible candidate matrix for 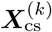 uniquely determines candidate values for the covariate means, since the average of the positive entries in each centered and scaled column equals the mean of the original binary covariate. Generally speaking, given the deterministic relationship between the mean and standard deviation for binary variables, ***X***^(*k*)^ becomes fully recoverable from **𝒦**^(*k*)^ if the covariate means are known (unless e.g. two columns in ***X***^(*k*)^ have exactly the same mean). With continuous covariates, the relationship between the mean and standard deviation is sample-dependent and therefore unknown, resulting in these parameters treated as free and not functionally related, making ***X***^(*k*)^ not recoverable from **𝒦**^(*k*)^ even when the covariate means are known. However, we note that every candidate for a column in 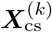 is associated with a fixed standard deviation when the column-associated exact parameter estimate is disclosed. While knowing the true mean and the fixed (but not true) standard deviation allows the response-node to retrieve the uncentered and unscaled version of every candidate, there still exists an infinite number of such candidates for cases that had an infinite number of candidates in Table 4. When only p-values are disclosed, if the mean is additionally disclosed, sign-flipped versions remain admissible candidates even for settings for which there was not an infinite number of candidates.

##### Covariate-nodes containing only binary covariates

In cases where all covariate-nodes—except possibly the one located at the response-node—contain only binary covariates, Privacy Level II may not be attained. Due to the binary nature of the covariates unknown to the response-node, it could theoretically enumerate all possible datasets formed by every combination of 0s and 1s for each entry. It could then identify those consistent with the quantities it possesses (i.e., parameter estimates or standard errors) by fitting the logistic regression model in (2) to each candidate dataset using the response vector it holds and comparing the resulting estimates to the disclosed values. In such cases, Privacy Level II may fail to hold, as it is possible that no two distinct datasets are consistent with the quantities available to the response-node. However, beyond the uncertainty regarding the uniqueness of the admissible candidate dataset, this procedure is computationally demanding.

##### Privacy analysis results for the response-node

The results were directly derived from the analysis in Methods 4.3.2. The empirical criterion to assess whether sharing ***c***^(*k*)^ achieves Privacy-Level II is detailed in Supplementary Methods 4.4.

### 4.4 Methodology for real health data applications

We applied the VALORIS algorithm detailed in Algorithm 1 to two cases involving real heath data and performed analyses in R (version 4.4.1) [24]. The parameter *λ* was set to *λ* = min(10^−4^, *n*^−1^), and we took *ϵ* = *n*^−1^𝕀(*n ≤* 10000)+5*n*^−1^𝕀(*n >* 10000). The applications where selected to include both numerical and binary covariates, to represent cases with sample size respectively lower than 100 and higher than 10000, and to allow for varying number of covariate-nodes. Complete-case analyses were conducted, whereby individuals with missing values were excluded.

When presented, Wald-type 1 − *α* CIs for the *j*th covariate were computed using 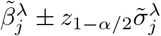. All results were rounded using three decimals.

During the analysis, the empirical criterion regarding the privacy-preserving properties for the response-vector was verified (see Supplementary Methods 4.4). The criterion was met in both applications, such that Privacy Level II was achieved for the response-node data in all presented examples.

## Data availability

The synthetic dataset to test the implementation in R is publicly available on GitHub [21]. The MIMIC-IV dataset is available online under certain conditions [19], including completion of mandatory training. The de-identified clinical dataset from the Necker-Enfants Malades Hospital used only as an example in this study is not publicly available following institutional officials recommendations [20].

## Code availability

The code used in this study is available on GitHub: https://github.com/OpenLHS/Distribanalysis/tree/main/Vertically_distributedanalysis/logistic_regression_nonpenalized

## Supplementary information

See Supplementary Information file for Supplementary Tables, Supplementary Notes 1 and Supplementary Methods 1-4.

## Acknowledgements

This work was supported by the Natural Sciences and Engineering Research Council of Canada – Discovery Grant; the Health Data Research Network Canada, an initiative funded by the Canadian Institutes of Health Research; and the Chaire de recherche en informatique de la santé de l’Université de Sherbrooke. Marie-Pier Domingue received a scholarship from the Natural Sciences and Engineering Research Council of Canada. This study was performed in the context of the C’IL-LICO project, a research initiative coordinated by the Imagine Institute that was supported by state funding from The French National Research Agency (ANR) (Reference: ANR-17-RHUS-0002). This work was supported by State funding from the Agence Nationale de la Recherche under “Investissements d’avenir” program (ANR-10-IAHU-01). We thank Xiaomeng Wang for preprocessing clinical data used in the application of the method.

## Conflicts of Interest

None declared.

## Supplementary Information

### Supplementary Tables 1

**Fig. S1.**
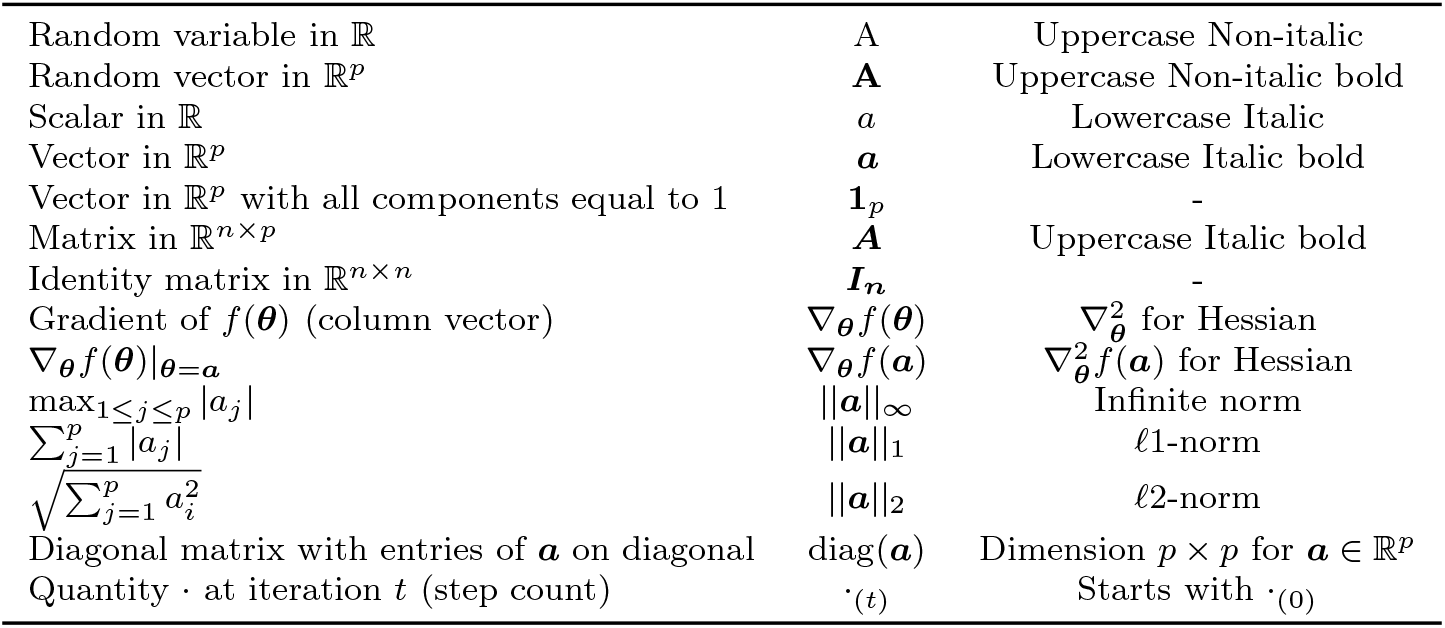
Glossary for general notation conventions

**Fig. S2.**
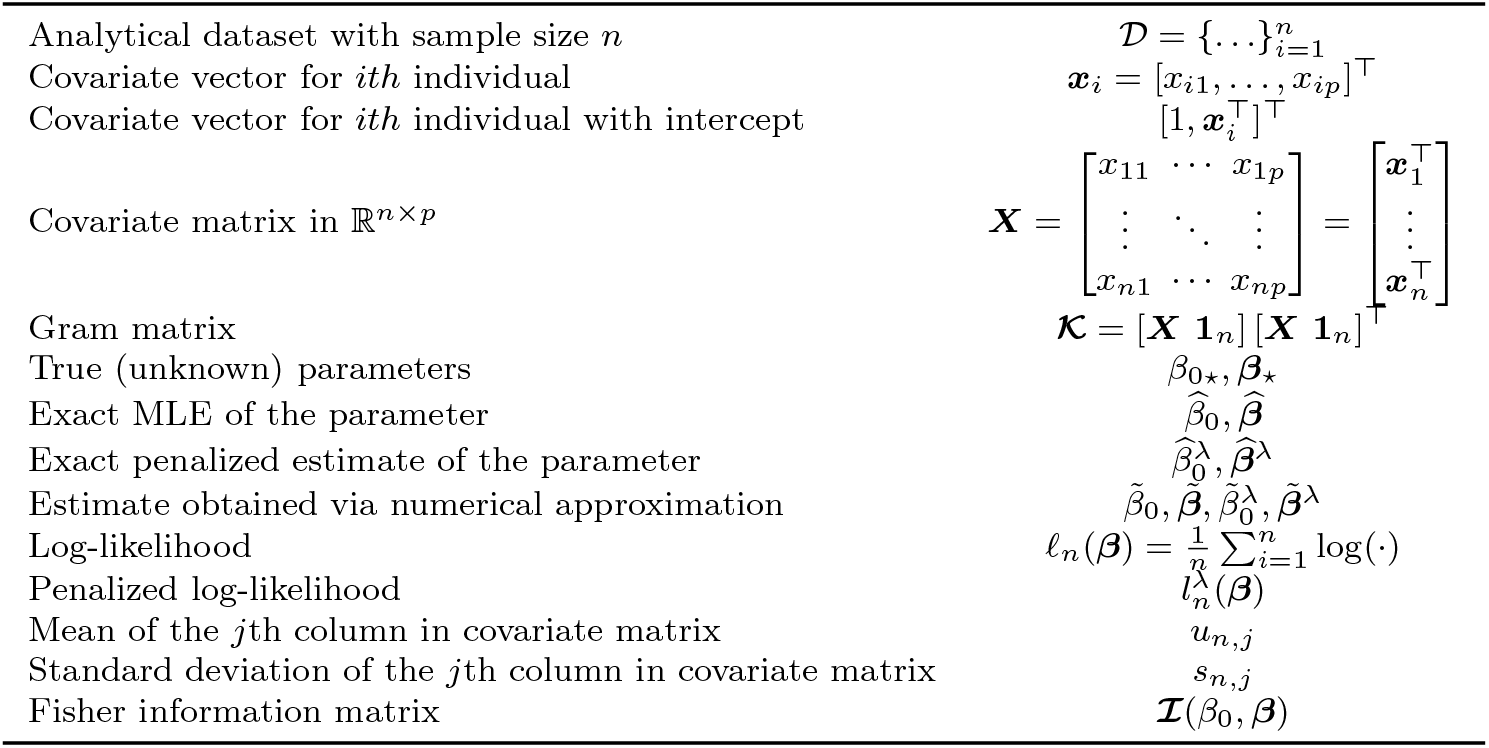
Glossary for quantities that pertain to the regression settings

**Fig. S3.**
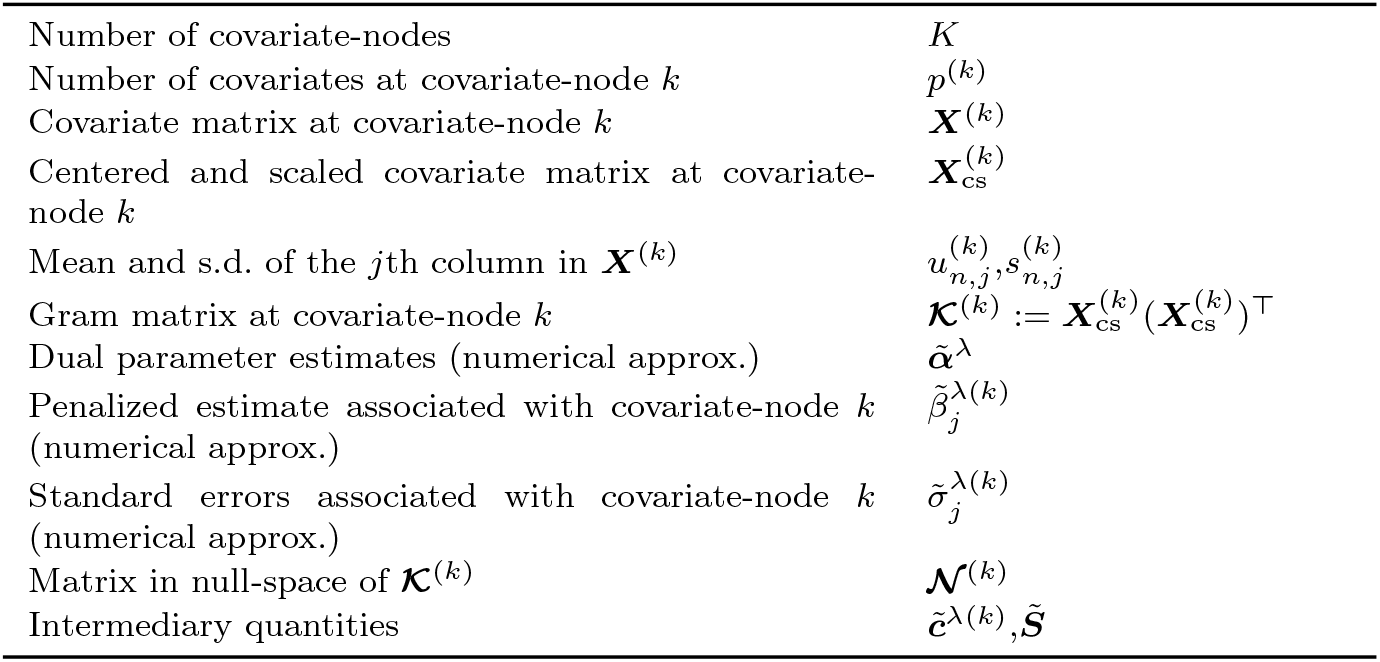
Glossary for quantities specific to the vertical setting

### Supplementary Notes 1

The code for the implementation of the algorithm using R is available at: https://github.com/OpenLHS/Distrib_analysis/tree/main/Vertically_distributed_analysis/logistic_regression_nonpenalized. It includes an automated example with simulated data. The folder also includes a basic implementation of the tool that supports the privacy assessment to verify if an infinite number of solutions exists in some settings.

### Supplementary Methods 1 Theoretical derivations for the optimization problem

In the followings, for any (*β*_0_, ***β***) let

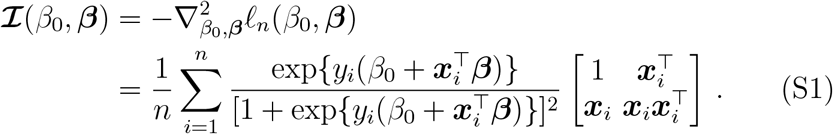

In this notation, 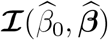 corresponds to the observed Fisher information matrix introduced in Equation (10) in the manuscript.

The following lemma establishes that the unique solution to the ridge-penalized log-likelihood maximization problem for logistic regression can be obtained by solving its dual formulation, which is a minimization problem over a compact search space. This result implies that, for a given sample and a fixed λ, the solution to the dual minimization problem cannot lie arbitrarily close to the boundary of the domain (0, 1)^*n*^.

#### Lemma S1.

*For any (y*_1_, ***x***_1_), …, (*y*_*n*_, ***x***_*n*_) ∈ {−1, 1} ×ℝ ^*p*^, *the unique maximizer* 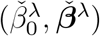 *of the maximization problem*

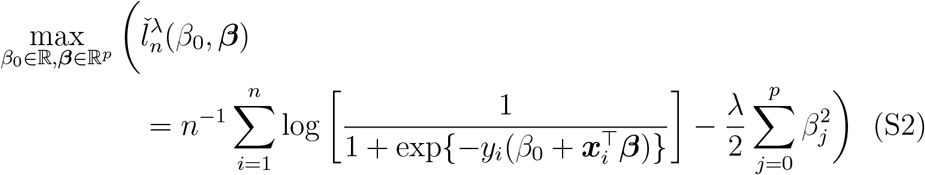

*satisfies*

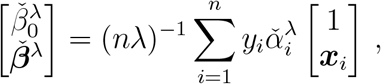

*where* 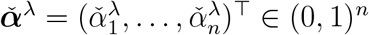 *is the unique solution to the following minimization problem:*

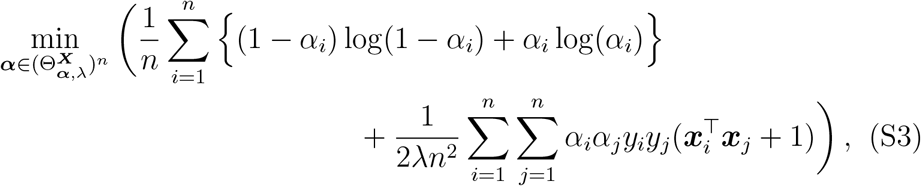

*with* 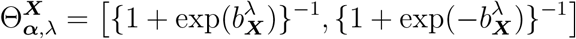,

*where* 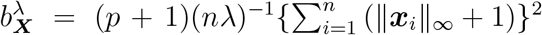. *Moreover*, 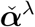 *is the unique stationary point of the objective function in* (S3), *and the set* 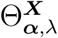 *can be replaced by* (0, 1).

*Proof of Lemma S1*. We begin by showing that the search space ℝ × ℝ^*p*^ in the maximization program on the first line at (S2) can be replaced by a suitably chosen compact set. To this end, we first note that the function 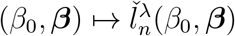 is strongly concave. This follows upon observing that as 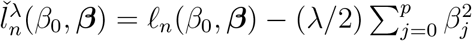, from (S1) we have

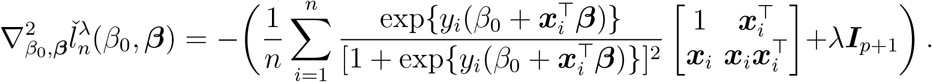

Since the first term inside the parentheses is a weighted sum of positive semi-definite matrices with strictly positive weights, and the second term is a diagonal matrix with strictly positive entries, their sum is positive definite. This implies that the Hessian 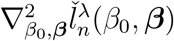 is negative definite, and therefore, the penalized log-likelihood function 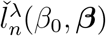 is strongly concave.

As 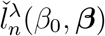 is strongly concave, its maximum is unique and is achieved at the point 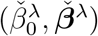 that satisfies

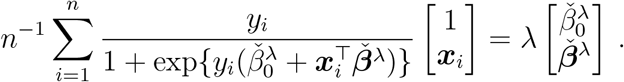

By the triangle inequality, the latter equation implies

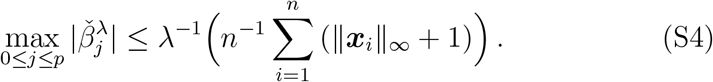

Therefore, letting 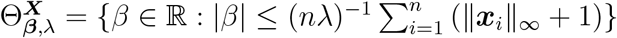, it holds for any ***x***_1_, …, ***x***_n_ ∈ ℝ^*p*^ that

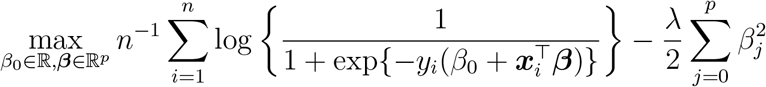

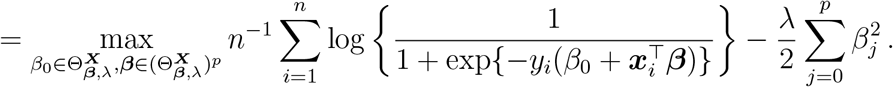

Next, we show that

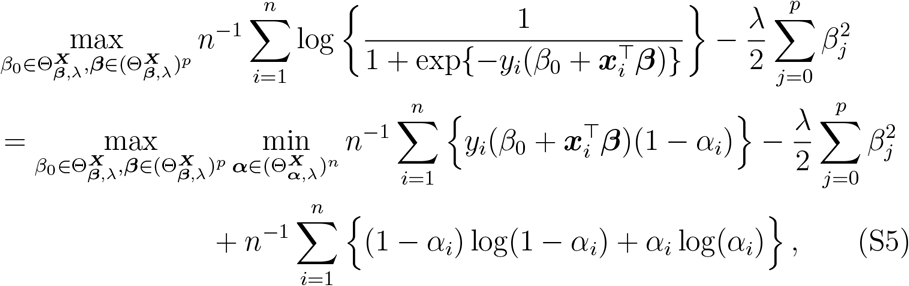

with 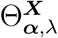 as in (S3).

To establish this result, we begin by noting that for any *x* ∈ [*a, b*], we have

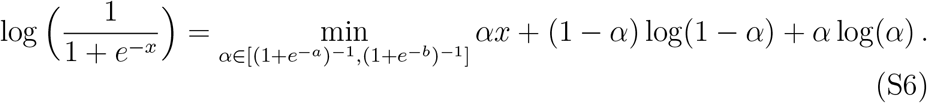

To see why (S6) holds, it suffices to first verify that the function *α* ↦ *αx* + (1 −*α*) log(1 − *α*) + *α* log(*α*) attains its minimum at *α* = (1 + *e*^*x*^)^*−*1^. Substituting *α* = (1 + *e*^*x*^)^*−*1^, we obtain

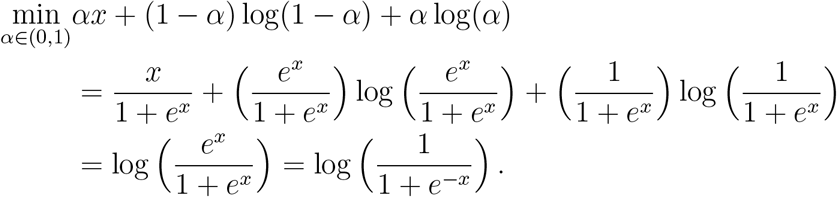

From this, (S6) follows directly from the fact that *x* ∈ [*a, b*].

Since

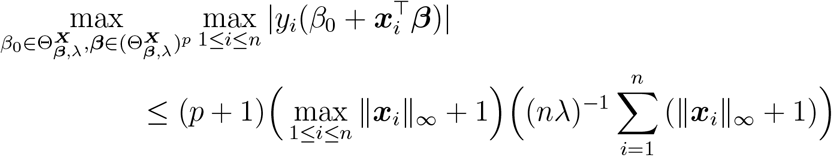

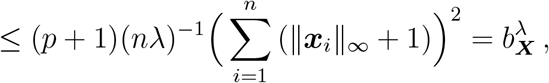

where 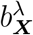 is defined in the statement of the lemma, the proof that (S5) holds follows from (S6), which ensures that

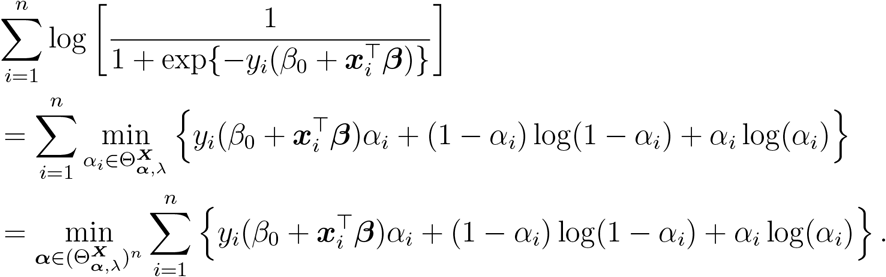

To conclude the proof of the lemma, it remains to show that in the optimization problem given on the second line of (S5), we can interchange the maximum and minimum operations and then solve the inner maximization problem explicitly.

To prove that we can swap the max and the min, we apply Sion’s minimax theorem [1, 2]. To justify the application of this theorem and conclude that the order of the minimum and maximum operators can be interchanged, we must verify that the function

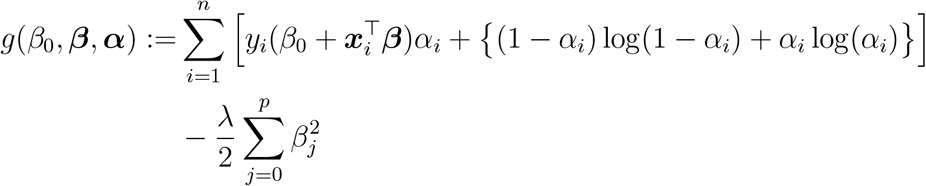

is such that for any fixed (*β*_0_, ***β***), ***α*** ↦ g(*β*_0_, ***β, α***) is convex, and for any fixed ***α***, (*β*_0_, ***β***) ↦ g(*β*_0_, ***β, α***) is concave. Once these conditions are established, Sion’s theorem guarantees that the maximum and minimum operators can be interchanged.

The convexity of ***α*** ↦ g(*β*_0_, ***β, α***) follows from the fact 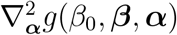 is a diagonal matrix, with diagonal entries given by the following, for *j* ∈ {1, …, *n*}:

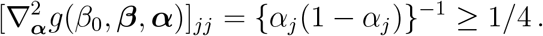

Since each diagonal entry is positive for all ***α*** ∈ (0, 1)^*n*^ and 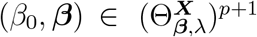, the Hessian is positive definite, which ensures that ***α*** → *g*(*β*_0_, ***β, α***) is convex in ***α*** for all (*β*_0_, ***β***).

The concavity of (*β*_0_, ***β***) → *g*(*β*_0_, ***β, α***) follows from the fact that 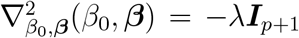, with ***I***_*p*+1_ the identity matrix of size (*p* + 1) *×* (*p* + 1). As the Hessian of *g*(*β*_0_, ***β, α***) with respect to (*β*_0_, ***β***) is a diagonal matrix with negative entries, it is therefore negative definite, which ensures that (*β*_0_, ***β***) → *g*(*β*_0_, ***β, α***) is concave for all (*β*_0_, ***β***) and ***α***.

Since we have just proved that for any fixed (*β*_0_, ***β***), ***α*** → *g*(*β*_0_, ***β, α***) is convex, and for any fixed ***α***, (*β*_0_, ***β***) → *g*(*β*_0_, ***β, α***) is concave, we can apply Sion’s minimax theorem, which, from (S5), ensures that

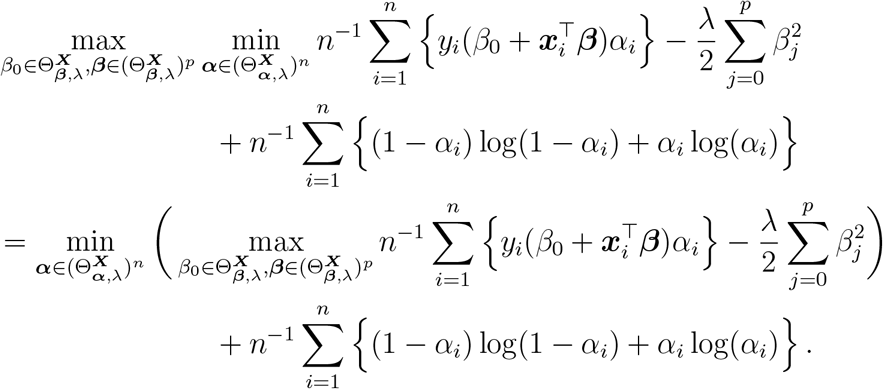

Now the inner maximization program can be solved exactly, since, for any 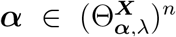, the maximum of 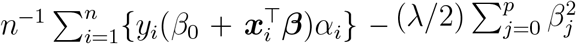 is achieved at

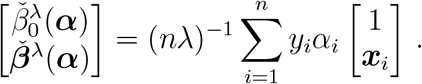

Since it can readily be verified that 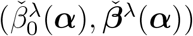 lies in the interior of 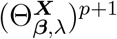 we therefore have

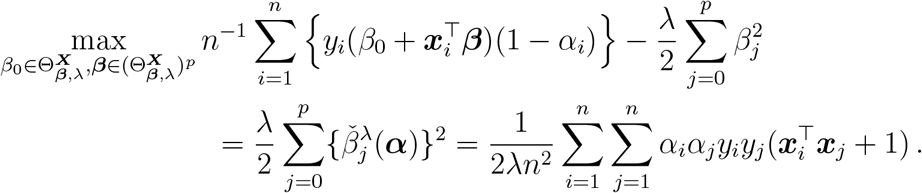

This concludes the proof of the lemma. □

Recall the maximization problem defined in (2) in the manuscript, namely,

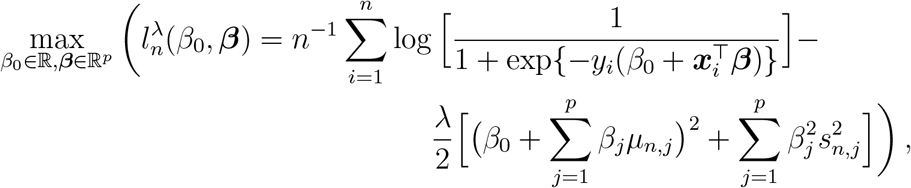

and recall from the manuscript that its solution is denoted by 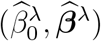. Also, recall the definition of *J*^*λ*^(***α***) in (3) in the manuscript, i.e., that

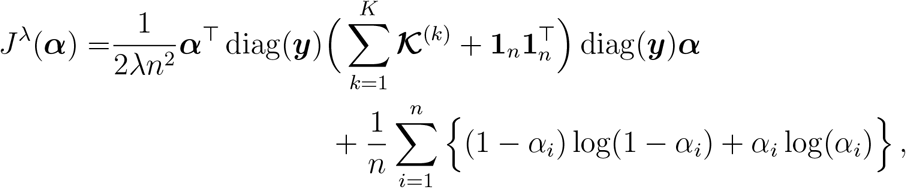

where 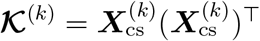.

The following proposition proves the assertion in Methods 4.2.2 that the solution 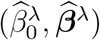 can be computed using Equation (11) in the manuscript.

#### Proposition S1.

>*Assume that s*_*n,j*_ *>* 0 *for all j* ∈ {1, *…, p*}. *Then*, 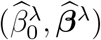 *satisfies*

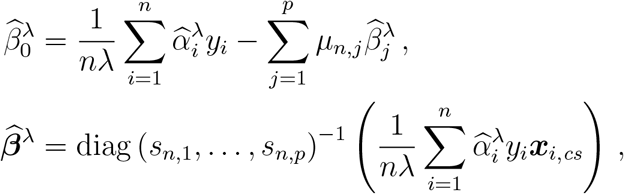

*where* 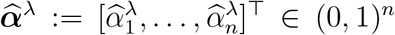 *is the unique minimizer of J*^*λ*^(***α***) *over* (0, 1)^*n*^.

*Proof*. Since, for all *i* ∈ {1, *…, n*} we have

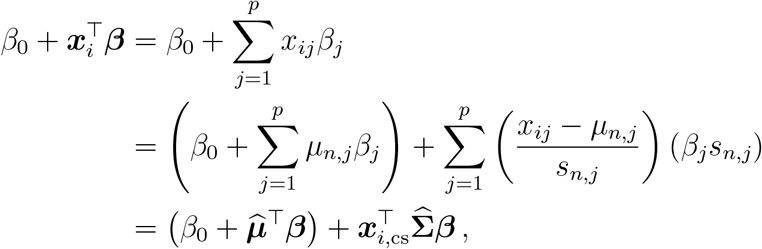

where we have introduced

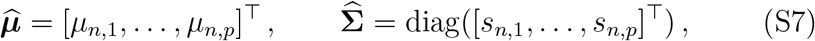

it follows upon adopting the re-parametrization 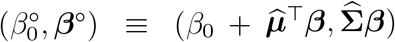 that

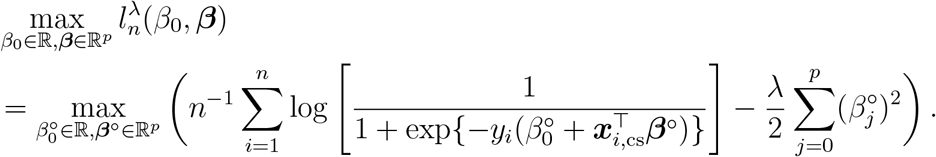

The maximization problem on the last line of the previous equation fits the framework of Lemma S1, which implies that its unique maximizer, denoted by 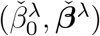, satisfies

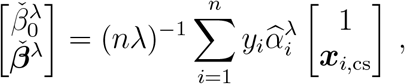

where 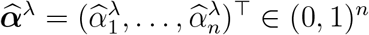 is the unique solution to the following minimization problem:

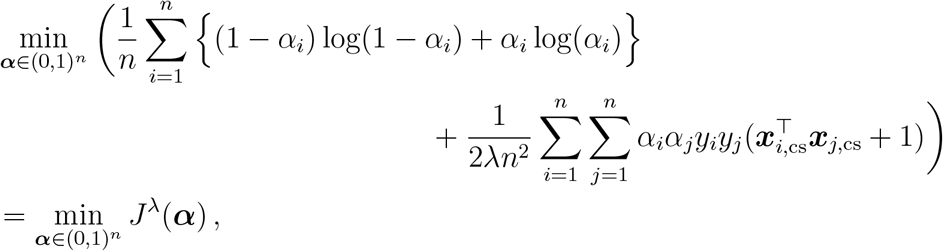

where, in applying Lemma S1, we replaced the set 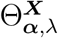 by (0, 1).

The proof of the proposition follows from the fact that since the bijective nature of the reparametrization implies

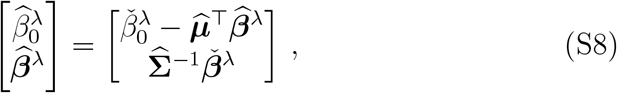

we have

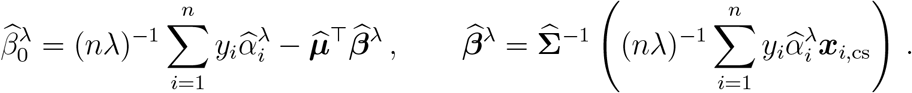

□

The following lemma establishes that, if the unpenalized maximum likelihood estimate 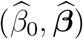 exists and is unique, then it is close to the penalized estimate 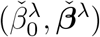 defined in Lemma S1 for sufficiently small *λ*. It is well known [3] that if the columns of the matrix ***X*** are linearly independent, and also linearly independent of the vector **1**_*n*_, then the Hessian 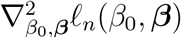 is strictly negative definite, implying that the log-likelihood function 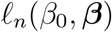 is strictly concave. In this case, if a maximizer exists for the problem 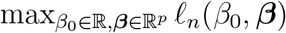 then it must be unique, and it must be a stationary point of 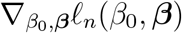. The existence of such a solution is guaranteed when the response vector ***y*** is not separable [4]. Specifically, ***y*** is said to be separable if there exists (*β*_0_, ***β***) such that 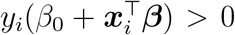 for all *i* ∈{1, *…, n*}. In the presence of separability, the log-likelihood function increases indefinitely, and a finite maximum likelihood estimate does not exist.

In what follows, for any positive definite matrix ***A***, let *ι*_min_(***A***) denote its smallest eigen value.

#### Lemma S2.

*Let* (*y*_1_, ***x***_1_), …, (*y*_*n*_, ***x***_*n*_) ∈{−1, 1} × ℝ^*p*^ *be such that the matrix* [**1**_*n*_, ***x***_1_, …, ***x***_*n*_]^*⊤*^ *has full column rank and such that* ***y*** *is not separable. Then, the unique solution* 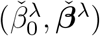 *to the maximization problem* 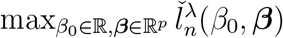, *with* 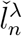 *as defined in Lemma S1, satisfies*

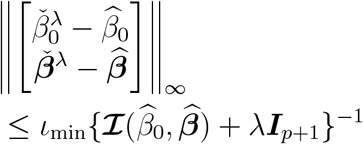

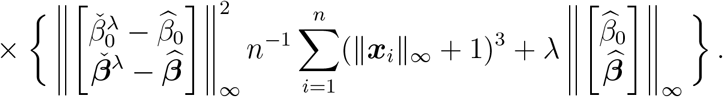

*Proof of Lemma S2*. Since 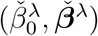 is a stationary point of 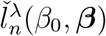, we have 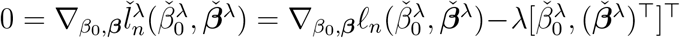, and therefore

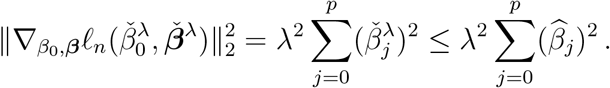

Since 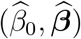 does not depend on *λ* and is finite, we conclude that as 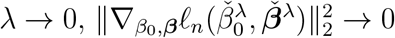, which implies that 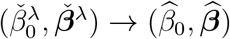 as *λ* → 0 since the maximum of *ℓ*_*n*_(*β*_0_, ***β***) is unique.

Now since 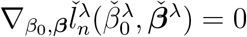 we have

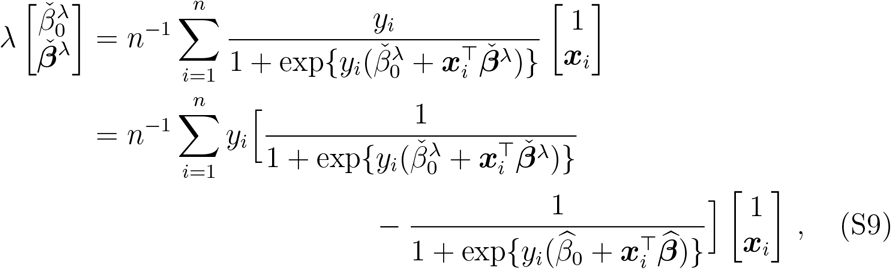

where, to obtain the second line, we used the fact that 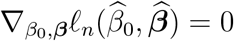.

As for any *x, y* ∈ ℝ a Taylor expansion of order two shows that

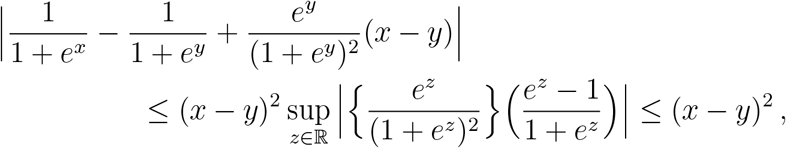

and since (S9) implies

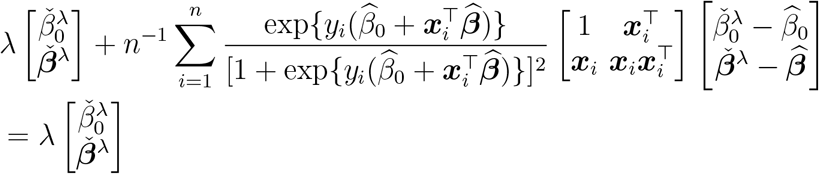

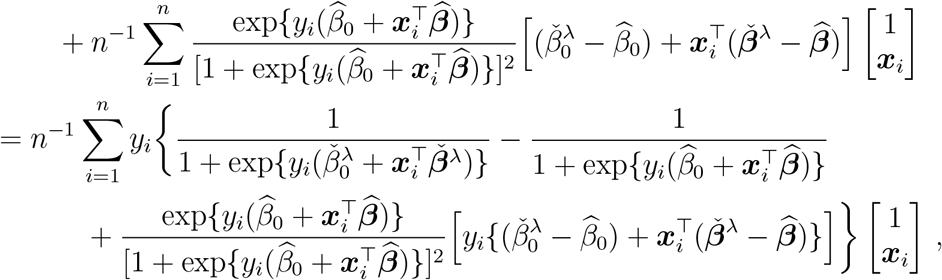

we conclude that

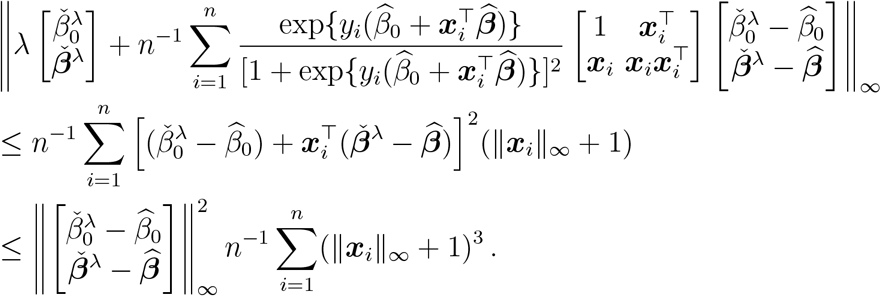

Recalling the definition of ℐ (*β*_0_, ***β***) above the statement of the lemma, by rearranging the terms and applying the triangle inequality, we obtain from the last equation that

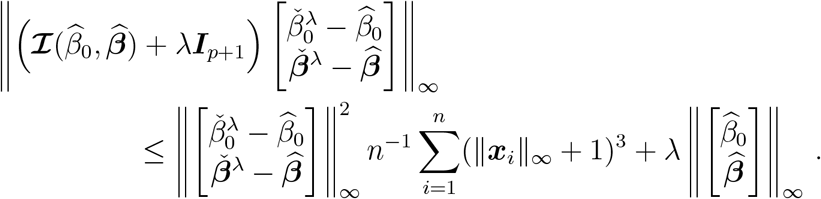

The result follows from the fact that for any positive matrix ***A***, ∥***Ax***∥ ≥ *ι*_min_(***A***)∥***x***∥ □

#### Remark S1.

>*Consider* 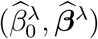, *and let* 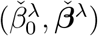 *be defined as in the proof of Proposition S1. Also, let* 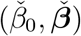 *and* 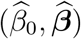 *be the maximum likelihood estimates that solves the log-likelihood maximization problem based on, respectively, the centered and scaled covariate data* ***x***_1,*cs*_, …, ***x***_*n,cs*_, *and the covariate data in their original scale* ***x***_1_, …, ***x***_*n*_. *Using arguments that are identical to the ones used in the proof of Proposition S1, it is straightforward to deduce that*

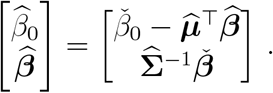

*Combining the last equation to* (S8) *implies*

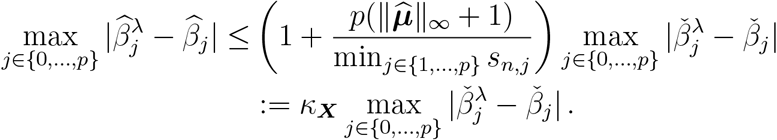

*Since* 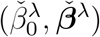 *and* 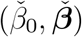 *fit the framework of Lemma S2, we have, under the lemma’s assumptions that*

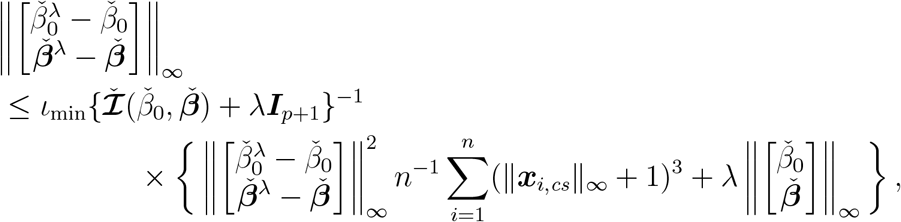

*with* 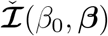 *a version of ℐ* (*β*_0_, ***β***) *(see* (S1)*) where the original data are replaced by the centered and scaled data, given by*

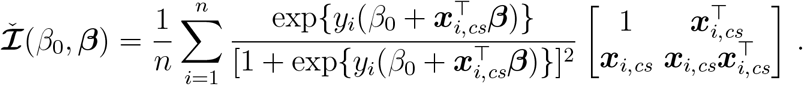

*Since the inequality* ∥***x***_*i,cs*_∥_*∞*_ *≤ κ*_***X***_∥***x***∥_*i*_ + *κ*_***X***_ *implies* 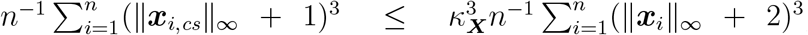, *and as* 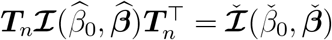, *with*

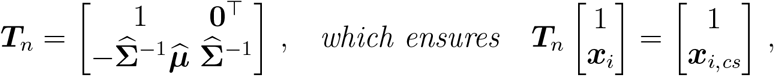

*then, we conclude that*

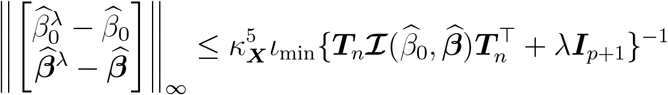

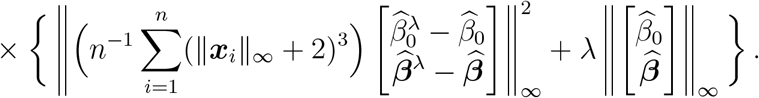

### Supplementary Methods 2 Auxiliary results related to the asymptotic normality of 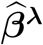 and computation of standard errors

#### 2.1 Asymptotic normality of 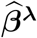 and consistency of standard error estimates

Recall from Results 2.1 that we assume a binary random variable *Y* ∈ {−1, 1} and a random vector of covariates **X** = [*X*_1_, *…, X*_*p*_]^*⊤*^ ∈ ℝ^*p*^ following a logistic regression model. In this model, there exists an unknown parameter vector *β*_0⋆_ ∈ R, ***β***_⋆_ ∈ ℝ^*p*^ such that

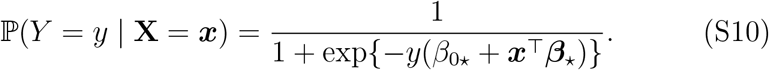

Let (Y_1_, **X**_1_), …, (Y_*n*_, **X**_*n*_) be i.i.d. random variables satisfying the model in (S10). Throughout this section, we use *ℓ*_*n*_(***β***) and 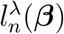, as defined in (9) and (2), respectively, where the (*y*_*i*_, ***x***_*i*_)’s in their definitions are replaced here by the random variables (Y_*i*_, **X**_*i*_). Specifically, we consider

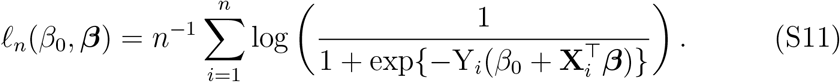

and

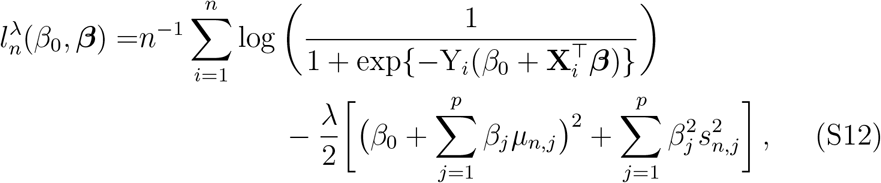

with 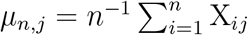 and 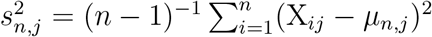.

Throughout this section, we also consider a version of 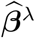 computed with the random variables (Y_1_, **X**_1_), …, (Y_*n*_, **X**_*n*_). That is, we define the estimator 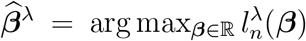, with 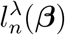 as in (S12) (recall from Lemma S1 that when *λ >* 0 the function 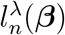 is strongly concave and has a unique maximizer).

We also consider a version of 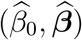 computed from the random variables (Y_1_, **X**_1_), …, (Y_*n*_, **X**_*n*_). That is, we define the maximum likelihood estimator as 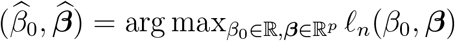. For sufficiently large *n*, this estimator exists with probability one, since for any finite 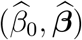, the response vector ***Y*** will be non-separable with probability one when *n* is large enough.

Likewise, we consider of version of ***ℐ*** (*β*_0_, ***β***) at (S1), where the (*y*_*i*_, ***x***_*i*_)’s are replaced here by the random variables (Y_*i*_, **X**_*i*_).

The following lemma establishes that, if 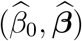 converges in probability to the true 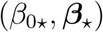, then 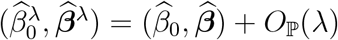

##### Lemma S3.

*Let* (Y_1_, **X**_1_), …, (Y_*n*_, **X**_*n*_) *be i*.*i*.*d. random variables satisfying the model in* (S10). *Assume that the matrix*

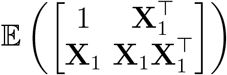

*is invertible, and that* 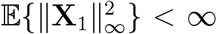. *Then, if λ* → 0 *and* 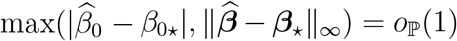 *as n* → *∞, it follows that* 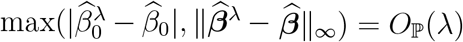

*Proof of Lemma S3*. We start by showing that

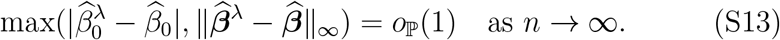

To this end, first note that under our conditions, 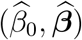 exists and is unique with probability one. Since 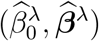 and 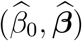 are respectively the maximizers of 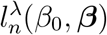 and *ℓ*_*n*_(*β*_0_, ***β***), we have

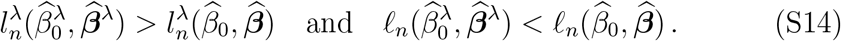

As

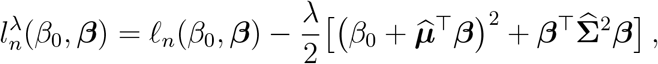

where 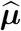 and 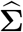 are defined as in (S7) with the ***x***_*i*_’s in the definition of the quantities *µ*_*n,j*_ and *s*_*n,j*_ replaced here by the random **X**_*i*_’s here, we have

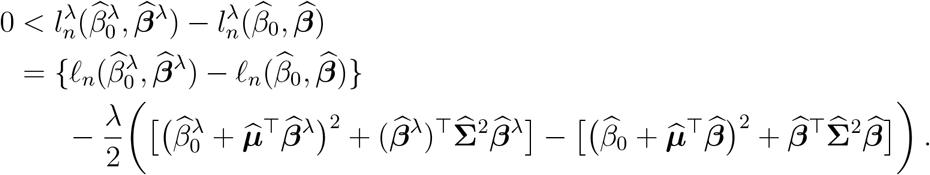

This implies that

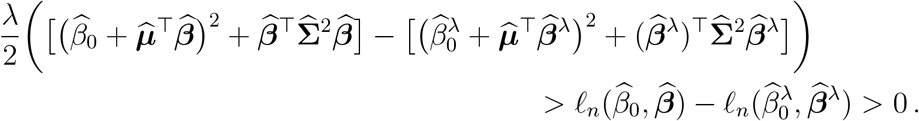

Since we have assumed that 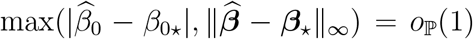 as *n* → ∞, as under our conditions the weak law of large numbers ensures *µ*_*n,j*_ = 𝔼 (X_*j*_) + *o* _ℙ_ (1), and since the weak law of large numbers combined with the continuous mapping theorem implies 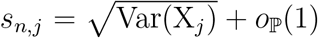, we conclude from the last equation display that as *n* → ∞,

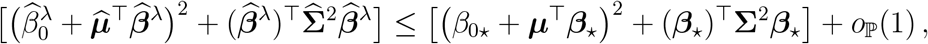

where ***µ*** = [𝔼 (X_1_), …, 𝔼 (X_*p*_)]^*⊤*^ and 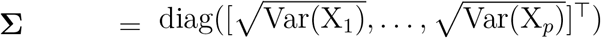.

As λ → 0 when *n* → ∞, and since 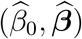 is bounded in probability as *n* → *∞*, we conclude from (S14) that 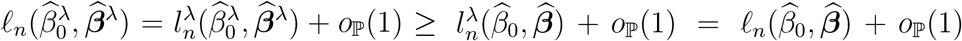. Therefore, 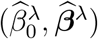 is a near-maximizer of *ℓ*_*n*_ (see e.g. [5] chapter 5), and we conclude that (S13) holds.

Next we show that the term on the right-hand side of the equality at (S13) can be replaced by *O*_ℙ_ (*λ*). To this end, note that we have from Remark S1 that

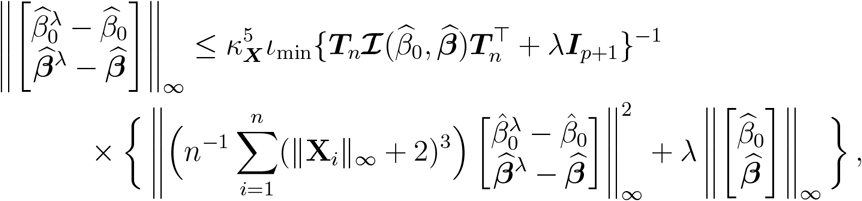

where

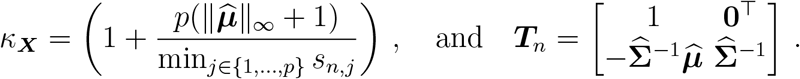

As for any *x, y* ∈ ℝ the mean value theorem ensures

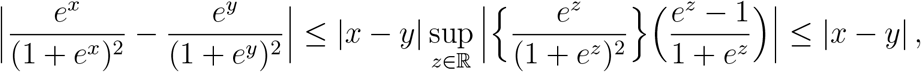

and since

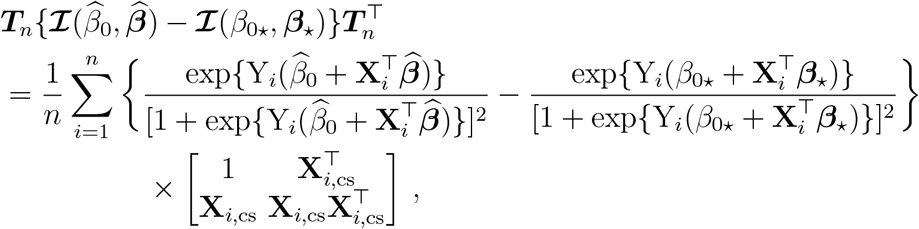

we deduce that

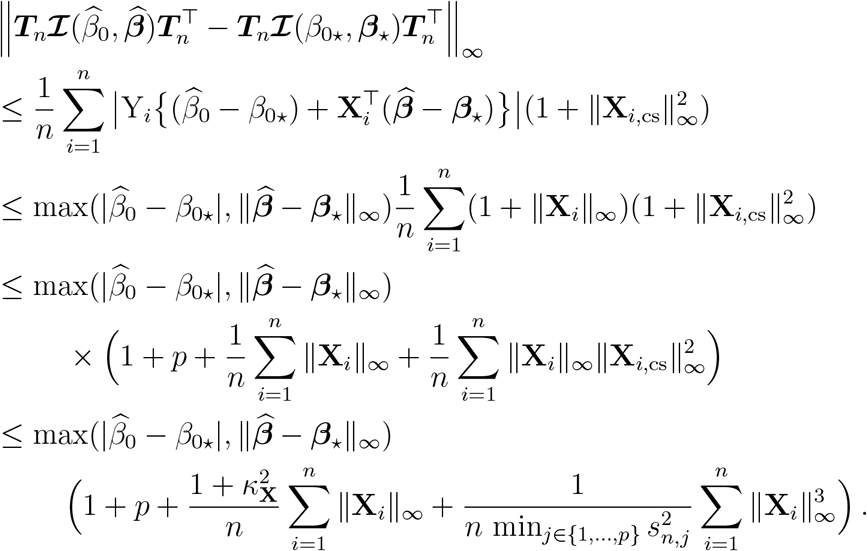

Since 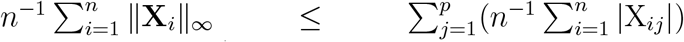 and 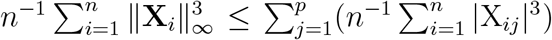, and as *p* is finite, we obtain from the weak law of large numbers that 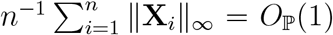 and 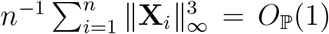 (recall that we have assumed 𝔼 (|X_*ij*_|^3^) < ∞ for all *j* ∈ {, *…, p*}). Furthermore, as we have established above that *µ*_*n,j*_ = 𝔼 (X_*j*_) + *o*_ℙ_ (1) and that 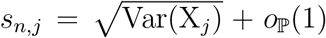, then, as *p* is finite, we get *κ*_**X**_ = *O*_ℙ_(1). Therefore, we conclude from the previous equation that

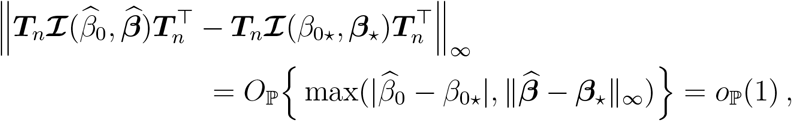

where the last equality followed from the fact that we have assumed 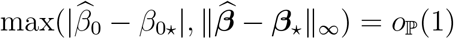 as *n* → ∞.

As the weak law of large numbers ensures, under our assumptions, that ***ℐ*** (*β*_0⋆_, ***β***_⋆_) = 𝔼{***ℐ*** (*β*_0⋆_, ***β***_⋆_)} + *o* _ℙ_ (1) as *n* → *∞*, and since ***T***_*n*_ = ***T*** + *o* _ℙ_ (1) with

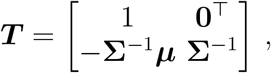

the assumption that

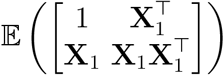

is invertible implies *ι*_min_{***T*** 𝔼{**ℐ** (*β*_0⋆_, ***β***_⋆_)}***T*** ^*⊤*^ + *λ****I***_*p*+1_} *≥ ι*_min_{***T*** E{**ℐ** (*β*_0⋆_, ***β***_⋆_)}***T*** ^*⊤*^} > 0. Therefore, we conclude from Slutsky’s lemma and the continuous mapping theorem that as *n* → ∞,

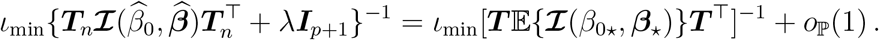

Therefore, as *n* → *∞*,

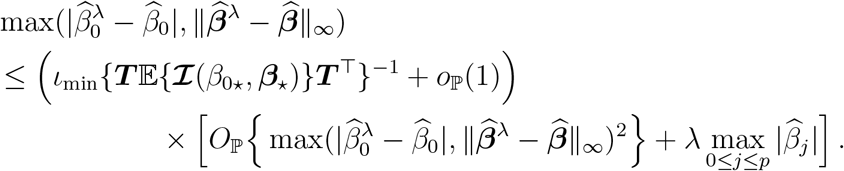

The proof follows from the fact that, as we have assumed 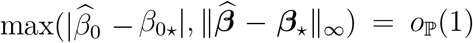 as *n* → *∞*, it follows that 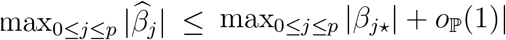.□

The preceding lemma implies that if λ = o(n^*−*1/2^), and if 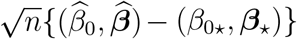 is asymptotically a mean-zero normal, then the penalized estimators used in our procedure can replace the unpenalized ones without affecting the asymptotic normality result. In other words, 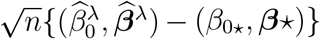 has the same asymptotic distribution than 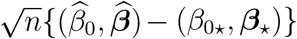.

Under our conditions, using arguments that are similar to those used in e.g. [5] chapter 5, under our conditions, the asymptotic variance-covariance matrix of 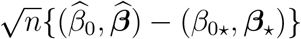 is given by

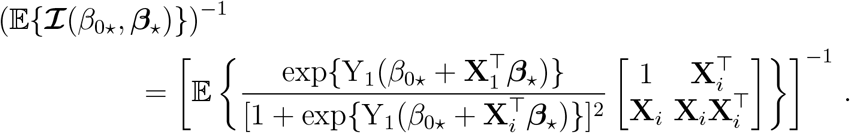

Inference on (*β*_0⋆_, ***β***_⋆_) based on the maximum likelihood estimator 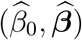 (for example, constructing confidence intervals or performing hypothesis tests) is typically carried out by combining these estimators with their corresponding standard errors. The standard deviation of 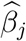, given by 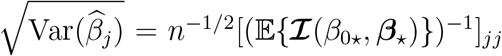 is commonly estimated using the following quantity:

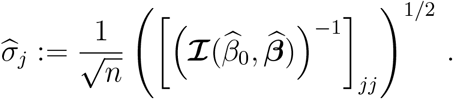

Based on Lemma S3, which implies that 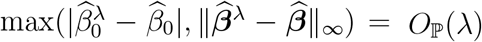 as *n*→ ∞, and using theoretical arguments similar to those employed therein, it is straightforward to show that, if *λ* = *o*(*n*^*−*1/2^), as *n* → *∞*,

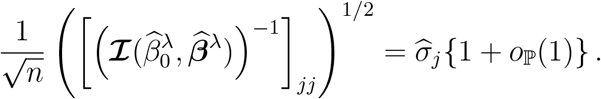

The proof of the consistency of our standard error computation procedure follows from the derivations provided in the following section, which show that 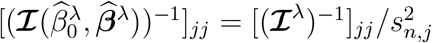, with ***ℐ***^λ^ defined at the beginning of Methods 4.2.3 in the manuscript.

#### 2.2 Computation of standard errors

Let (y_1_, ***x***_1_), …, (*y*_*n*_, ***x***_*n*_) to denote *n* independent realizations of the random pair (**X**, Y). We next describe the derivation of the expression for the estimates of standard errors.

As in the proof of Proposition S1, let 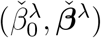 to denote the solutions of the following maximization problem:

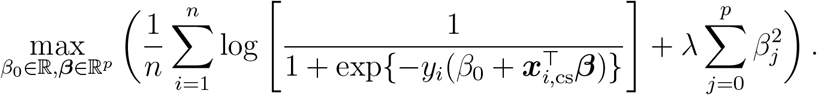

Then, for *j* ∈ {1, …, *p*}, one obtains from the relationship between 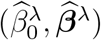 and 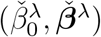 that

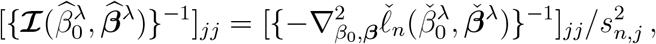

where

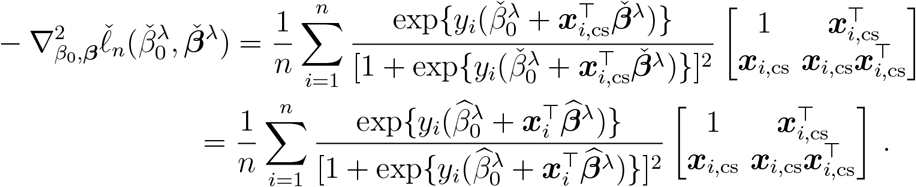

Now, recall that, for each *k* ∈ {1, …, *K*}, the vector 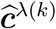 defined in (12) satisfies 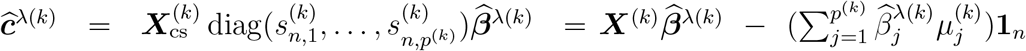, and that the response-node has access to 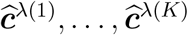. Since the response-node can also compute 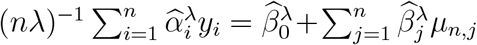 (recall the expression given in (11)), it is therefore able to compute

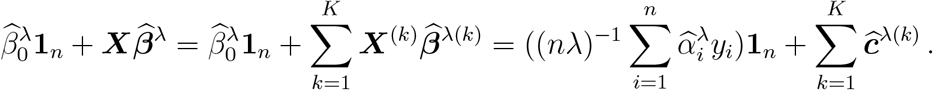

Then, upon defining a version 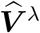 introduced above (7) with 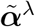 there replaced by 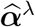, whose diagonal entries 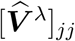 satisfying

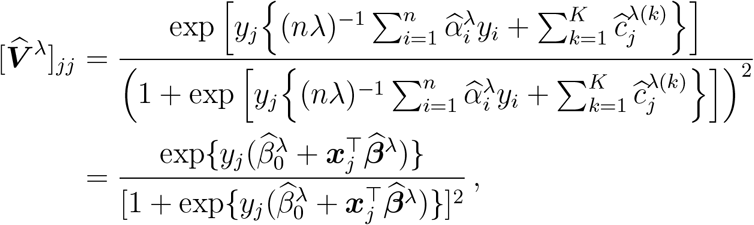

the 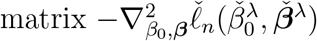 can be computed as

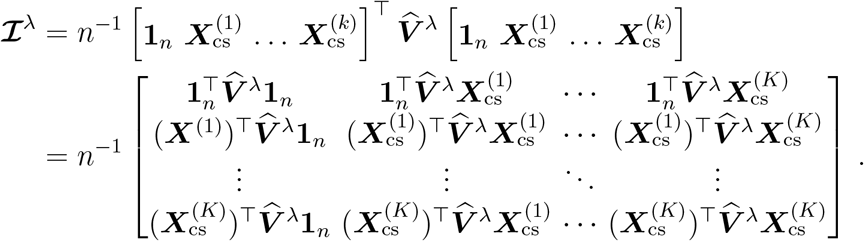

### Supplementary Methods 3 Selected box-constrained optimization algorithm and stopping criteria

#### 3.1 Two-metric projected newton algorithm

The convexity of the dual problem to solve at the response-node ensures that a unique solution exists on the domain of the objective function. The algorithm used to solve the problem should allow sufficient descent to reach an adequate approximation of this unique solution. Since the components of ***α*** are bounded by a compact set included in the open set (0, 1) (see Supplementary Methods 1), an algorithm adequate for box-constrained convex optimization problem had to be selected. While many methods exist for box-constrained optimization [6], the chosen method should allow to reach convergence with sufficient precision given the potentially small magnitude of the dual parameter ***α*** while still offering efficient computation when the dimension of the dual is high. We used the Two-Metric Projected Newton method suggested by Bertsekas [7], applicable because Lemma S1 ensures that the dual parameter estimates lie in a compact parameter space 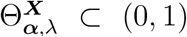. We refer to [8] for an extensive description of the method and convergence details. Briefly, all components of the estimate 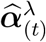 at step *t* at a boundary of the search domain and for which the gradient would pull the search toward the opposite side of the search domain are updated through gradient descent projected in the domain, while all other components are updated using Newton descent projected in the domain. The update is therefore 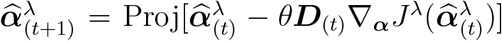, where ***D***_(*t*)_ depends of the component as described before and Proj[·] denotes the projection under the Euclidean norm. The step size *θ* is selected through back-tracking line search (Armijo rule) along projection arc detailed in [6, 8]. An initial admissible estimate has to be provided, which was set at 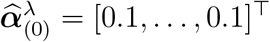.

#### 3.2 Stopping criteria

The error entailed by the approximation of 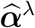 in the chosen algorithm should ideally be low enough such that it preserve the asymptotic properties derived for the primal estimate. We notice that λ holds a scaling role over the dual parameter ***α*** when it comes to retrieving the associated primal parameter ***β***. A restriction in function of λ consequently needs to be imposed in the estimation of the dual parameter to preserve the asymptotic properties of the primal parameters. The following proposition will allow to derive a stopping criteria for the dual estimation that ensures the asymptotic properties of the primal parameter hold.

##### Proposition S2.

>*For any* (*y*_1_, ***x***_1_), …, (*y*_*n*_, ***x***_*n*_) ∈ {*−*1, 1} *×* ℝ^*p*^ *and any* ϵ > 0, *consider* 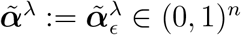 *such that*

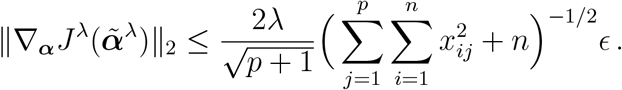

*Then*, 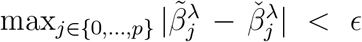, *where* 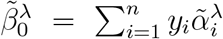 *and* 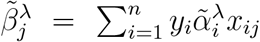 *for j* ∈ {1, …, *p*}, *and with the* 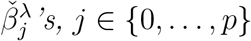, *defined as in Lemma S1*.

*Proof of Proposition S2*. Fix *ϵ* > 0, and let 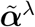 and 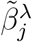, for *j* ∈ {0, …, *p*}, be as defined in the statement of the lemma. (Although 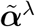 and the 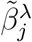’s implicitly depend on *ϵ*, this dependence is not explicitly reflected in the notation, for simplicity in the exposition.) For simplicity in the proof, for *i* ∈ {1, …, *n*} let *x*_0*i*_ = 1, so that 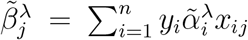 and 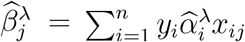 for *j* ∈ {0, …, *p*}.

Let 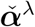 be defined as in Lemma S1, and recall from that lemma that 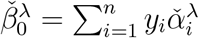 and 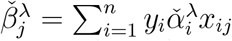 for *j* ∈ {1, …, *p*}.

Using the fact that *y*_*i*_ ∈ {*−*1, 1} for all *i* ∈ {1, …, *n*}, we derive

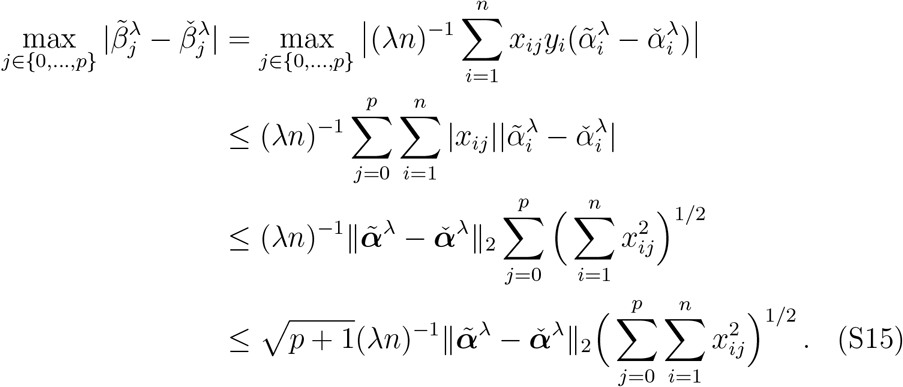

To obtain the one-to-last line, we used Cauchy-Schwartz inequality, and to obtain the last line, we used the fact that for any positive *a*_0_, …, *a*_*p*_ we have 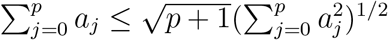

Now observe that, using standard vector calculus manipulations, the Hessian matrix of *J*^λ^(***α***) can be expressed as

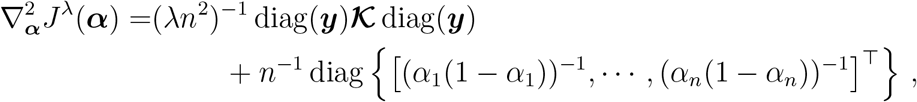

where we used the notation 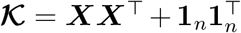 as defined in Supplementary Tables 1.

In the equation above, the matrix in the first term of the right-hand side of the equality is semi-positive definite, since for any vector 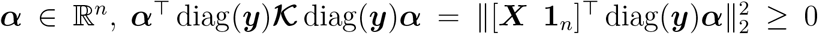. As the matrix n^*−*1^ diag {[(*α*_1_(1−*α*_1_))^*−*1^, · · ·, (*α*_*n*_(1 −*α*_*n*_))^*−*1^]^⊤^} is positive definite for all ***α*** ∈ (0, 1)^*n*^, with (*α*_*i*_(1*−α*_*i*_))^*−*1^ ≥ 4 for all *i* ∈{1, …, *n*}, it follows that 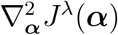 is strongly convex, with strong convexity parameter *m* = 4*n*^*−*1^, since it follows from the last discussion that the matrix

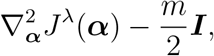

is positive definite.

This allows us to conclude as in e.g. [9], Section 9.1.2, p.459, that it holds for all ***α*** ∈ (0, 1)^*n*^ that

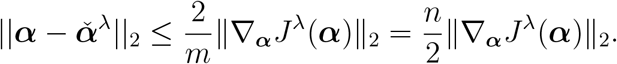

Combining this result with the inequality derived at (S15), we obtain

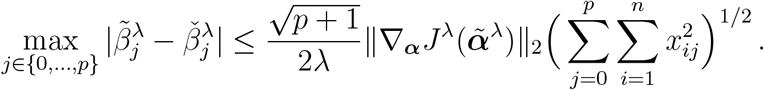

The proof of the lemma follows from the assumption over 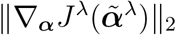.□

As shown in the proof of Proposition S1, the maximizer 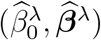 of 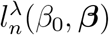, defined in (2) in the manuscript, satisfies

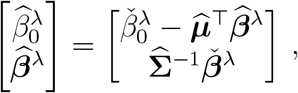

where 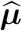 and 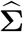 are defined in (S7), and where 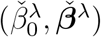, satisfies

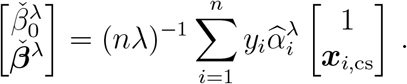

In the last equation, 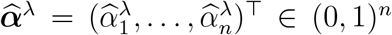 denotes the unique solution to the minimization problem 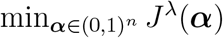, with *J*^*λ*^ as in (3).

From Proposition S2, if 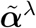 is a point such that

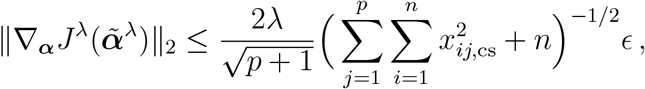

it follows that 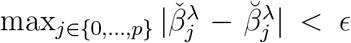, where 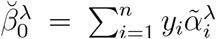 and 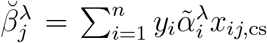 for *j* ∈ {1, …, *p*}. Therefore, if 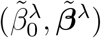 denotes a version of 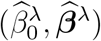 computed based on 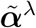 instead of 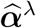, i.e, if 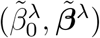 satisfies

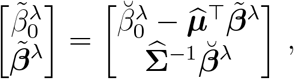

then, for *j* ∈ {1, …, *p*}, we have

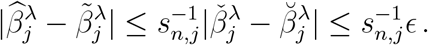

### Supplementary Methods 4 Privacy-preserving properties

#### 4.1 Proof of Proposition 1

Before proving Proposition 1, we state and prove the following lemma, which provides the foundation for the proof of Proposition 1. Let 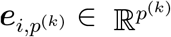 denote the standard basis vector with a 1 in the *i*^th^ position and 0 elsewhere.

##### Lemma S4.

*Let* ***A*** ∈ 𝕊 (**𝒦**^(*k*)^), *and consider* 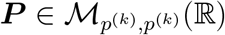 *such that* 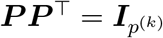. *Then, if* 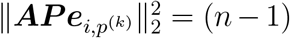 *for all* 1 *≤ i ≤ p*^(*k*)^ − 1, *we have* ***AP*** ∈ 𝕊 (**𝒦**^(*k*)^).

*Proof of Lemma S4*. First note that 𝕊(***K***^(*k*)^) is non-empty since 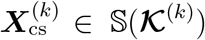. From this, to show that ***AP*** ∈ 𝕊 (**𝒦**^(*k*)^) whenever 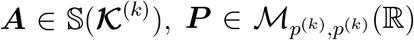 with 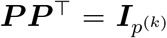 and 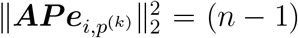 for all 1 *≤ i ≤ p*^(*k*)^ − 1, we need to verify that for such ***A*** and ***P*** we have (1) (***AP***)(***AP***)^⊤^ = ***K***^(*k*)^; (2) (***AP***)^⊤^**1**_*n*_ = 0; and (3) 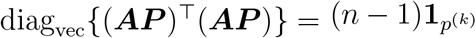.

To verify (1), it suffices to note that, since by definition ***AA***^⊤^ = **𝒦**^(*k*)^ and 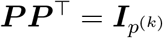, we have (***AP***)(***AP***)^⊤^ = ***A***(***P P*** ^⊤^)***A***^⊤^ = ***AA***^⊤^ = **𝒦**^(*k*)^.

To verify (2), since ***A*** ∈ 𝕊 (**𝒦** ^(*k*)^) implies ***A*** ^⊤^**1**_*n*_ = 0, one directly computes that (***AP***)^⊤^**1**_*n*_ = ***P*** ^⊤^(***A***^⊤^**1**_*n*_) = 0.

To verify (3), note that 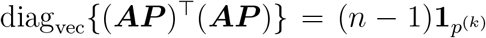 if and only if 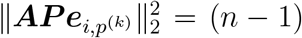 for all 1 *≤ i ≤ p*^(*k*)^. Since we have only assumed that 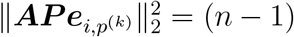 for all 1 *≤ i ≤ p*^(*k*)^ − 1, we need to prove that, under our conditions, 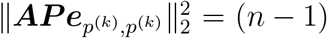. To see why this is the case, note that since 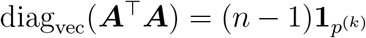, we have

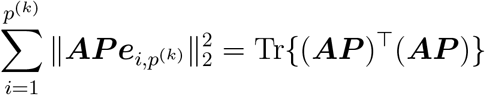

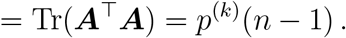

This implies that 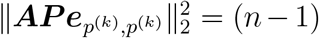, thereby concluding the proof of the proposition. □

*Proof of Proposition 1*. First note that since 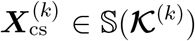, Lemma S4 implies that for any orthogonal matrix 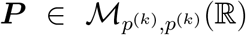 such that 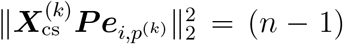 for all 1 *≤ i ≤ p*^(*k*)^ − 1, we have 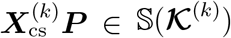. Since 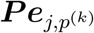 corresponds to the *j*^th^ column of ***P***, say ***p***_*j*_, which has unit norm, and each column of 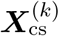 has squared Euclidian norm equal to *n* − 1, it follows that 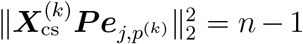 for all 1 *≤ j ≤ p*^(*k*)^ if and only if

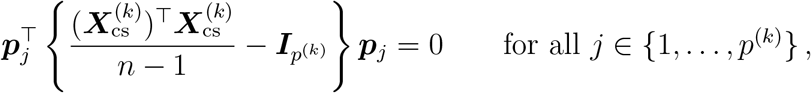

or equivalently, if and only if

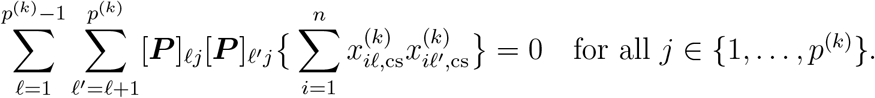

One of these equations is redundant, since 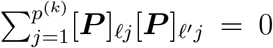 when *ℓ* = *ℓ*^*′*^, due to the orthogonality of the rows of ***P***. We conclude by dividing each side of the previous equation by *n* that if ***P*** is orthogonal and satisfies

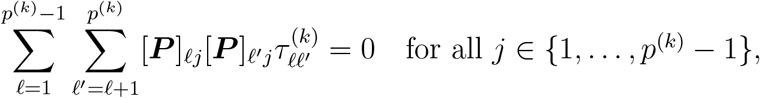

Then 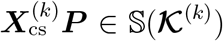. From this, to conclude the proof of the proposition, we need to show that any ***A*** ∈ 𝕊 (**𝒦**^(*k*)^) expresses as 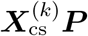, with ***P*** an orthogonal matrix such that 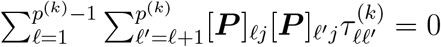 for all *j* ∈ {1, …, *p*^(*k*)^ ™ 1}.

First, from Theorem 7.3.11 in [10] (p.452), if 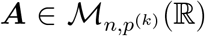 is such that 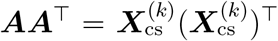, then there exists an orthogonal matrix 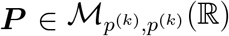 such that 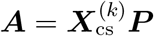.

The proposition result then follows from the fact that, since ***A*** ∈ 𝕊 (**𝒦** ^(*k*)^) implies 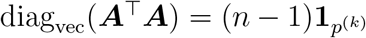, we have

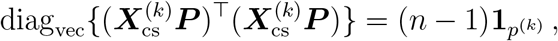

which implies that 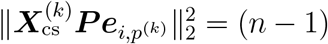 for all 1 *≤ i ≤ p*^(*k*)^ − 1. □

#### 4.2 Orthogonal matrices that preserve the binary nature of covariates

##### Proposition S3.

>*Let p*^(*k*)^ *≥* 2, *and for each j* ∈ {1, …, *p*^(*k*)^}, *define* 𝒟_*j*_ = {*a*_*j*_, *b*_*j*_} *with a*_*j*_ ≠ *b*_*j*_. *Assume that* 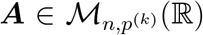 *satisfies* ***A***_*ij*_ ∈ 𝒟_*j*_ *for all i* ∈ {1, …, *n*} *and j* ∈ {1, …, *p*^(*k*)^}. *If* 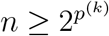 *and* ***A*** *contains exactly* 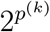 *distinct rows, then any orthogonal matrix* 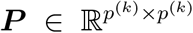 *such that* 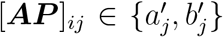 *for some* 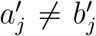 *for all i* ∈ {1, …, *n*} *and j* ∈ {1, …, *p*^(*k*)^} *must be a sign-permutation matrix*.

*Proof of Proposition S3*. Let 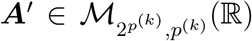 denote a submatrix of ***A*** consisting of 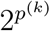 distinct rows. Assume without loss of generality that the rows of ***A***^*′*^ are arranged in a way that

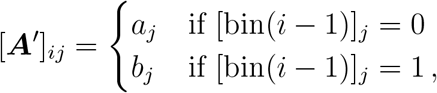

where, for any integer *i*, we use bin(*i*) to denote a vector containing its binary representation. In this notation, ***A***^*′*^ has the form

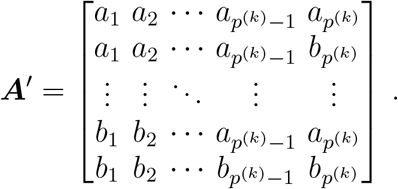

To prove the proposition, it suffices to show that, if 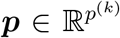 is a unit vector such that the entries of the vector ***A***^*′*^***p*** satisfy [***A***^*′*^***p***]_*i*_ ∈ {*r, s*} for some *r* ≠ *s*, for all 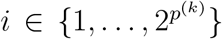, then ***p*** has exactly one non-zero entry. To do this, we proceed by induction on *p*^(*k*)^: we first show that the result is true for *p*^(*k*)^ = 2, then, we prove that if it is true for *p*^(*k*)^ −1, it implies that it is also true for *p*^(*k*)^.

In the case *p*^(*k*)^ = 2, ***A***^*′*^ satisfies

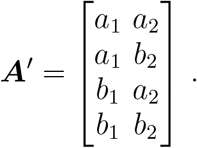

We then need to show that if ***p*** ∈ ℝ^2^ is a unit vector such that the entries of the vector ***A***^*′*^***p*** satisfy [***A***^*′*^***p***]_*i*_ ∈ {*r, s*} for some *r* ≠ *s*, for all *i* ∈ {1, …, 4}, then ***p*** has exactly one non-zero entry. To this end, assume that it is not the case and that both entries of ***p*** = [*p*_1_, *p*_2_]^⊤^ are non-zero. In this case, since *a*_*j*_ ≠ *b*_*j*_ for *j* ∈ {1, 2}, we have [***A***^*′*^***p***]_1_ − [***A***^*′*^***p***]_2_ ≠ 0, [***A***^*′*^***p***]_1_ − [***A***^*′*^***p***]_3_ ≠ 0 and [***A***^*′*^***p***]_2_ − [***A***^*′*^***p***]_4_ ≠ 0. As [***A***^*′*^***p***]_*i*_∈ {*r, s*} for some *r* ≠ *s*, we therefore have [***A***^*′*^***p***]_1_ = [***A***^*′*^***p***]_4_ and [***A***^*′*^***p***]_2_ = [***A***^*′*^***p***]_3_. These equalities implies that

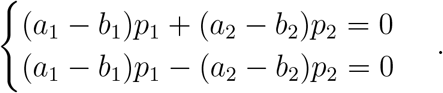

This system of equations shows a contradiction, since adding these equations implies (*a*_1_ − *b*_1_)*p*_1_ = 0, which is not possible since *a*_1_ ≠ *b*_1_ and we had assumed that *p*_1_ ≠ 0. This implies that ***p*** = [*p*_1_, *p*_2_]^⊤^ has at least one non-zero entry. Since ***p*** has norm equal to 1, and therefore has exactly one non-zero entry (equal to *±*1), which concludes the proof for the case *p*^(*k*)^ = 2.

We next show that if it is true for *p*^(*k*)^ − 1, it implies that it is also true for *p*^(*k*)^. To this end, first note that for any 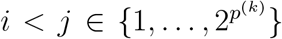 such that bin(*i* − 1) and bin(*j* − 1) differ by exactly one bit, say the ℓ^th^ one, we have by construction that

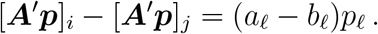

Now assume that all components of ***p*** are different from 0. By the last equation, this implies that for any 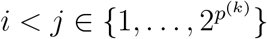 such that bin(*i* − 1) and bin(*j* − 1) differ by exactly one bit, we have

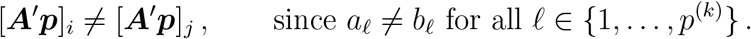

Within the first four elements of ***A***^*′*^***P***, this implies that [***A***^*′*^***p***]_1_≠ [***A***^*′*^***p***]_2_, [***A***^*′*^***p***]_1_≠ [***A***^*′*^***p***]_3_ and [***A***^*′*^***p***]_2_≠ [***A***^*′*^***p***]_4_, or again that [***A***^*′*^***p***]_1_ = [***A***^*′*^***p***]_4_ and [***A***^*′*^***p***]_2_ = [***A***^*′*^***p***]_3_ since [***A***^*′*^***p***]_*i*_ can only take one of two possible values {*r, s*}. By extending this logic, we deduce that for all *i* such that bin(*i*−1) has an even number of 1’s, we have [***A***^*′*^***p***]_1_ = [***A***^*′*^***p***]_*i*_, and [***A***^*′*^***p***]_1_≠ [***A***^*′*^***p***]_*i*_ otherwise. Specifically, with 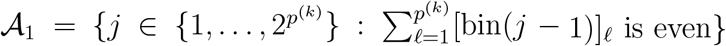 and 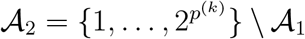, we have [***A***^*′*^***p***] = [***A***^*′*^***p***]_*j*_ for all *i, j*∈ 𝒜 _1_, and [***A***^*′*^***p***]_*i*_ = [***A***^*′*^***p***]_*j*_ for all *i, j* ∈ 𝒜_2_.

Now note that for any ℓ ≠ ℓ^*′*^ ∈ {1, …, *p*^(*k*)^}, there exists *i, j* ∈ 𝒜_1_ such that bin(*i* − 1) and bin(*j* − 1) are equal everywhere except in position ℓ and ℓ^*′*^, where their bits are flipped. Since *i, j*∈ 𝒜 _1_ implies [***A***^*′*^***p***]_*i*_ = [***A***^*′*^***p***]_*j*_, then

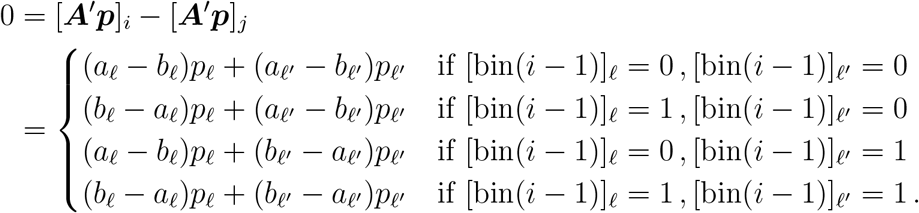

Now let *i*^*′*^, *j*^*′*^ be such that bin(*i* − 1) and bin(*i*^*′*^− 1) are identical except at position ℓ^*′*^, where their bits are flipped, and bin(*j* − 1) and bin(*j*^*′*^− 1) are identical except at position ℓ^*′*^, where their bits are flipped. Then, since *i, j*∈ 𝒜 _1_, we have *i*^*′*^, *j*^*′*^∈ 𝒜 _2_, which therefore implies 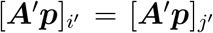 and that

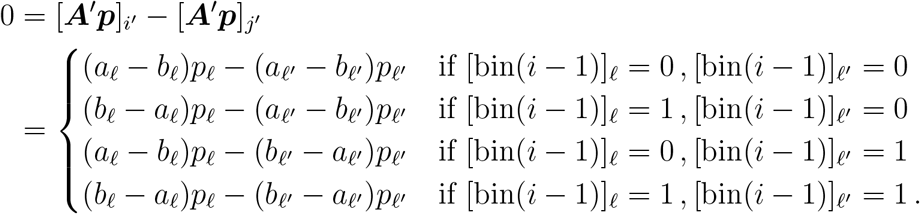

Adding the equations 0 = [***A***^*′*^***p***]_*i*_− [***A***^*′*^***p***]_*j*_ and 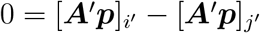 leads to a contradiction, as their validity would require that (*a*_ℓ_ − *b*_ℓ_)*p*_ℓ_ = 0. However, this cannot hold under the assumption that all *p*_ℓ_ are nonzero and *a*_ℓ_ ≠ *b*_ℓ_. Therefore, there must exist at least one component of ***p*** that is equal to 0. Without loss of generality, assume that this component is the first one, i.e., *p*_1_.

Consider the matrix ***Ā***^*′*^, obtained by removing the first column of ***A***^*′*^ and discarding its last 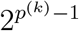 rows. Let 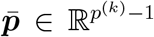 denote the vector obtained by removing the first component of ***p***. Then 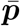 is a unit vector such that the entries of the vector 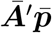 satisfy 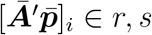 for some *r* ≠ *s* and for all 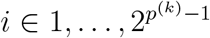. By the induction hypothesis, it follows that 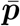 has exactly one nonzero component. Since 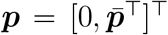, it follows ***p*** also has exactly one nonzero component. This concludes the proof that if 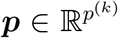 is a unit vector such that the entries of the vector ***A***^*′*^***p*** satisfy [***A***^*′*^***p***]_*i*_ ∈ {*r, s*} for some *r*≠ *s*, for all 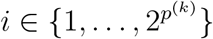, then ***p*** has exactly one non-zero entry. The result follows. □

#### 4.3 Proof of Proposition 2

Before establishing the proof of Proposition 2, consider the case *p*^(*k*)^ = 3. In this case, any orthogonal matrix ***P*** can be obtained as 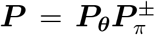 where, for ***θ*** = (*θ*_1_, *θ*_2_, *θ*_3_) ∈ [0, 2*π*),

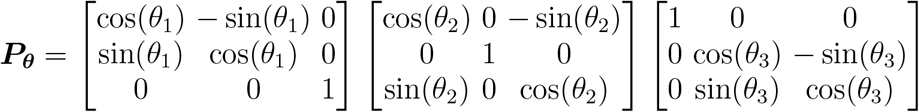

(see for example [11]). Matrices of the form ***P***_***θ***_ generate all orthogonal matrices ***P*** with determinant equal to 1. Since 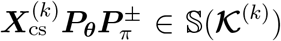 if and only if 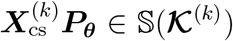, to characterize the matrices belonging in the set 𝕊 (**𝒦**^(*k*)^) it suffices to search for the values of ***θ*** ∈ (−*π, π*]^3^ such that 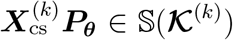. It follows from Proposition 1 that 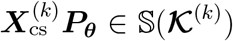 provided ***θ*** ∈ (−*π, π*]^3^ satisfies 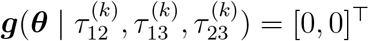, where

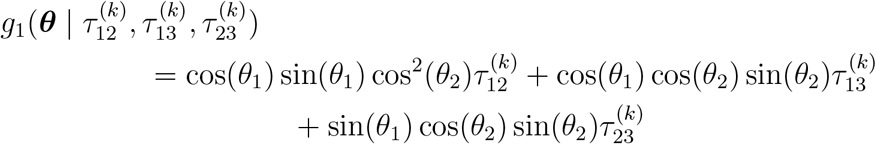

and

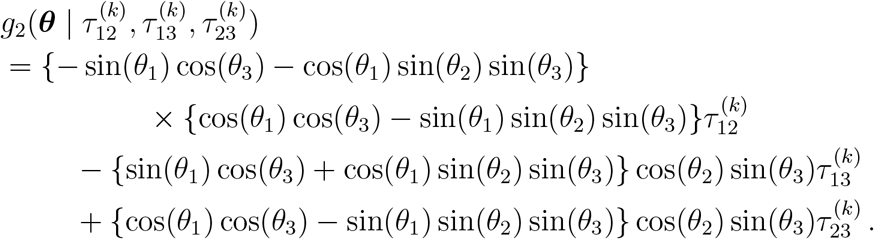

By the Implicit Function Theorem [12], since the system consists of two continuously differentiable equations and three unknowns, the existence of a solution in the interior of (*π, π*]^3^ implies that the system admits infinitely many solutions. Such a solution always exists, since ***θ***_0_ = **0** corresponds to 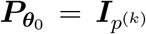 always satisfies 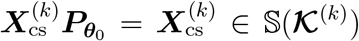. The rank *r* of the Jacobian matrix of 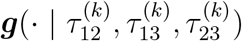 at the solution determines the dimension of the solution set, but regardless of the rank, the solution set is infinite: it is locally a manifold of dimension 3 − *r*.

The following proof formalizes this argument.

*Proof of Proposition 2*. By Proposition 1, ***A*** ∈ 𝕊 (**𝒦**^(*k*)^) if and only if 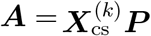, with 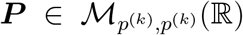 an orthogonal matrix satisfying (14). Moreover, any orthogonal 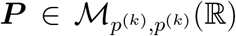 can be expressed as 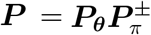, where ***P***_***θ***_ is a rotation matrix parametrized using the Givens rotation basis described in the statement of the proposition. Recalling the definition of ***g***(***θ***) in the statement of the proposition, and noting that ***A*** ∈ 𝕊 (**𝒦** ^(*k*)^) if and only if 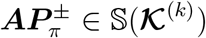, it suffices, to prove the proposition, to show that the set {***θ*** : ***g***(***θ***) = **0**} has infinite cardinality. This follows directly from the smoothness of ***g***(***θ***), the fact that when *p*^(*k*)^ *≥* 3 the dimension of ***θ*** exceeds the number of equations defined by ***g***(***θ***) = **0**, the existence of a solution 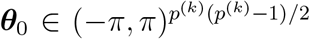 satisfying ***g***(***θ***_0_) = **0** (i.e., the solution ***θ*** = **0**, which corresponds to 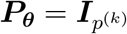), and an application of the Implicit Function Theorem. □

#### 4.4 Privacy assessment - Empirical criterion

An empirical criterion was derived to verify if, using the quantities available at the covariate-nodes, every entry of the response-node’s data can be flipped (recall ***y*** ∈ {− 1, 1} ^*n*^) while still leading to an admissible candidates for the response vector. This criterion, based on theoretical details from in Methods 4.3.2, is described in Algorithm 1 to support numerical implementation. We recall that the solution space that defines the solutions derived from the shared quantity ***ĉ***^*λ*(*k*)^ is given by

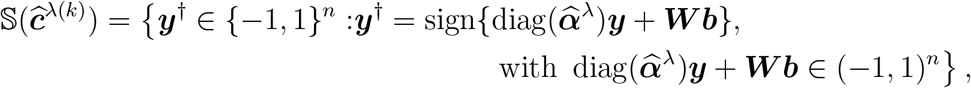

This criterion can be verified at the response-node for any covariate-node *k* not co-located at the response-node.

##### Algorithm 1 Empirical criterion for the privacy assessment of the response vector ***y*** at covariate-node *k*

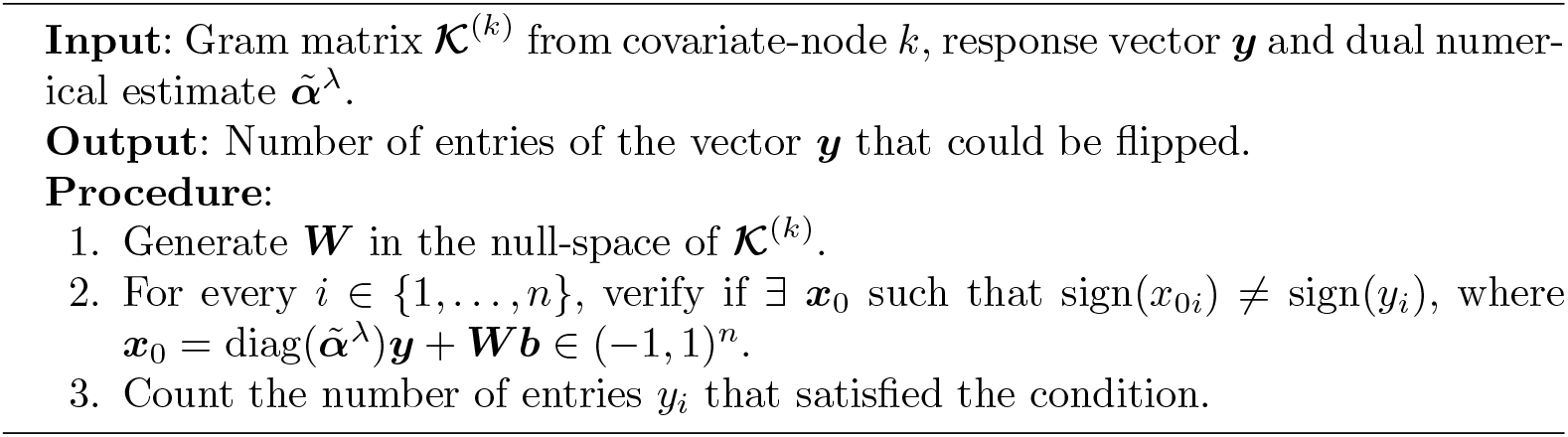

## Notes

### Competing Interest Statement

The authors have declared no competing interest.

### Funding Statement

Study funded by FRSQ / CIHR / NSERC

